# The Genetic Architecture of Amygdala Nuclei

**DOI:** 10.1101/2021.06.30.21258615

**Authors:** Mary S. Mufford, Dennis van der Meer, Tobias Kaufmann, Oleksandr Frei, Raj Ramesar, Paul M. Thompson, Neda Jahanshad, Rajendra A. Morey, Ole A. Andreassen, Dan J. Stein, Shareefa Dalvie

**Affiliations:** South African Medical Research Council Genomic and Precision Medicine Research Unit, Division of Human Genetics, Department of Pathology, Institute of Infectious Disease and Molecular Medicine, University of Cape Town, Cape Town, South Africa; Fellow, Global Initiative for Neuropsychiatric Genetics Education in Research (GINGER) program, Harvard T.H. Chan School of Public Health and the Stanley Center for Psychiatric Research at the Broad Institute of Harvard and MIT; NORMENT, KG Jebsen Centre for Psychosis Research, Division of Mental Health and Addiction, Oslo University Hospital & Institute of Clinical Medicine, University of Oslo, Oslo, Norway; School of Mental Health and Neuroscience, Faculty of Health, Medicine and Life Sciences, Maastricht University, Maastricht, The Netherlands; Tübingen Center for Mental Health, Department of Psychiatry and Psychotherapy, University of Tübingen, Tübingen, Germany; Center for Bioinformatics, Department of Informatics, University of Oslo, Oslo, Norway; Imaging Genetics Center, Stevens Neuroimaging and Informatics Institute, Keck School of Medicine of USC, Marina del Rey, CA, USA; Duke-UNC Brain Imaging and Analysis Center, Duke University, Durham, NC, USA; SA MRC Unit on Risk & Resilience in Mental Disorders, Department of Psychiatry and Neuroscience Institute, University of Cape Town, Cape Town, South Africa; South African Medical Research Council (SAMRC), Unit on Child & Adolescent Health, Department of Paediatrics and Child Health, University of Cape Town, Cape Town, South Africa

## Abstract

**Background:** Whereas a number of genetic variants influencing total amygdala volume have been identified in previous research, genetic architecture of its distinct nuclei have yet to be thoroughly explored. We aimed to investigate whether increased phenotypic specificity through segmentation of the nuclei aids genetic discoverability and sheds light on the extent of shared genetic architecture and biological pathways between the nuclei and disorders associated with the amygdala.

**Methods:** T1-weighted brain MRI scans (n=36,352, mean age= 64.26 years, 52% female) of trans-ancestry individuals from the UK Biobank were segmented into nine amygdala nuclei with FreeSurfer v6.1, and genome-wide association analyses were performed on the full sample and a European-only subset (n=31,690). We estimated heritability using Genome-wide Complex Trait Analysis, derived estimates of polygenicity, discoverability and power using MiXeR, and determined genetic correlations and shared loci between the nuclei using Linkage Disequilibrium Score Regression, followed by functional annotation using FUMA.

**Results:** The SNP-based heritability of the nuclei ranged between 0.17-0.33, and the central nucleus had the greatest statistical power for discovery. Across the whole amygdala and the nuclei volumes, 38 novel significant (p < 5×10^−9^) loci were identified, with most loci mapped to the central nucleus. The mapped genes and associated pathways revealed both unique and shared effects across the nuclei, and immune-related pathways were particularly enriched across several nuclei.

**Conclusions:** These findings indicate that the amygdala nuclei volumes have significant genetic heritability, increased power for discovery compared to whole amygdala volume, may have unique and shared genetic architectures, and a significant immune component to their aetiology.

## 1 Introduction

The amygdala is a subcortical brain region involved in emotional processing, social cognition, memory, and decision making (Amunts et al., 2005; Hortensius et al., 2016). Twin-based heritability of amygdala volume is approximately 43% (Hibar et al., 2015), with SNP-based heritability ranging between 9 to 17% (Satizabal et al., 2019). Thus far, only one locus on chromosome 12, rs17178006, has been significantly associated with amygdala volume (Satizabal et al., 2019). However, the genetic architectures of amygdala nuclei volumes have yet to be explored, which may contribute to our understanding of amygdala neurobiology.

Both human and animal models have identified three broad subdivisions of the amygdala, the basolateral, centromedial, and cortical-like complexes (Janak & Tye, 2015). Additionally, there remain nuclei that do not fit into these groups, including the anterior amygdaloid area, about which very little is known. These complexes have different functional connectivity and roles in threat processing (Janak and Tye, 2015). The basolateral amygdala consists of the lateral, basal, accessory basal and paralaminar nuclei. These nuclei evaluate sensory information and integrate with cortical association areas that regulate cognitive processing, fear and other emotional responses (Jovanovic et al. 2010). The centromedial amygdala consists of the central and medial nuclei and is critical for the orchestration of fear responses, such as increased cardiovascular output, via connections with the hypothalamus, basal forebrain, and brainstem (Janak & Tye, 2015). The cortical-like nuclei include the cortical nucleus and cortico-amygdaloid transition area, with roles in olfactory fear conditioning (Cádiz-Moretti et al., 2013) and social communication (Bzdok et al., 2013), respectively.

Reduced amygdala volume has been reported in several disorders, e.g. schizophrenia (Okada et al., 2016) and bipolar disorder (Hibar et al., 2016). A case-control comparison of schizophrenia and bipolar disorder localized these reductions to all nuclei, except the medial and central nuclei (Barth et al., 2021). Reduced volumes in patients may partly originate in genetics, making investigations into shared genetic architectures between brain volumes and disorders a valuable target for the clinical neurosciences. Studies investigating the genetics of subregion volume alterations in other brain regions e.g. the hippocampus (van der Meer et al., 2018), thalamus (Elvsåshagen et al., 2021) and brain stem (Elvsåshagen et al., 2020) have highlighted the potential of this approach through identification of loci associated with each of these structures and genetic overlap with a range of disorders.

Investigations of the genetic architecture of these nuclei are still needed to provide insights into the extent of shared genetic architecture and biological pathways between the nuclei and disorders to improve our understanding of the neurobiology of the amygdala and thereby the pathophysiology of associated disorders. We aimed to segment the amygdala into its nuclei and study the genetic underpinnings of amygdala nuclei volume alterations and associations with disorders, including implied genes and pathways. This was done through comprehensive analysis of heritability, genetic mixing, genetic correlations and shared loci.

## 2 Methods

### 2.1 Participants

Individual-level genotype and structural MRI data were sourced from the UK Biobank (UKB) (Bycroft et al., 2018; Miller et al., 2016) under accession code 27412. Each sample was collected with participants’ written informed consent and with approval by local institutional review boards. This dataset consisted of 42,067 participants of European, Asian, Chinese, African, mixed and other ancestries. Of these participants, 3,742 (8.9% of total) had an ICD-10 code corresponding to a neurological or mental disorder (codes F or G). These individuals were excluded from downstream analyses. The cohort’s age range is 44– 82 (mean age= 64.26 years, standard deviation (SD)= 7.5) and 52% of the participants are female. Figures illustrating the distributions of sex and age with amygdala volume are given in the supplementary information (SI) Figures S1 and S2, respectively.

### 2.2 MRI data processing

T1-weighted MRI volumes were processed using the standard FreeSurfer recon-all stream (v.5.3, http://surfer.nmr.mgh.harvard.edu). The amygdala was segmented into nine nuclei (anterior amygdaloid area, corticoamygdaloid transition area, basal, lateral, accessory basal, central, cortical, medial, and paralaminar nuclei) using the algorithm that was released as part of FreeSurfer v6.1 (Saygin et al., 2017) (Figure 1). The distribution of nuclei volumes is shown in Figure S3. The same algorithm has successfully been used to segment the hippocampus (Iglesias et al., 2015; van der Meer et al., 2018). This algorithm employs Bayesian inference combined with an amygdala atlas, created through the manual delineation of ultra-high resolution (100-150µm) images at 7T field strength of ex-vivo amygdala tissue (Saygin et al., 2017). Individuals ±4 SD from the mean of the Euler number were excluded from further analyses (Rosen et al., 2018).

**Figure 1:**
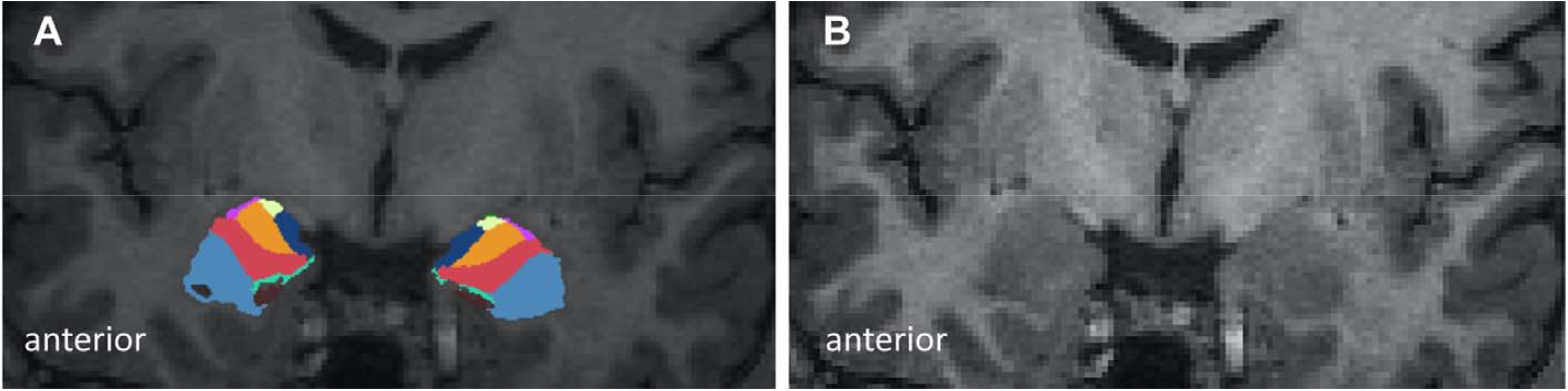
Segmentation of the amygdala nuclei. A) Using FreeSurfer v6.1 the amygdala was segmented into nine nuclei: anterior amygdaloid area = yellow, corticoamygdaloid transition area = dark blue, basal = red, lateral= light blue, accessory basal = orange, central= purple, medial = green, cortical and paralaminar nuclei. The cortical and paralaminar nuclei are not shown here. (B) Structural T1 scan provided for reference. Images provided by Morey et al., 2012 (1)

### 2.3 Genotyping and quality control

Phased and imputed genome-wide genetic data was obtained from the UKB (version 3) (Bycroft et al., 2018) using SHAPEIT3 (O’Connell et al., 2016) and the 1000 Genomes phase 3 dataset (Auton et al., 2015), respectively. Initially, participants were restricted to European ancestry as determined through self-report and validated with principal component analysis (PCA) by the UKB (Reich et al., 2008). We also included these European ancestry participants, self-reported European participants (that did not meet PCA requirements), African, Asian, Chinese, mixed and other ancestries in a trans-ancestry analysis. Post-imputation quality checks included the removal of poorly imputed SNPs (estimated R^2^<0.5), SNPs with low minor allele frequency (<0.1%), or SNPs that were not in Hardy-Weinberg Equilibrium (p<1×10^−9^). These filters were applied using PLINK version 1.9 (Chang et al., 2015). The number of SNPs remaining after each filtering step is shown in Table S1.

### 2.4 Statistical analyses

As volumetric (Pearson’s *r*_*g*_ between 0.54-0.86, all p < 2.2×10^−16^ across nuclei) and genetic (Pearson’s *r*_*g*_ between 0.56-0.91, all p < 5.27×10^−11^ across nuclei) correlations between right and left-brain hemispheres were relatively high for most structures, and to reduce the number of analyses, we summed the estimates of both hemispheres together. Heritability estimates and genetic correlations for each of the hemispheres (Table S2) were calculated using linkage disequilibrium score regression (LDSC) (Bulik-Sullivan, Loh, et al., 2015). Generalized additive model (GAM)-fitting in R (v3.5) (R Development Core Team, 2018) on the total sample was used to regress out the effects of the covariates from each outcome measure. The covariates were scanner, sex, age, age^2^, the first ten principal components to account for population stratification and genotyping artifacts, intracranial volume (ICV), and whole amygdala volume. Whole amygdala volume was included as a covariate to isolate the contribution from the unique genetic architecture of each nucleus. The Manhattan plots, QQ-plots and significant associations without this correction for the whole amygdala volume are displayed in Figure S4 and Table S3. We further removed all individuals ±4 SD from the mean on any of the amygdala measures or ICV. For the trans-ancestry analysis, self-reported ancestry was also included as a covariate. The number of participants remaining after each filtering step is shown in Table S1. Bonferroni correction was used to account for multiple testing. The total number of tests amounted to ten, considering the nine nuclei volumes and whole amygdala volume (α=5×10^−9^). These analyses were performed in R (v3.6). Scripts are available from the corresponding author.

### 2.5 Genome-wide association analyses

After quality control, 31,690 participants and 12,245,112 SNPs remained in the European dataset. For the trans-ancestry analyses, 36,352 participants and 9,915,367 SNPs remained after quality control. The trans-ancestry dataset consisted of 97% European (10% of which were not validated with PCA) and 3% non-European participants, including African, Chinese, Asian, mixed and other ancestries (Table S4 and Figure S5). GWAS was performed using PLINK version 1.9 for the amygdala and its nuclei, using the GAM-residualized volume estimates. Loci were defined with r^2^>0.1 and a genomic window of 250kb, which were deemed significant after correction for multiple testing (p< 5×10^−9^).

### 2.6 Functional annotation

The Functional Mapping and Annotation of Genome-Wide Association Studies (FUMA) platform was used for functional annotation of the GWAS results, with default settings (Watanabe et al., 2017). FUMA maps the top SNP associations to genes based on position. Subsequently, through hypergeometric testing, these genes are investigated for enrichment of biological processes, tissue, and cell types and whether they have previously been associated with traits in the GWAS catalogue.

### 2.7 SNP-based heritability

Genome-wide Complex Trait Analysis (GCTA) (Yang et al., 2011) was used to calculate the SNP-based heritability for each of the GAM-residualized nuclei volume estimates and additional subcortical regions of interest. Other subcortical regions were included as validation to be compared with previous findings. GCTA employs a restricted maximum likelihood (REML) approach using individual-level data. Regions with high linkage disequilibrium (LD) were pruned before analysis, using a sliding window approach with a window size of 50, a step size of 5, and an r^2^ of 0.2. An adjustment for cryptic relatedness was also applied with a threshold of 0.05 (Zaitlen et al., 2013), excluding 1,081 participants from the European analysis and 1,848 participants from the trans-ancestry analysis. To test whether heritability estimates and nucleus volume are correlated, we performed a simple linear regression between the two variables. Further, to validate the estimates from GCTA, SNP-based heritability was also calculated using LDSC (Bulik-Sullivan, Loh, et al., 2015).

### 2.8 Genetic overlap between nuclei, other subcortical regions and other traits

The summary statistics from the European mega-analysis were used to determine genetic correlation between the amygdala nuclei and selected subcortical volumes using cross-trait LDSC (Bulik-Sullivan, Finucane, et al., 2015). LD Hub (using version 1.9.3) (http://ldsc.broadinstitute.org/ldhub/) (Zheng et al., 2016) was used to calculate pairwise genetic correlations between the amygdala nuclei and 190 non-UKB traits of interest, including pathological and anthropometric categories. We restricted the analyses to studies of European ancestry, as this type of analysis is currently best suited to compare studies of matched ancestry (Zheng et al., 2016).

### 2.9 Estimating polygenicity, discoverability, power and residual inflation

To estimate the proportion of causally associated SNPs (polygenicity), the effect size variance (discoverability), the power to detect causal variants, and elevation of z-scores due to residual inflation, we used univariate MiXeR (version 1.2) (Frei et al., 2019; Holland et al., 2020). This model utilizes GWAS summary statistics and detailed LD structure of a reference panel and assumes a Gaussian distribution of effect sizes at a fraction of SNPs randomly distributed across the autosomal genome. Based on how closely the data follows the predicted model in the QQ plots (Figure S6) and the positive Akaike Information Criterion (AIC) values (Table 1), the additional complexity of the MiXeR model is justified as compared to LDSC (Frei et al., 2019; Holland et al., 2020). All z-score estimates are close to one, indicating minimal global inflation.

**Table 1:** MiXeR model fit and parameter estimations.

| Analysis | Region of interest | Polygenicity | Discoverability | Elevation of z-scores | $h^2$ | AIC | nc@p9 | Predicted number of causal SNPs |
| --- | --- | --- | --- | --- | --- | --- | --- | --- |
| **European meta-analysis** | Whole amygdala | $5.50 \times 10^{-04}$ | $2.06 \times 10^{-04}$ | 1.05 | 0.23 | 84.78 | 1,754.26 | $5.25 \times 10^3$ |
| | Anterior amygdaloid area | $4.93 \times 10^{-04}$ | $1.92 \times 10^{-04}$ | 1.04 | 0.20 | 45.50 | 1,573.51 | $4.71 \times 10^3$ |
| | Accessory basal nucleus | $7.59 \times 10^{-04}$ | $1.23 \times 10^{-04}$ | 1.04 | 0.19 | 34.78 | 2,420.71 | $7.24 \times 10^3$ |
| | Basal nucleus | $8.28 \times 10^{-04}$ | $1.20 \times 10^{-04}$ | 1.05 | 0.21 | 36.95 | 2,639.76 | $7.90 \times 10^3$ |
| | Corticoamygdaloid area | $3.41 \times 10^{-04}$ | $1.37 \times 10^{-04}$ | 1.03 | 0.10 | 16.27 | 1,088.85 | $3.25 \times 10^3$ |
| | Cortical nucleus | $8.14 \times 10^{-04}$ | $9.49 \times 10^{-05}$ | 1.03 | 0.16 | 16.67 | 2,595.32 | $7.77 \times 10^3$ |
| | Central nucleus | $1.09 \times 10^{-04}$ | $4.92 \times 10^{-04}$ | 1.03 | 0.11 | 119.72 | 344.50 | $1.04 \times 10^3$ |
| | Lateral nucleus | $9.43 \times 10^{-04}$ | $1.05 \times 10^{-04}$ | 1.05 | 0.20 | 27.82 | 3,006.38 | $9.00 \times 10^3$ |
| | Medial nucleus | $4.94 \times 10^{-04}$ | $1.05 \times 10^{-04}$ | 1.03 | 0.11 | 12.10 | 1,576.79 | $4.72 \times 10^3$ |
| | Paralamina nucleus | $1.19 \times 10^{-03}$ | $7.44 \times 10^{-05}$ | 1.03 | 0.18 | 15.47 | 3,808.80 | $1.14 \times 10^4$ |
| **Trans-ancestry analysis** | Whole amygdala | $5.53 \times 10^{-04}$ | $1.88 \times 10^{-04}$ | 1.06 | 0.22 | 100.16 | 1,763.74 | $5.28 \times 10^3$ |
| | Anterior amygdaloid area | $6.93 \times 10^{-04}$ | $1.35 \times 10^{-04}$ | 1.04 | 0.19 | 55.72 | 2,209.33 | $6.61 \times 10^3$ |
| | Accessory basal nucleus | $6.39 \times 10^{-04}$ | $1.38 \times 10^{-04}$ | 1.05 | 0.18 | 52.01 | 2,039.44 | $6.10 \times 10^3$ |
| | Basal nucleus | $6.66 \times 10^{-04}$ | $1.33 \times 10^{-04}$ | 1.05 | 0.18 | 51.49 | 2,125.33 | $6.36 \times 10^3$ |
| | Corticoamygdaloid area | $3.71 \times 10^{-04}$ | $1.21 \times 10^{-04}$ | 1.04 | 0.09 | 16.50 | 1,183.44 | $3.54 \times 10^3$ |
| | Cortical nucleus | $7.05 \times 10^{-04}$ | $1.01 \times 10^{-04}$ | 1.04 | 0.15 | 20.50 | 2,248.06 | $6.73 \times 10^3$ |
| | Central nucleus | $8.14 \times 10^{-05}$ | $5.14 \times 10^{-04}$ | 1.05 | 0.09 | 152.78 | 259.46 | $7.77 \times 10^2$ |
| | Lateral nucleus | $8.50 \times 10^{-04}$ | $1.04 \times 10^{-04}$ | 1.05 | 0.18 | 32.33 | 2,710.38 | $8.11 \times 10^3$ |
| | Medial nucleus | $8.61 \times 10^{-04}$ | $6.80 \times 10^{-04}$ | 1.02 | 0.12 | 18.11 | 2,744.28 | $8.22 \times 10^3$ |
| | Paralamina nucleus | $1.20 \times 10^{-03}$ | $6.75 \times 10^{-5}$ | 1.03 | 0.17 | 16.00 | 3,825.68 | $1.15 \times 10^4$ |

$h^2$ , heritability estimate; AIC, Akaike Information Criterion; SNPs, single nucleotide polymorphisms. The number of causal SNPs is determined by polygenicity x nSNP. The number of reference SNPs included in this model is 9,545,381.

## 3 Results

### 3.1 SNP-based Heritability

Heritability estimates for the European dataset, determined using GCTA, were all statistically significant (p<1×10^−16^, on the diagonal of Figure 2 and Table S5). The heritability estimate for the whole amygdala volume was 0.37 and the heritability estimates for the nuclei volumes ranged from 0.17 to 0.33 for the corticoamygdaloid transition area and accessory basal nucleus volumes, respectively (Figure 2 and Table S5). The heritability estimates determined with LDSC (Table S5) were approximately 37% lower than those estimated with GCTA.

**Figure 2:**
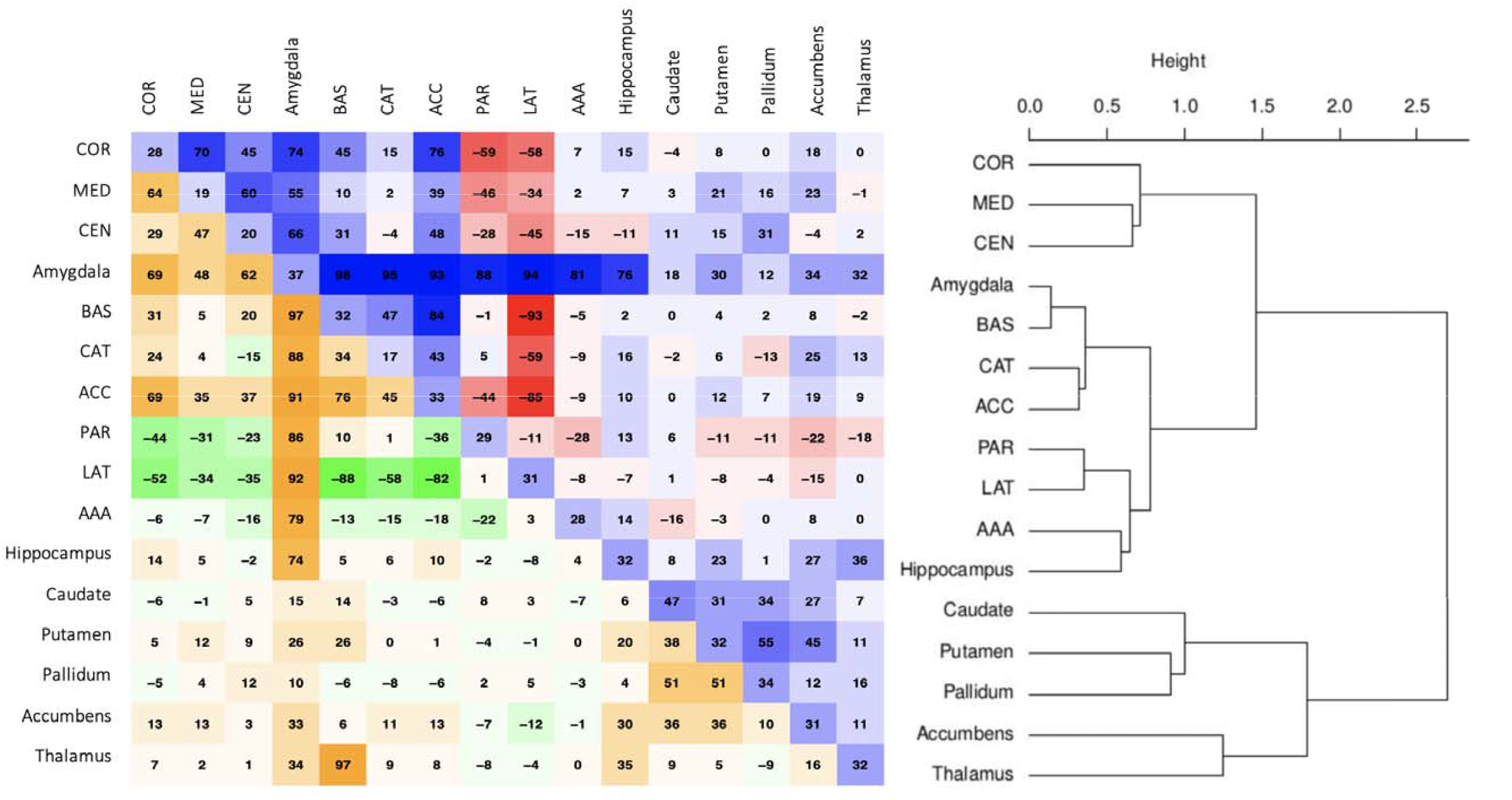
Correlation matrix of the volume estimates for the nuclei as well as several other subcortical regions of interest. All correlations are multiplied by a factor 100. The volumetric correlations are shown in the lower triangle of the matrix (green-orange), the heritability estimates on the diagonal, and the genetic correlations in the upper triangle (blue-red). This heatmap was generated using *corrplot* (2) in R (v3.6). The order, indicated by the dendrogram on the left, is determined by hierarchical clustering using Ward’s D2 method. COR= cortical nucleus, MED= medial nucleus, CEN= central nucleus, BAS= basal nucleus, CAT= corticoamygdaloid transition area, ACC= accessory basal nucleus, PAR= paralaminar nucleus, LAT= lateral nucleus, AAA= anterior amygdaloid area.

**Figure 3:**
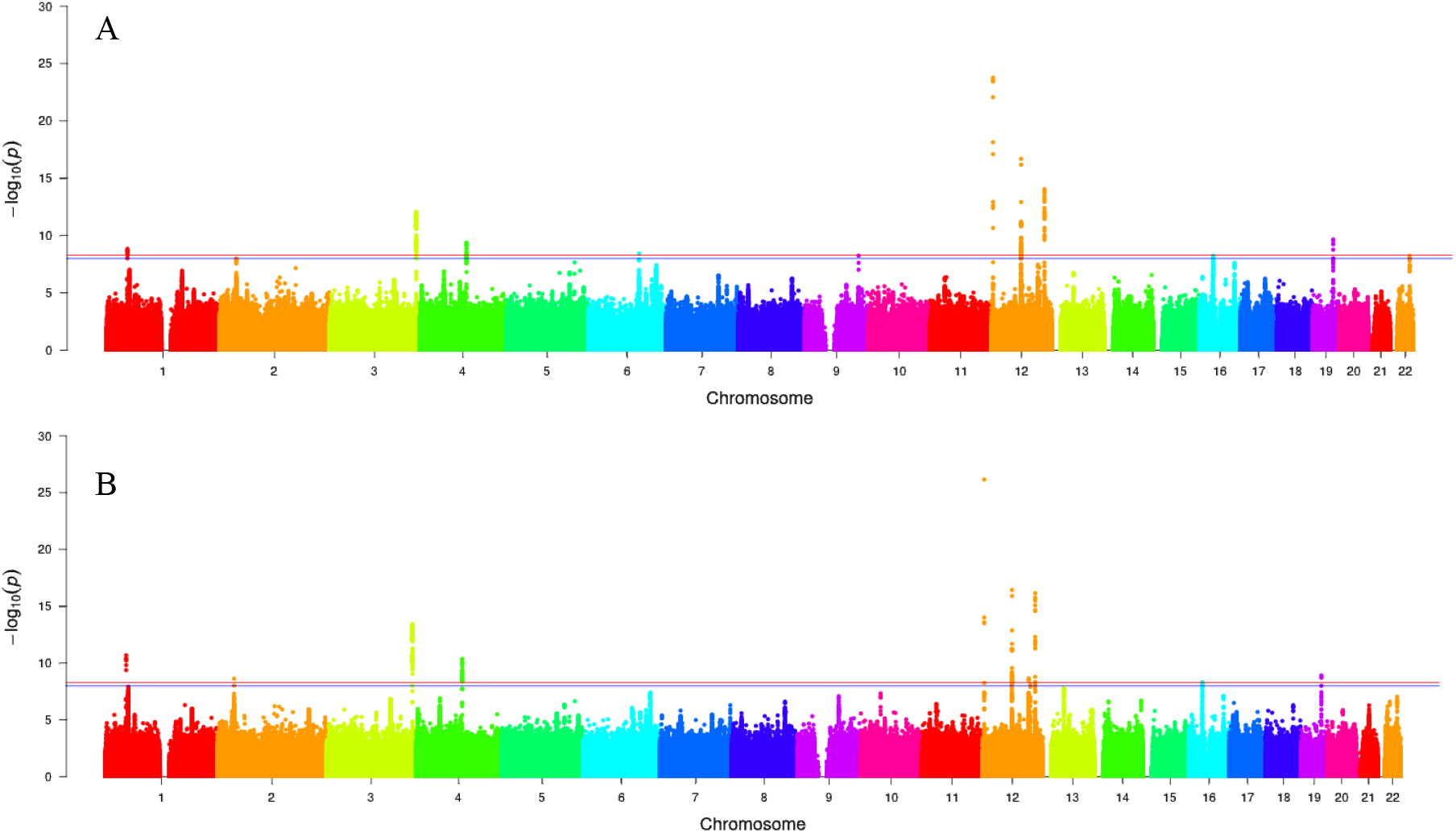
Manhattan plots for whole amygdala GWAS across European and trans-ancestry datasets. The chromosomal position is shown on the x-axis and log_10_-transformed p-values are on the y-axis. The red dashed line represents the Bonferroni corrected p-value threshold (p=5×10^−9^) and the blue dashed line represents the standard genome-wide-significance significance threshold (p=5×10^−8^). A) Manhattan plot for the European meta-analysis, B) Manhattan plot for the trans-ancestry dataset. The plots were generated using *ggplot2* (3) in R (v3.6).

### 3.2 GWAS of the amygdala nuclei volumes

Our whole amygdala volume mega-analysis identified eight independent genome-wide significant loci (Figure 2A, Table 2, Table T5), of which seven loci are novel. Of the variants identified here 83% had a p<0.05 and the same direction of effect in the GWAS of amygdala volume conducted by Satizabal et al., 2019. Our GWAS for the nuclei volumes identified an additional 21 novel significant loci (Table 2). We did not observe a correlation (r^2^=0.001) between the number of significant loci and each nucleus’ average volume (Figure S9). The majority of these associated loci are unique to specific nuclei and are predominantly intergenic or intronic. Only the missense variant, rs13107325, is shared between the whole amygdala and six of the nuclei volumes.

**Table 2:**
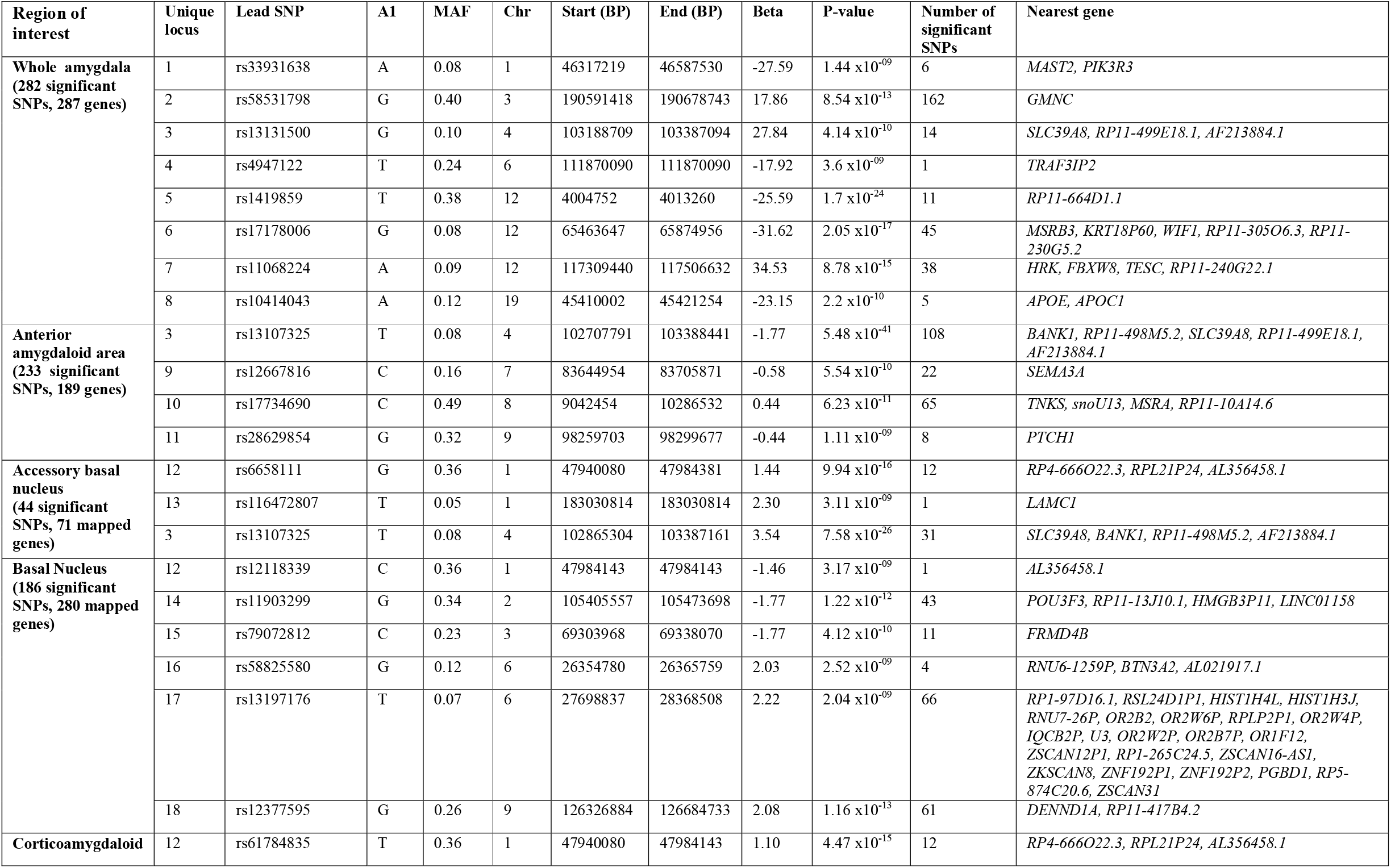
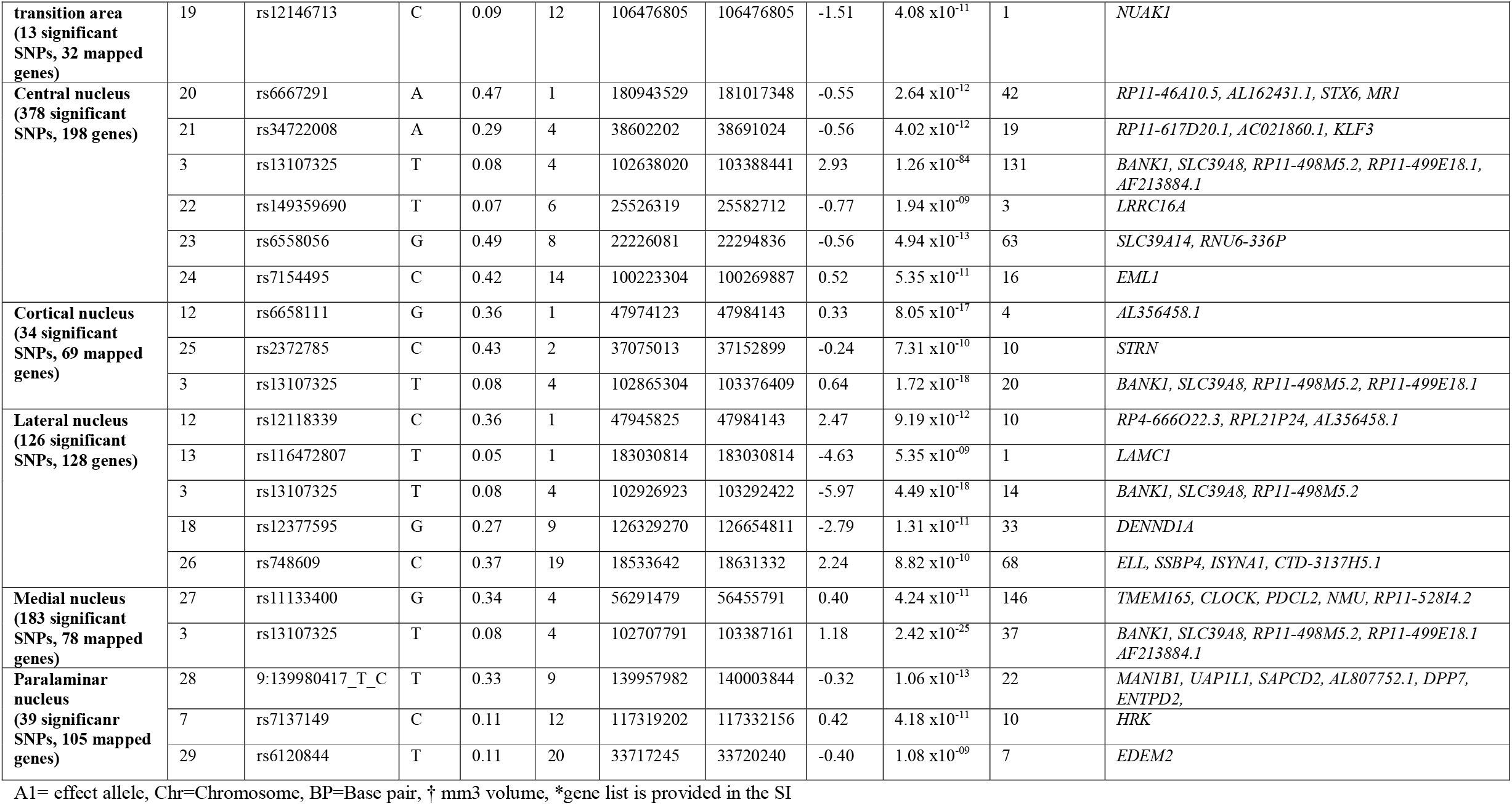
Genome-wide significant loci for whole amygdala and nuclei volumes from the European meta-analysis.

The trans-ancestry GWAS of the whole amygdala volume identified ten significant loci (Figure 2B, Table 3, Table T5, Figure S10). Of these, seven were shared with the European mega-analysis. Two of these shared loci (rs33931638, rs11068224) had decreased p-values in relation to the European analysis. A consistent direction of allelic effect was observed across significant loci that the trans-ancestry results shared with the European sample results. An additional 29 significant loci were identified across the nine nuclei volumes. More associations were identified for most nuclei in the trans-ancestry analysis than the European mega-analysis, except for the cortical nucleus, which fully replicated the European analysis results. Twenty-five of the 29 significant associations from the European analysis were also observed in the trans-ancestry analysis.

**Table 3:**
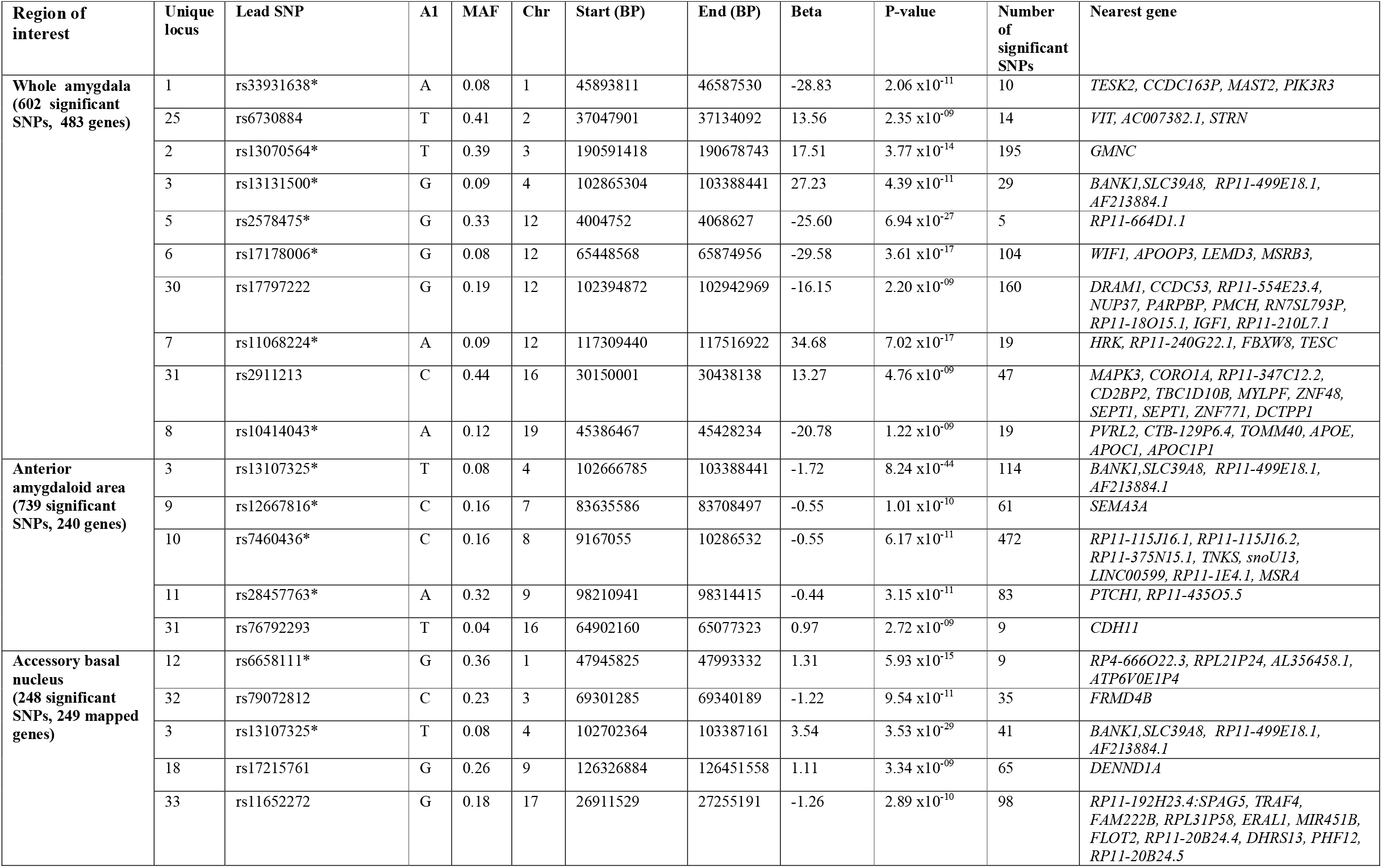
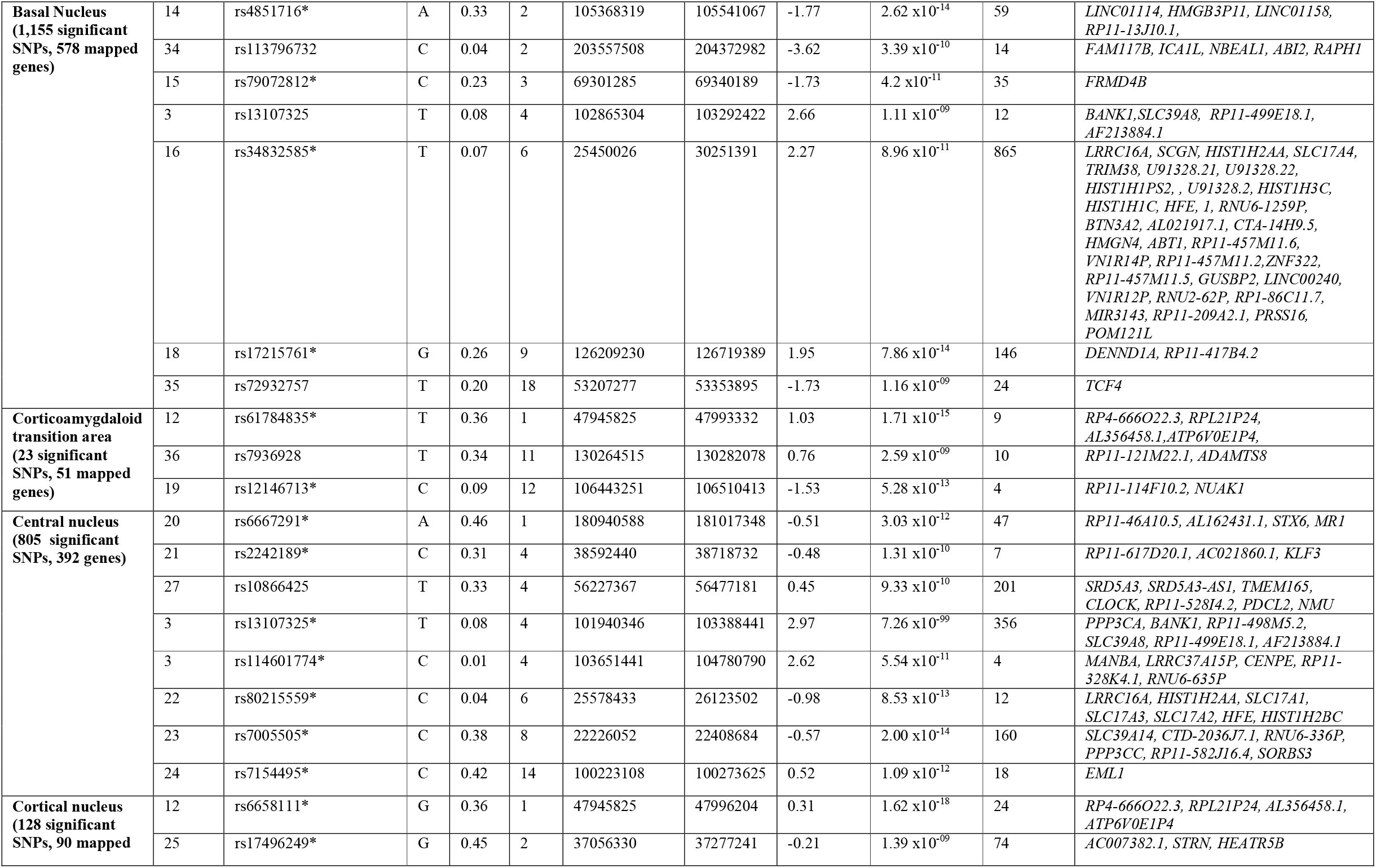
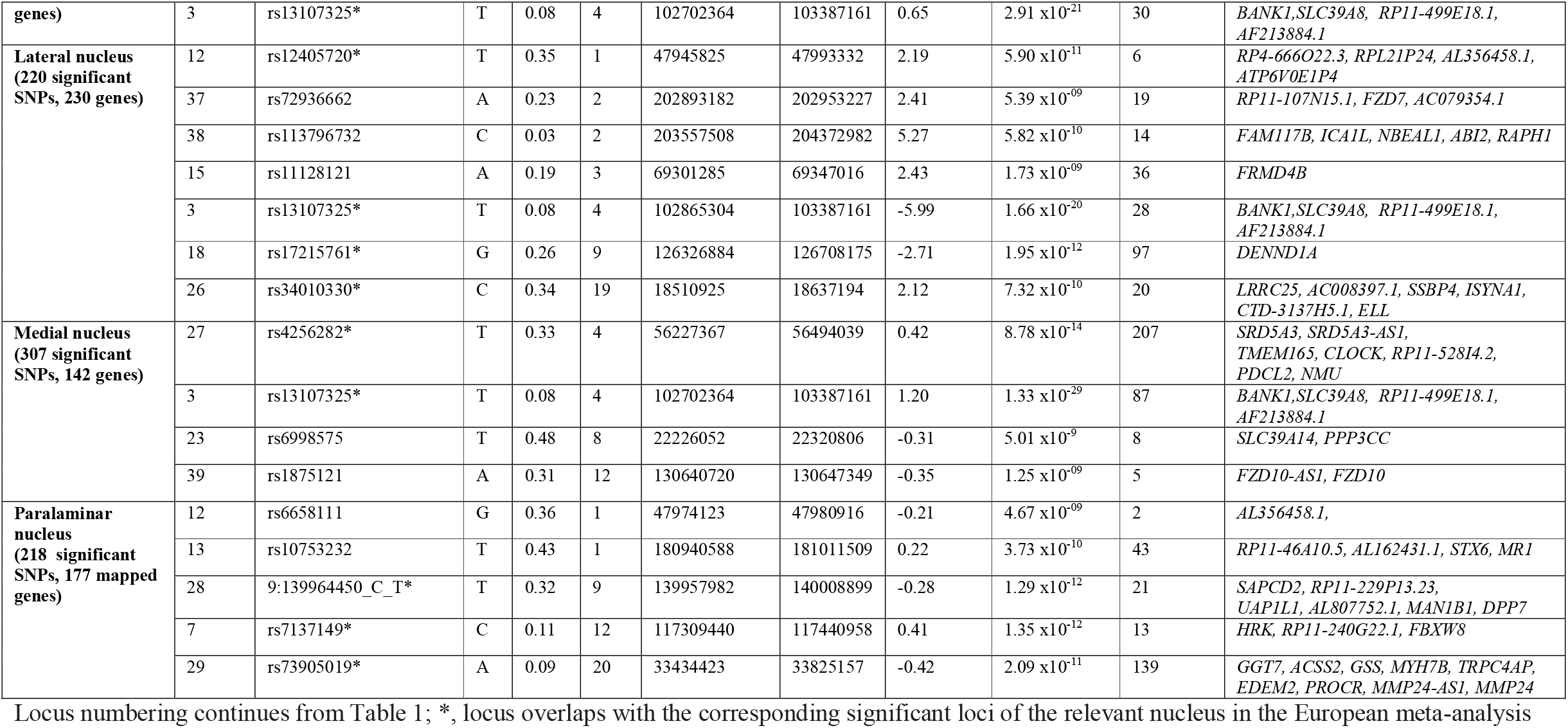
Genome-wide significant loci for whole amygdala and nuclei volumes from the trans-ancestry meta-analysis.

Figure 4 illustrates the lead SNPs’ genomic locations for each structure, highlighting that particular loci affect specific nuclei, e.g. rs12667816 affects the anterior amygdaloid area. In contrast, others have global effects, e.g. rs13131500 affects the accessory basal, anterior amygdaloid, central, cortical, lateral and medial nuclei volumes.

**Figure 4:**
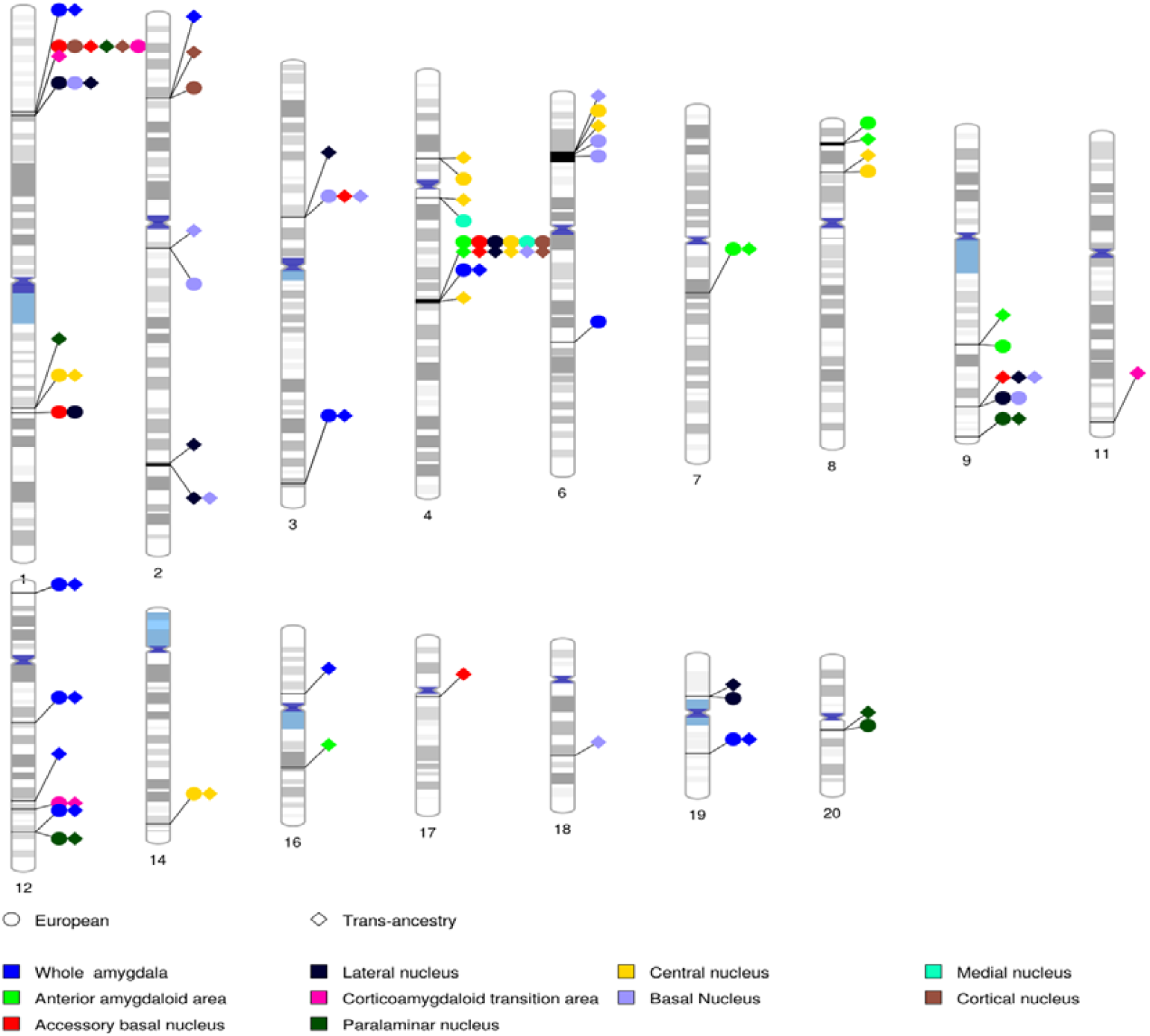
Genomic locations of genetic variants that influence amygdala nuclei volume. Each nucleus corresponds to a colour-coded circle or diamond extending to the lead SNP’s genomic location on the relevant chromosome. The European meta-analysis is denoted by a circle and the trans-ancestry analysis by a diamond. This plot was generated using *phenogram* (4)

### 3.3 Functional annotation

Across the whole amygdala and the nine nuclei, 3,630 genes were mapped to significant loci, of which 1,555 were protein-coding. Hypergeometric tests, performed as part of the *gene2func* in FUMA, identified enriched pathways for the genes mapped to all nuclei volumes except for paralaminar nucleus and corticoamygdaloid transition area volumes (Table S2). Most of the identified pathways were unique to each nucleus, except for immune-related pathways (whole amygdala volume and medial, cortical, basal, accessory basal, central and lateral nuclei volumes). The complete list of associated pathways is shown in Table T2. Further, to determine if any of the GWAS hits have been previously associated with other disorders or traits, the mapped genes were also compared against the GWAS catalogue in FUMA. Most of the genes/variants across the nuclei had been previously associated with psychiatric and behavioural traits, e.g. schizophrenia, anxiety and autism spectrum disorder (ASD), cardiovascular traits (basal, accessory basal, medial, and central nuclei volumes), and hippocampus and its subfield volumes (whole amygdala and paralaminar nucleus volumes). The complete GWAS catalogue associations are shown in Table T2.

### 3.4 Genetic overlap between nuclei, other subcortical regions and other traits

The volumetric correlations broadly mirrored the genetic correlations between each nucleus’ volume and additional subcortical regions (Figure 2). The genetic correlations further revealed two primary clusters, using Wards D2 minimum variance hierachical clustering method (Ward, 1963). The first cluster consisted of the whole amygdala, the amygdala nuclei and hippocampus volumes. The second cluster included the basal ganglia and the thalamus.

Using LD Hub, we assessed the genetic correlation between 190 traits and the whole amygdala and each of the nuclei volumes (adjusted p<2.63×10^−5^, 0.05/190*10). This was a hypothsis-free approach, including all possible traits, to allow discovery of novel associations. Significant genetic overlap was only observed between whole amygdala volume and whole hippocampus volume (r_g_=0.61, SD=0.12, p=1.85×10^−7^). The trending significant (p<0.05) associations for each nucleus is shown in Table S6.

### 3.5 Polygenicity, discoverability, power and residual inflation

The polygenicity and discoverability estimates are mainly on the same order of magnitude across the nuclei, except for the paralaminar nucleus. The paralaminar nucleus and the central nucleus volumes have the highest and lowest overall polygenicity, respectively (Table 4). The inverse relationship was observed for discoverability.

The current analyses uncovered less than 1% of the estimated variance for all of the nuclei, except for the whole amygdala (1.1% and 1.4%) and central nucleus (6% and 9%) volumes in the European meta-analysis and trans-ancestry analysis, respectively (Figure 5). Only the central nucleus volume captured more of the genetic variance than the whole amygdala volume. Gains in power were observed for the whole amygdala volume and accessory basal, basal, central, cortical and lateral nuclei in the trans-ancestry analysis compared to the European analyses. Decreased power was only observed for the anterior amygdaloid area. Using current methods, the power curve further suggests that an effective population size of 10 million samples is required to capture each nucleus’s full genetic variance.

**Figure 5:**
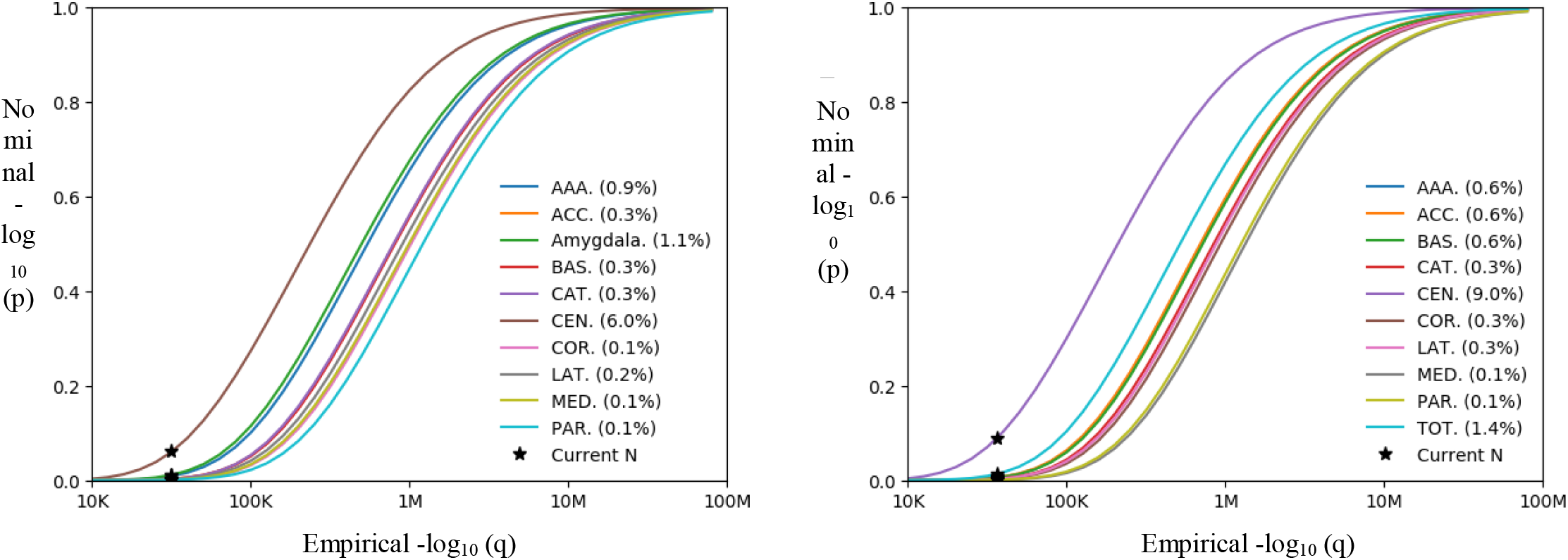
MiXeR power curves and model fit for all amygdala nuclei volumes from the European meta-analysis and trans-ancestry analysis. A and B) refer to the power curves for the whole amygdala for the European meta-analysis, respectively. This figure depicts the sample size required so that a given proportion of phenotypic variability is captured by significant SNPs for the nuclei volumes. Each curve on the plot represents a different nucleus and the right-to-left curve is determined by decreasing discoverability. The proportion of phenotypic variance explained is shown in brackets next to the corresponding nucleus volume in the legend. AAA, anterior amygdaloid area; ACC, accessory basal nucleus; BAS, basal nucleus; CAT, corticoamygdaloid transition area; CEN, central nucleus; COR, cortical nucleus; LAT, lateral nucleus; MED, medial nucleus; PAR, paralaminar nucleus; TOT, whole amygdala.

## 4 Discussion

We identified significant heritability and 39 genome-wide significant loci associated with either whole amygdala volume or with an individual amygdala nuclei volume. As evidenced by the genome-wide significant loci and genetic correlation, much of the genetic architecture is shared across the nuclei due to the high level of colinearity across their volumes, but it is also suggested that each nucleus’ volume has a unique genetic component. Our findings demonstrate that the divergent cytoarchitectures of the amygdala nuclei, forming the basis of the segmentation, are driven by both overlapping and unique genetic influences.

To the best of our knowledge, the current study is the first to investigate the SNP-based heritability of the nine amygdala nuclei volumes. SNP-based heritabilities ranged from 0.17- 0.33 across the nuclei. We also report SNP-heritability of 0.37 for the whole amygdala, which is in keeping with previous large-scale studies (Satizabal et al., 2019). Further, we show that the heritabilities vary significantly across the nuclei, with the accessory basal nucleus having approximately two-fold the heritability of the corticoamygdaloid transition area. The differences in heritabilities across the nuclei may be a consequence of neuroplasticity influenced by endogenous and exogenous sources with variable sensitivities across the nuclei (Callaghan & Tottenham, 2016).

Our findings highlight both unique and shared genetic architectures across the amygdala nuclei. The majority of the loci associated across the amygdala and the nuclei are unique to each region, and the genetic correlations across the nuclei range from – 93 to 84. Only the missense variant, rs13107325 located within *Solute Carrier Family 39 Member 8 (SLC39A8)* on chromosome four, is shared across the whole amygdala and six of the nuclei volumes. *SLC39A8* encodes *ZIP8*, a divalent metal ion transporter, highly expressed in T-cells with a significant role in innate immune function (M. J. Liu et al., 2013). The role of the immune system in the brain is discussed below. The rs13107325 variant has previously been associated with addictive behaviours (Liu et al., 2019), intelligence (Savage et al., 2018), schizophrenia (Goes et al., 2015), blood pressure (Ehret et al., 2011) and higher risk of cardiovascular death (Johansson et al., 2016). All of these traits have been associated with altered amygdala volumes (LaLumiere, 2014). Rs13107325 has further been associated with the volumes of the caudate nucleus, putamen and pallidum (Elliott et al., 2018). These basal ganglia structures have also been associated with many of the traits mentioned above, e.g. (Bernard et al., 2017). It is likely that rs13107325 has far-reaching effects across the amygdala and basal ganglia and perhaps the connections between these structures, which in turn influence certain disorders. Disorders associated with disrupted functional connectivity between the amygdala and basal ganglia have been reported, for example, reduced functional connectivity between these regions has been associated with tremor-dominant Parkinson’s disease (Guan et al., 2017).

In the whole amygdala volume GWAS, 83% of the significant variants had a p<0.05 and the same direction of effect in the previous GWAS of amygdala volume (Satizabal et al., 2019). We also replicated the previously reported genome-wide significant locus associated with amygdala volume (rs17178006 on chromosome 12) (Satizabal et al., 2019). This locus spans an intronic region associated with several psychiatric phenotypes. For example, cognitive performance (Cirulli et al., 2010) and bipolar disorder (Smith et al., 2009). The previous amygdala volume GWAS (Satizabal et al., 2019) had approximately the same number of study participants as the present study; however, we have identified many more variants. First, this discrepancy may be due to the nature of mega-versus meta-analyses. It has been shown that only under strict conditions do these two methods have equal power (Lin & Zeng, 2010). Second, the Satzibal et al. study comprised data from three consortia, with European participants from across the globe and at variable age ranges (9 - 90 years). The UKB samples are sourced from the same geographical location and only include adult participants (44 – 82 years) (Bycroft et al., 2018; Miller et al., 2016). The inclusion of a limited age range and European participants from the same country significantly reduce sample heterogeneity, allowing for improved power to detect associated variants.

The traits associated with the genes mapped to the nuclei volumes revealed associations that were not identified when considering the whole amygdala and refined trait associations with the whole amygdala to specific nuclei. For example, many of the genes mapped to the basal, accessory basal, medial, and central nuclei are associated with cardiovascular system traits, and traits related to anxiety and neuroticism were enriched across the genes mapped to the anterior amygdaloid area and basal nucleus volumes. These associations were not observed when considering the loci for the whole amygdala volume. The traits associated with the genes mapped to the nuclei volumes may also refine trait associations with the whole amygdala to specific nuclei. For example, schizophrenia was associated with the genes mapped to the whole amygdala volume and the anterior-amygdaloid-area, basal, cortical, and central nuclei volumes in our study. The specific roles of these nuclei in schizophrenia pathophysiology, therefore, warrant further investigation. The genetic architecture of the nuclei has the potential to reveal novel and refine pre-existing trait associations with the whole amygdala, which may inform the pathophysiology of these traits.

Immune-related pathways were particualrly associated with the genes mapped across several nuclei. The brain has a resident immune system that interacts with peripheral immunity and impacts behaviour (Bennett & Molofsky, 2019). Many of the trait associations identified across the nuclei, e.g. schizophrenia (Sekar et al., 2016), ASD (Onore et al., 2012) and anxiety and mood disorders (Passos et al., 2015; Zorrilla et al., 2001), have immune components in their aetiology. Animal models have demonstrated that maternal immune activation during pregnancy significantly affects brain development and is a risk factor for many neurological disorders (Knuesel et al., 2014). The resident immune and immune-related cells in the brain affect synaptic and myelin formation and pruning throughout the lifespan, affecting brain volume and communication (Bennett & Molofsky, 2019). One such resident group of immune cells, the microglia, may directly influence behaviour. For example, studies in mice have shown an increased risk for obsessive-compulsive symptoms when microglia are hyperactivated (Krabbe et al., 2017) and ASD-like symptoms when microglial signalling is impaired (Zhan et al., 2014). Our findings suggest that these immune pathways may be pertinent to amygdala nuclei structure and function.

When investigating the polygenicity and discoverability of our dataset, we showed 1) minimal inflation of z-scores due to cryptic relatedness, 2) the central nucleus had the greatest statistical power, 3) polygenicity and discoverability vary across the nuclei, and 4) significantly larger sample sizes are needed to elucidate the genetic architecture of each nucleus fully. The increased power of the central nucleus, as compared to the whole amygdala, supports the need to study the nuclei separately in future research. A possible explanation for the incread power of the central nucleus may be that this nucleus has more evolutionarily conserved genetics that are linked to its role in fear expression.

Some limitations should be mentioned. First, most of our participants were of European ancestry, with only 3% of the trans-ancestry analysis consisting of non-European participants. However, with the addition of these diverse population groups, we identified ten additional candidate loci, added support for 25 loci identified in the European-only analysis and suggested that the four variants that were not replicated may be population specific or false positives. This highlights the need to improve the representation of other ethnicities, which will likely drive further discovery, and which can be used in fine-mapping to determine causation and population specificity. Second, specialized validation of the segmentation and consideration of a genetic expression atlas (e.g. as done for the hippocampus (Thompson et al., 2008)) may aid in further noise reduction and amplify power. Harmonizing the definitions and segmentation algorithms used in the literature will significantly improve discoverability (Fan et al., 2018). Third, as demonstrated with MiXeR, larger sample sizes are needed to fully elucidate each nucleus’ genetic architecture. Fourth, trait associations between the mapped genes across the nuclei should be validated with global measures of genetic concordance or overlap using approaches such as LDSC (Bulik-Sullivan et al., 2015) and Conjunctional False Discovery Rate Analysis (Andreassen et al., 2013). Due to the polygenic nature of the traits discussed here, such approaches are necessary to ensure significant correlation before conclusions can be made about the involvement of specific nuclei in these traits.

Overall, we show that investigating the amygdala with increased power and phenotypic specificity through segmentation of the nuclei aids genetic discoverability. We have shown significant heritability and identified 39 novel variants associated with the amygdala and its nuclei. Our findings indicate that the amygdala nuclei are mainly genetically distinct and have unique associations with biological processes and genetic link with brain disorders and cardiovascular traits. We further highlight the role of immune-related pathways across several nuclei. Continued efforts are needed to further our understanding of the genes implicated here to fully elucidate the amygdala nuclei’s functions.

## Data Availability

All summary statistics are available from https://github.com/norment/open-science

## 5 Downloads

All summary statistics are available from https://github.com/norment/open-science

## 6 Acknowledgements

This work was partly performed on the TSD (Tjeneste for Sensitive Data) facilities, owned by the University of Oslo, operated and developed by the TSD service group at the University of Oslo, IT-Department (USIT). Dr. Morey was supported by the US Department of Veterans Affairs (VA) Mid-Atlantic Mental Illness Research, Education, and Clinical Center (MIRECC) core funds of the Department of Veterans Affairs Office of Mental Health Services. Dr. Morey also received financial support from the VA Office of Research and Development (5I01CX000748-01, 5I01CX000120-02). Additional financial support was provided by the National Institute for Neurological Disorders and Stroke (R01NS086885-01A1). MM is supported by the South African National Research Fund, the David and Elaine Potter Foundation and the GINGER program. The GINGER program is, in part, supported by an award from the National Institute for Mental Health (1R01MH120642). TK, DM and OA are funded by the Research Council of Norway (#276082, #223273). RR, DS and SD are supported by the South African Medical Research Council.

