## Supplemental Figures S1-10 and tables S1-5 for "The Genetic Architecture of Amygdala Nuclei"

**Supplementary materials for “The Genetic Architecture of Amygdala Nuclei.”**

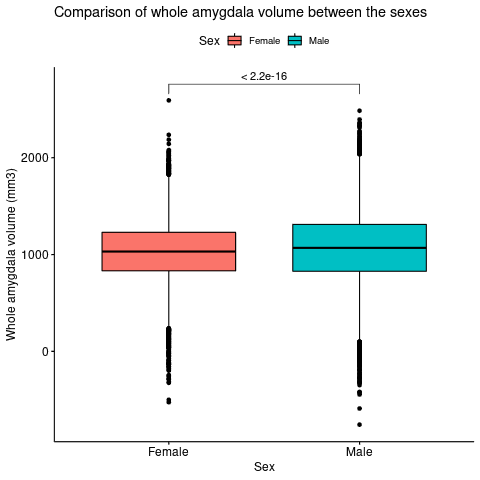

**Figure S1: Comparison of whole amygdala volume between males and females**

*The y-axis shows amygdala volume in cubic millimeter and the x-axis displays the sex of the two groups being compared. Whole amygdala volume has been adjusted for intracranial volume.*

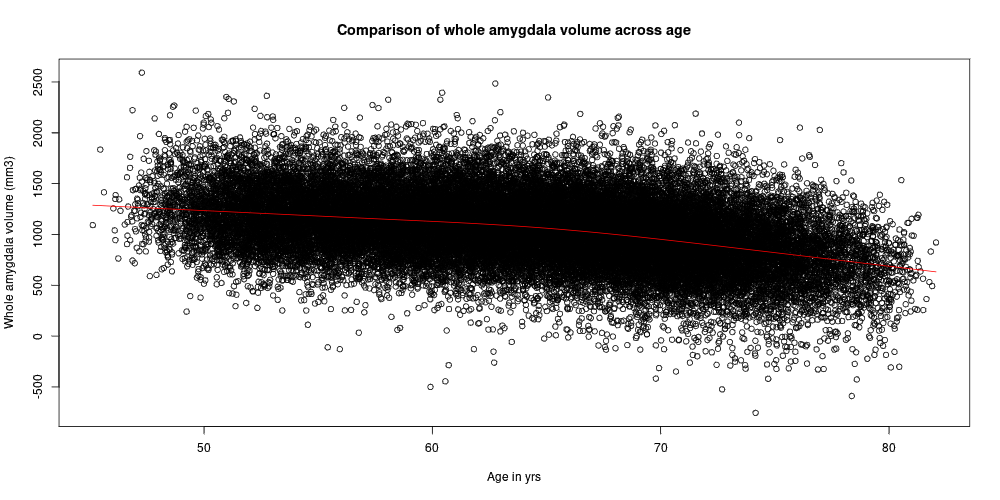
**Figure S2: Comparison of amygdala volume across** **age**

*The y-axis of the scatter plot shows the* *shows amygdala volume in cubic millimeter and the x-axis displays the age range in years. Whole amygdala volume has been adjusted for intracranial volume. There is a trend towards lower amygdala volume with increasing age.*

**Figure S3: Distribution of amygdala nuclei volumes**

| Amygdala nuclei | Nuclei volume (mm3) |
| --- | --- |
| Basal nucleus | 1423.2 |
| Lateral nucleus | 1327.58 |
| Accessory basal nucleus | 527.68 |
| Corticoamygdaloid transition area | 358.671 |
| Anterior amygdaloid area | 117.972 |
| Paralaminar nucleus | 101.519 |
| Central nucleus | 90.3791 |
| Cortical nucleus | 51.0127 |
| Medial Nucleus | 43.1244 |

*The y-axis shows the volume in cubic millimeter per nucleus, indicated on the x-axis. The blue bars represent the mean volume per nucleus across study participants and the orange error bars reflect the standard deviation of this mean.*

**Table S1: The number of participants and SNPs remaining after quality control**

|  | European | | Trans-ancestry* | |
| --- | --- | --- | --- | --- |
| Filtering steps | # Subjects | #SNPs | #Subjects | #SNPs |
| START: Version 3 imputed bgen files provided by UKBB** | 487,411 | 89,387,505 | 487,411 | 89,387,505 |
| Remove participants without MRI data | 42,067 | 89,387,505 | 42,067 | 89,387,505 |
| Remove participants with missing covariates | 41,035 | 89,387,505 | 41,035 | 9,915,367 |
| Remove duplicate participants | 41,035 | 89,387,505 | 41,035 | 9,915,367 |
| Remove unused factor levels | 41,035 | 89,387,505 | 41,035 | 9,915,367 |
| Keep validated Europeans only | 35,660 | 89,387,505 | N/A | N/A |
| Genotyping filters:  SNPs only, poorly imputed SNPs (r^2^<0.5), low minor allele frequency (<0.1%), genotyping rate (<0.1), Hardy-Weinberg Equilibrium (p<1x10^-9^), duplicate markers | 35,660 | 12,245,112 | 40,964 | 9,915,367 |
| Remove participants with missing Euler numbers | 35,657 | 12,245,112 | 40,961 | 9,915,367 |
| Remove participants with Euler number outliers (4SD) | 35,186 | 12,245,112 | 40,540 | 9,915,367 |
| Remove participants with a disorder affecting the brain | 32,215 | 12,245,112 | 36,798 | 9,915,367 |
| Remove participants for which the amygdala nuclei segmentation failed | 31,971 | 12,245,112 | 36,798 | 9,915,367 |
| Remove participants with amygdala nuclei volume outliers for each hemisphere (4SD) | 31,714 | 12,245,112 | 36,531 | 9,915,367 |
| Remove participants with ICV outliers (4SD) | 31,690 | 12,245,112 | 36,512 | 9,915,367 |
| Remove participants without ancestry | N/A | N/A | 36,352 | 9,915,367 |
| Total | 31,690 | 12,245,112 | 36,352 | 9,915,367 |

** The trans-ancestry analysis includes the 31,690 participants of European decent and an additional 4,662 participants from other ancestries; ** Participants who had withdrawn consent by March 2020 were also excluded; SNPs, single nucleotide polymorphisms; UKBB, UK-Biobank; SD, standard deviation; ICV, intercranial volume*

**Table S2: Heritability estimates and genetic correlations of both hemispheres of the amygdala nuclei volumes form the European meta-analysis as determined with LDSC**

| Region of interest | | Heritability | | | Genetic correlation with right hemisphere | | |
| --- | --- | --- | --- | --- | --- | --- | --- |
|  |  | h2 | SE | Z score | r_g_ | SE | p-value |
| Left hemisphere | Whole amygdala | 0.24 | 0.02 | 12.00 | 0.96 | 0.02 | 1.46 x10^-43^ |
|  | Anterior amygdaloid area | 0.22 | 0.02 | 11.00 | 0.57 | 0.02 | 8.64 x10^-23^ |
|  | Accessory basal nucleus | 0.14 | 0.02 | 7.00 | 0.96 | 0.04 | 1.65 x10^-110^ |
|  | Basal nucleus | 0.12 | 0.02 | 6.00 | 0,96 | 0.07 | 1.89 x10^-42^ |
|  | Corticoamygdaloid transition area | 0.09 | 0.02 | 4.50 | 0.64 | 0.09 | 5.28 x10^-11^ |
|  | Cortical nucleus | 0.11 | 0.02 | 5.50 | 0.88 | 0.06 | 6.84 x10^-44^ |
|  | Central nucleus | 0.11 | 0.02 | 5.50 | 0.99 | 0.07 | 1.46 x10^-43^ |
|  | Lateral nucleus | 0.14 | 0.02 | 7.00 | 0.92 | 0.05 | 16.91 x10^-86^ |
|  | Medial nucleus | 0.10 | 0.02 | 5.00 | 0.82 | 0.08 | 3.16 x10^-34^ |
|  | Paralaminar nucleus | 0.18 | 0.02 | 9.00 | 0.97 | 0.05 | 1.02 x10^-76^ |
| Right hemisphere | Whole amygdala | 0.25 | 0.02 | 12.00 |  |  | |
|  | Anterior amygdaloid area | 0.16 | 0.03 | 5.33 |  |  |  |
|  | Accessory basal nucleus | 0.20 | 0.02 | 10.00 |  |  |  |
|  | Basal nucleus | 0.13 | 0.02 | 76.50 |  |  |  |
|  | Corticoamygdaloid transition area | 0.10 | 0.02 | 5.00 |  |  |  |
|  | Cortical nucleus | 0.15 | 0.02 | 7.50 |  |  |  |
|  | Central nucleus | 0.11 | 0.02 | 5.50 |  |  |  |
|  | Lateral nucleus | 0.21 | 0.02 | 10.50 |  |  |  |
|  | Medial nucleus | 0.11 | 0.02 | 5.50 |  |  |  |
|  | Paralaminar nucleus | 0.16 | 0.02 | 8.00 |  |  |  |

*h^2^, heritability; SE, standard error; rg, regression coefficient*

**Figure S4. Manhattan and QQ plots per amygdala nucleus volume uncorrected for whole amygdala volume for the European meta-analysis.**

*For the Manhattan plots, the red line indicates the adjusted whole-genome significance threshold (5x10^-9^ p-value) and the blue line the standard genome-wide significance threshold (1x10^-8^).*

*Anterior amygdaloid area*

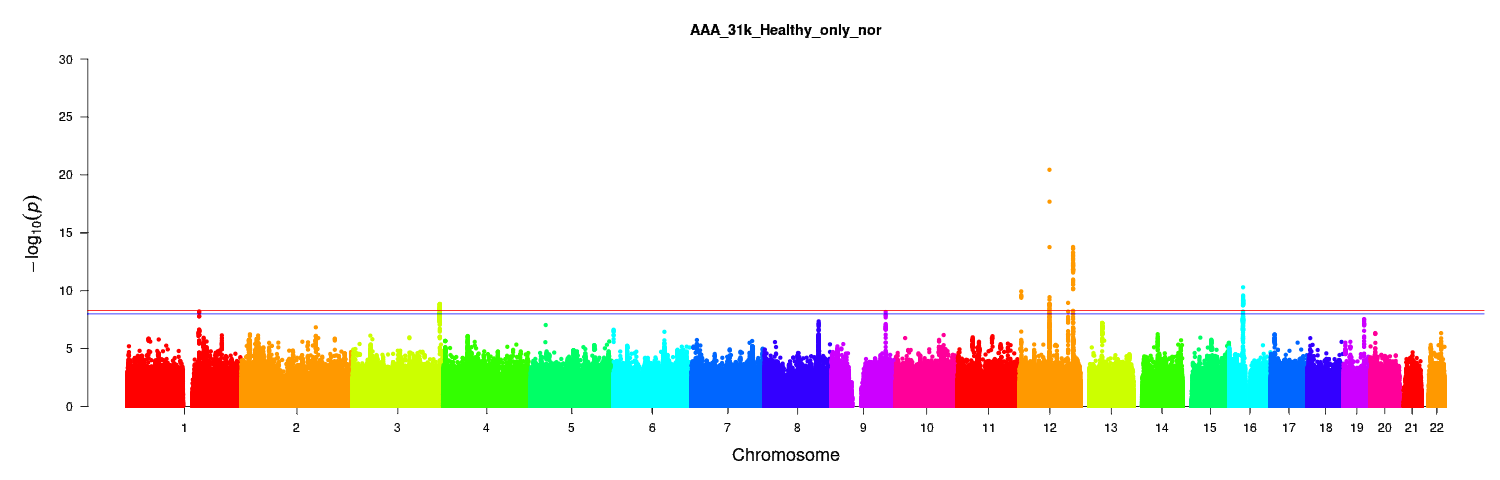

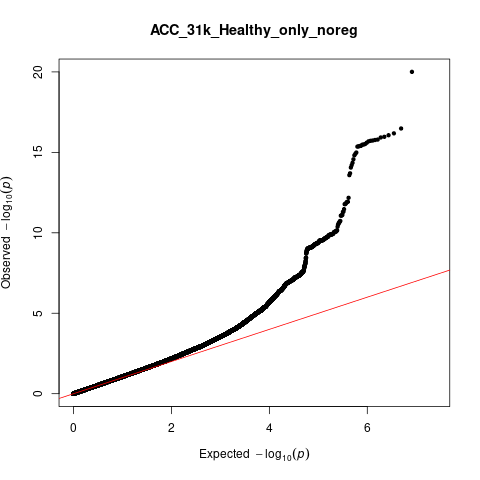

λ_GC_=1.07

Accessory basal nucleus

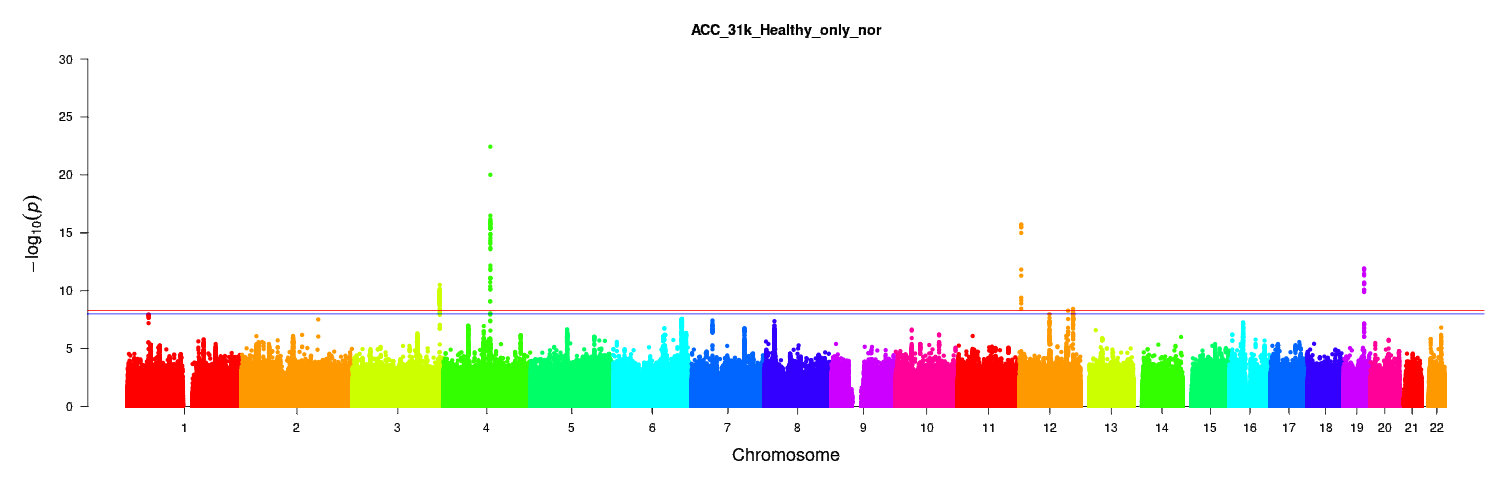

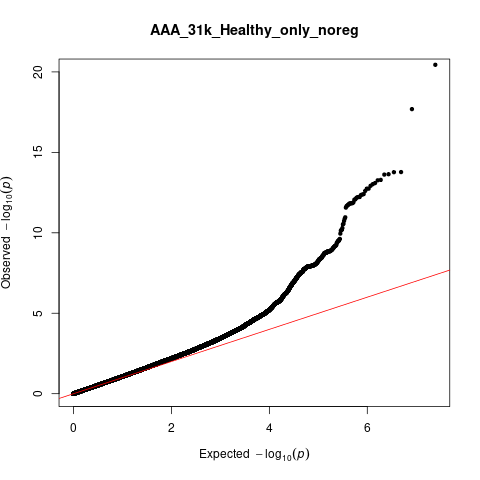

λ_GC_=1.08

Basal nucleus

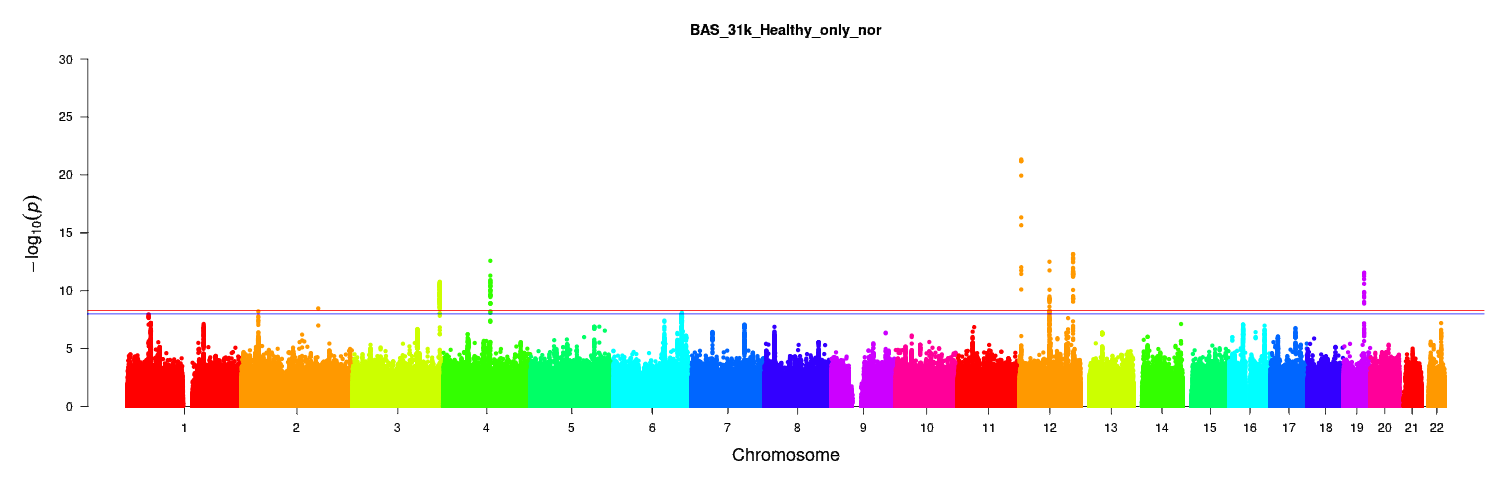

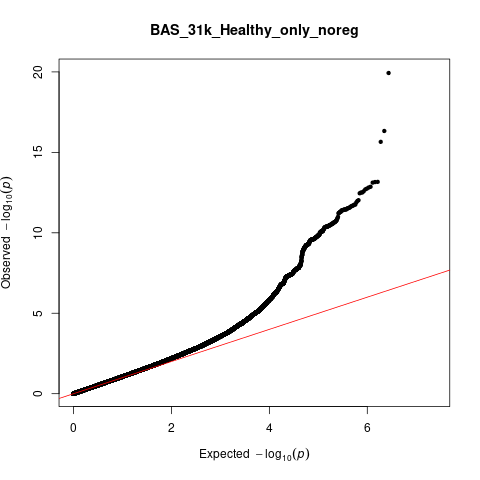

λ_GC_=1.08

Corticoamygdaloid transition area

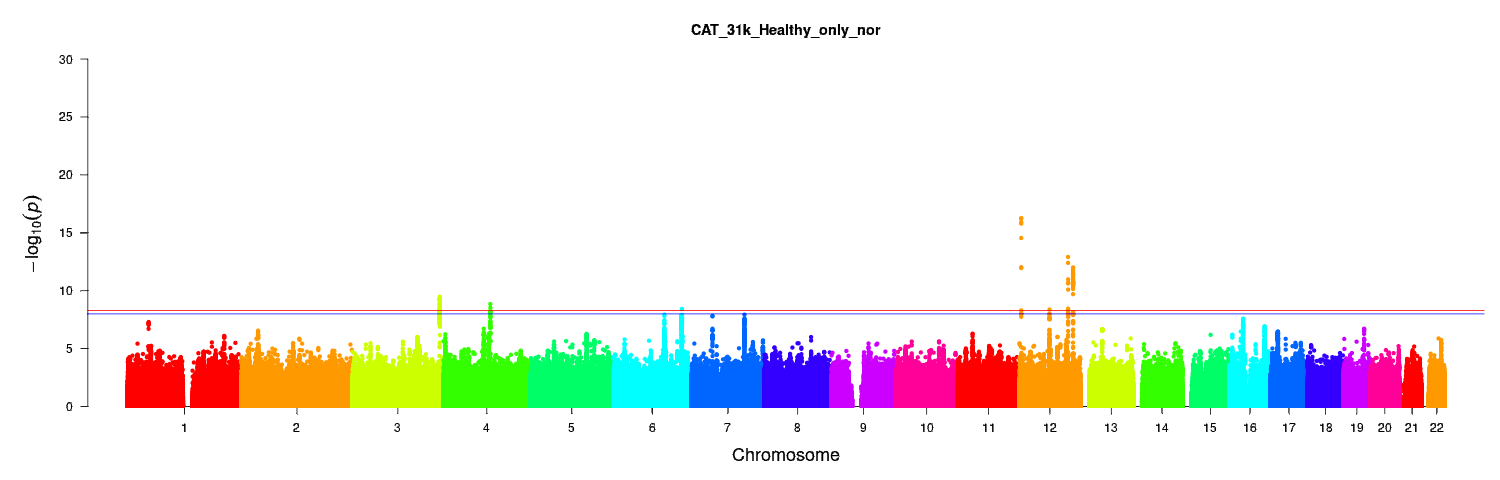

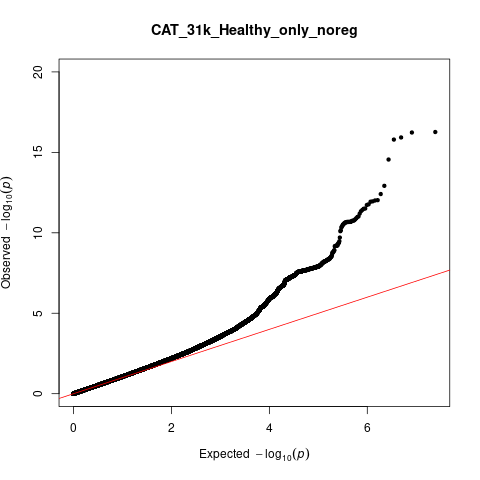

λ_GC_=1.07

Central nucleus

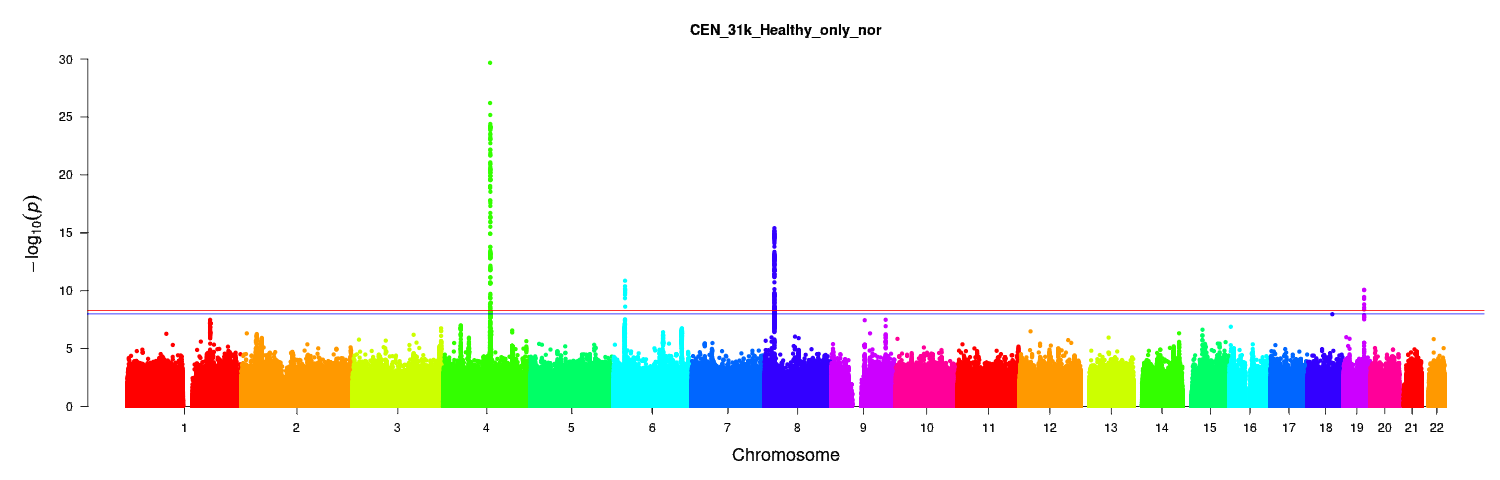

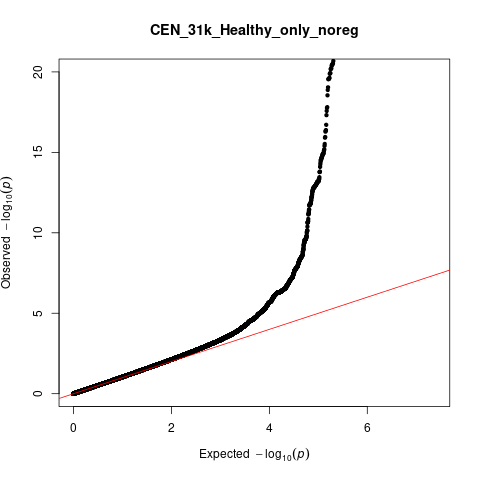

λ_GC_=1.05

Cortical nucleus

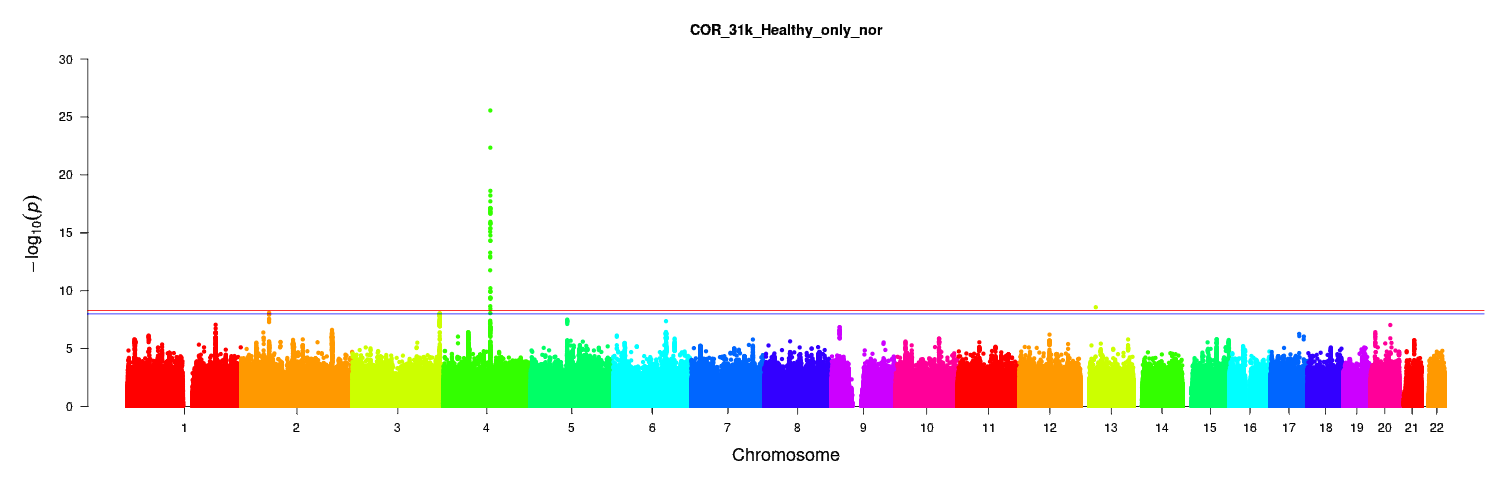

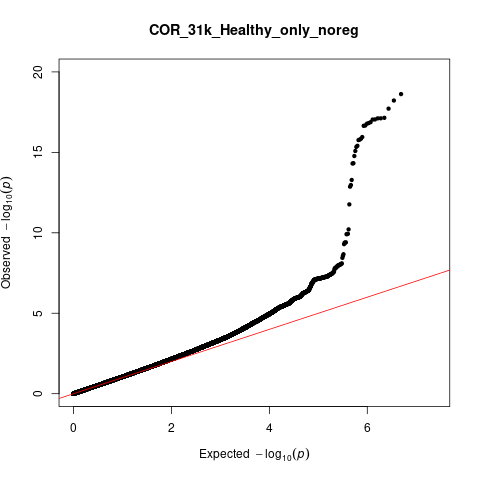

λ_GC_=1.06

Lateral nucleus

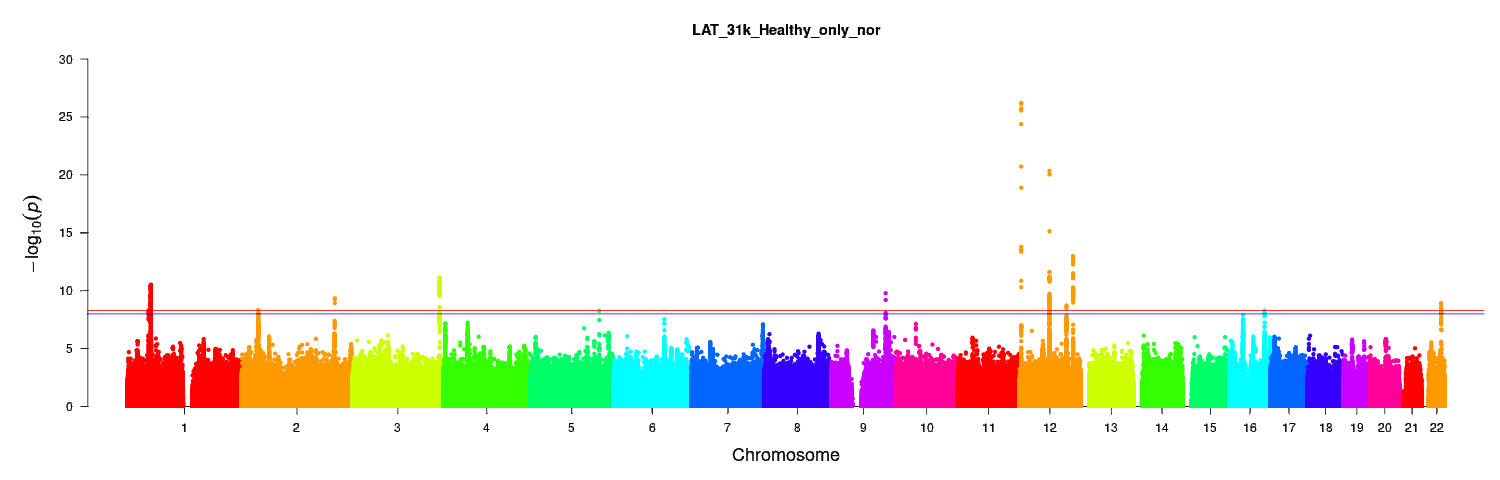

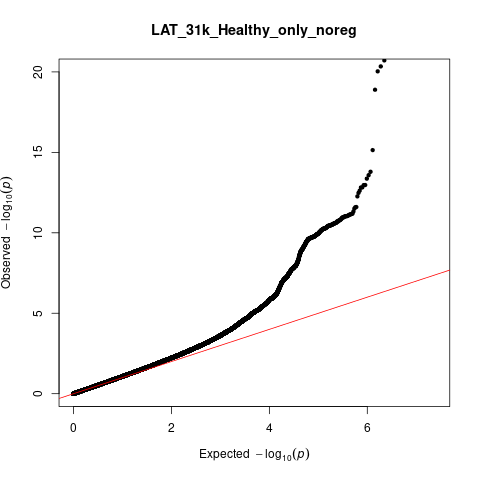

λ_GC_=1.09

Medial nucleus

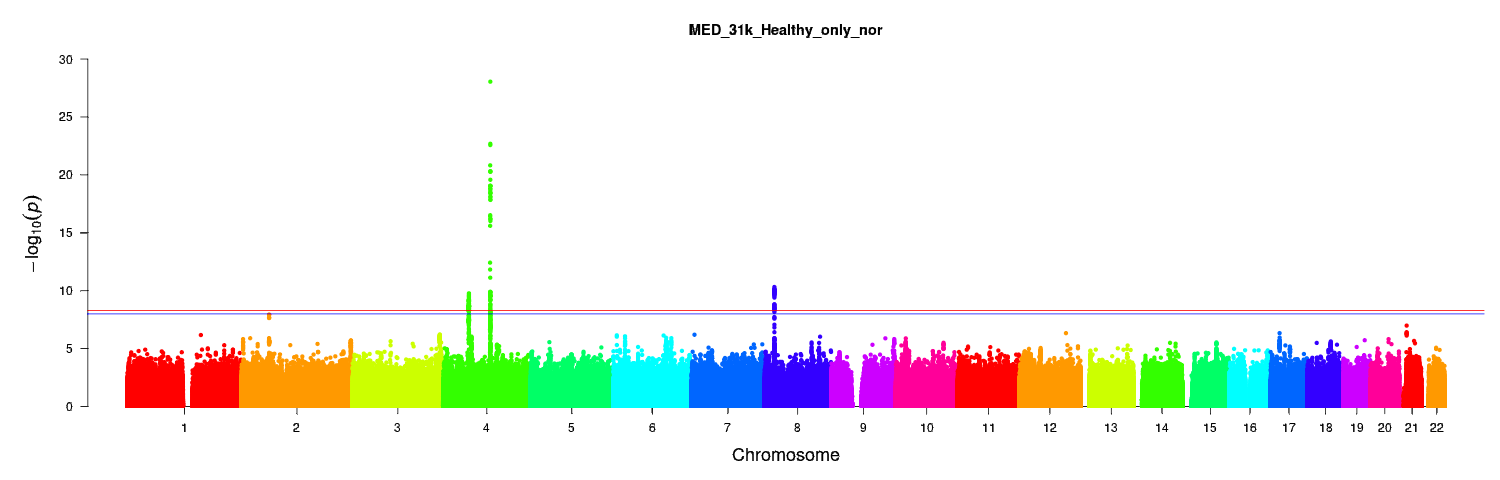

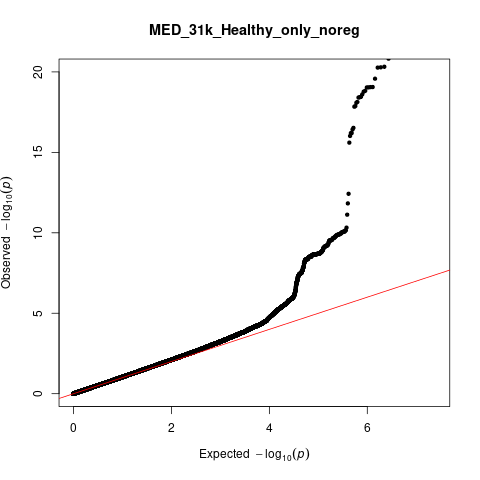

λ_GC_=1.05

Paralaminar nucleus

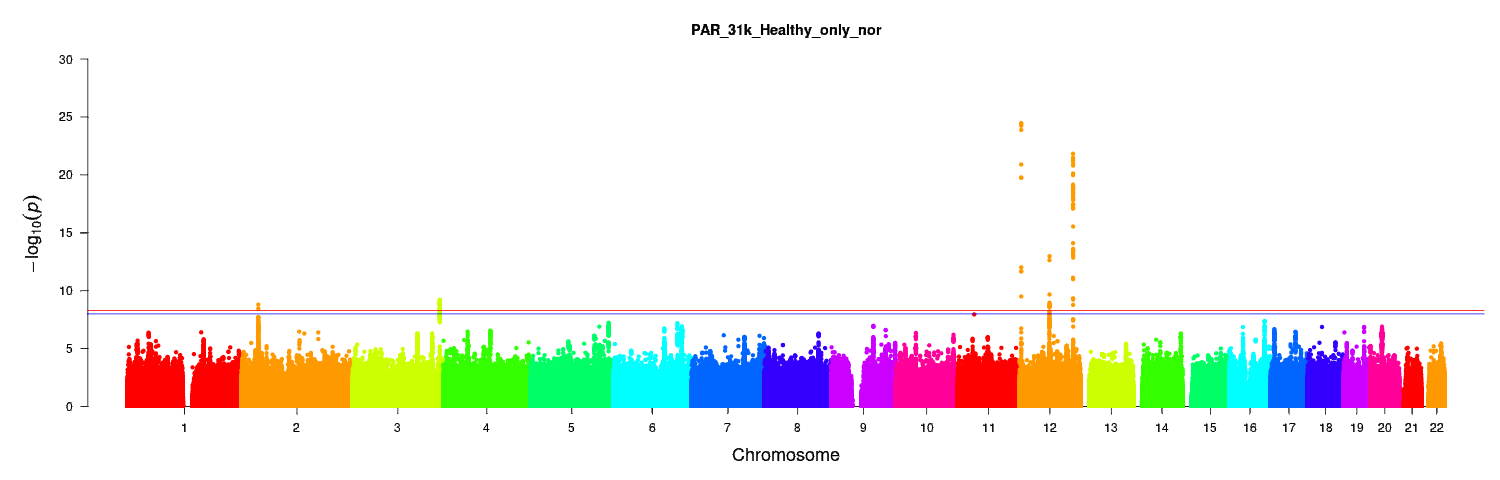

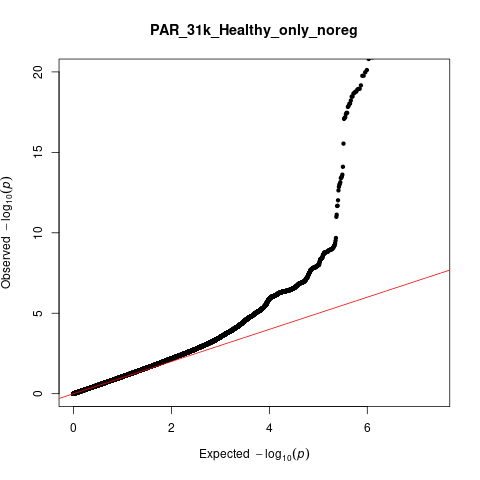

λ_GC_=1.08

**Table S3: Whole-genome significant loci for the nuclei without covarying for total amygdala volume in the European meta-analysis**

| **Structure** | **Number of unique loci** | **Lead SNP** | **A1** | **MAF** | **Chr** | **Start (BP)** | **End (BP)** | **Beta** | **P-value** | **Number of significant SNPs** | **Nearest gene** |
| --- | --- | --- | --- | --- | --- | --- | --- | --- | --- | --- | --- |
| **Anterior amygdaloid area**  **(456 significant SNPs, 310 genes)** | 1 | rs4308325 | C | 0.39 | 3 | 190591911 | 190678743 | 0.68 | 1.37 x10^-09^ | 159 | *GMNC* |
|  | 2 | rs1419859 | T | 0.37 | 12 | 4005023 | 4013260 | -0.73 | 1.12x10^-10^ | 7 | *RP11-664D1.1* |
|  | 3 | rs17178006 | G | 0.08 | 12 | 65448568 | 65905126 | -1.60 | 3.61x10^-21^ | 130 | *MSRB3, RP11-230G5.2* |
|  | 4 | rs17038091 | C | 0.18 | 12 | 106443251 | 106502283 | -0.85 | 1.11x10^-09^ | 16 | *RP11-114F10.2, NUAK1* |
|  | 5 | rs11068224 | A | 0.08 | 12 | 117309440 | 117516922 | 1.55 | 1.68x10^-14^ | 45 | *HRK, RP11-240G22.1* |
|  | 6 | rs8054179 | T | 0.43 | 16 | 29986627 | 30438138 | -0.74 | 5.06x10^-11^ | 99 | *MAPK3, CORO1A, RP11-347C12.2, CD2BP2, TBC1D10B, MYLPF, ZNF48, SEPT1, ZNF771, DCTPP1* |
| **Accessory basal nucleus**  **(355 significant SNPs, 186 mapped genes)** | 1 | rs113591830 | A | 0.08 | 3 | 190591418 | 190678743 | -5.27 | 3.14x10^-11^ | 221 | *GMNC* |
|  | 7 | rs13107325 | T | 0.07 | 4 | 102865304 | 103388441 | 8.19 | 3.69x10^-23^ | 44 | *BANK1, RP11-498M5.2, SLC39A8* |
|  | 8 | rs11062908 | G | 0.39 | 12 | 4004752 | 4013260 | -3.59 | 1.91x10^-16^ | 11 | *RP11-664D1.1* |
|  | 4 | rs17038091 | C | 0.18 | 12 | 106443251 | 106502283 | -3.17 | 5.32x10^-09^ | 16 | *RP11-114F10.2, NUAK1* |
|  | 5 | rs11068224 | A | 0.08 | 12 | 117309440 | 117485833 | 4.60 | 3.66x10^-09^ | 38 | *HRK, RP11-240G22.1, FBXW8, TESC* |
|  | 9 | rs429358 | C | 0.15 | 19 | 45388500 | 45428234 | -4.18 | 1.19x10^-12^ | 25 | *PVRL2, CTB-129P6.4, TOMM40, APOE, APOC1* |
| **Basal Nucleus**  **(475 significant SNPs, 246 mapped genes)** | 10 | rs76208066 | A | 0.01 | 2 | 168094504 | 168197895 | 57.66 | 3.36x10^-09^ | 2 | *XIRP2, AC074363.1* |
|  | 1 | rs58531798 | G | 0.4 | 3 | 190591418 | 190678743 | 7.31 | 8.24x10^-11^ | 227 | *GMNC* |
|  | 7 | rs13107325 | T | 0.08 | 4 | 102865304 | 103388441 | 15.01 | 2.62x10^-13^ | 44 | *BANK1, RP11-498M5.2, SLC39A8* |
|  | 8 | rs1419859 | T | 0.38 | 12 | 4004752 | 4013260 | -10.52 | 4.65x10^-22^ | 11 | *RP11-664D1.1* |
|  | 3 | rs17178006 | G | 0.08 | 12 | 65448568 | 65874956 | -11.80 | 3.14x10^-13^ | 127 | *MSRB3, RP11-230G5.2* |
|  | 5 | rs77956314 | C | 0.09 | 12 | 117309440 | 117516922 | 14.38 | 6.85x10^-14^ | 43 | *HRK,*  *RP11-240G22.1* |
|  | 9 | rs10414043 | A | 0.12 | 19 | 45386467 | 45428234 | -11.09 | 2.75x10^-12^ | 21 | *PVRL2, CTB-129P6.4, TOMM40,*  *APOE, APOC1* |
| **Corticoamygdaloid transition area**  **(775 significant SNPs, 286 mapped genes)** | 1 | rs34113929 | A | 0.45 | 3 | 190591418 | 190678743 | -0.92 | 3.32x10^-10^ | 221 | *GMNC* |
|  | 7 | rs13107325 | T | 0.08 | 4 | 102865304 | 103388441 | 3.41 | 1.42x10^-09^ | 30 | *BANK1, RP11-498M5.2, SLC39A8* |
|  | 11 | rs14314 | C | 0.37 | 6 | 149874899 | 150244689 | 1.80 | 3.69x10^-09^ | 454 | *RP1-12G14.5, GINM1, RP1-12G14.7,*  *RPS18P9, KATNA1,*  *LATS1, NUP43, PCMT1, RP11-350J20.4, LRP11, RP11-244K5.8, CCT7P1, RAET1E* |
|  | 8 | rs11062908 | G | 0.4 | 12 | 4004752 | 4013260 | -2.50 | 5.44 x10^-17^ | 11 | *RP11-664D1.1* |
|  | 3 | rs73123652 | C | 0.08 | 12 | 65718299 | 65874956 | -2.73 | 4.21 x10^-09^ | 3 | *MSRB3, RP11-230G5.2* |
|  | 4 | rs12146713 | C | 0.09 | 12 | 106443251 | 106510413 | -3.63 | 1.21 x10^-13^ | 18 | *NUAK1* |
|  | 5 | rs77956314 | C | 0.09 | 12 | 117309440 | 117477082 | 3.76 | 9.79 x10^-13^ | 38 | *HRK, RP11-240G22.1, FBXW8* |
| **Central nucleus**  **(583 significant SNPs, 313 genes)** | 7 | rs13107325 | T | 0.07 | 4 | 102637936 | 103388441 | 3.66 | 2.61 x10^-81^ | 307 | *BANK1, RP11-498M5.2, SLC39A8* |
|  | 12 | rs114601774 | C | 0.01 | 4 | 103651441 | 104780790 | 3.26 | 1.07 x10^-09^ | 4 | *MANBA, LRRC37A15P, CENPE, RP11-328K4.1, RNU6-635P* |
|  | 13 | rs80215559 | C | 0.04 | 6 | 25578433 | 26123502 | -1.25 | 1.37 x10^-11^ | 14 | *LRRC16A, HIST1H2AA, SLC17A1, SLC17A2, SLC17A3* |
|  | 14 | rs7001171 | G | 0.38 | 8 | 22226027 | 22408684 | -0.83 | 3.97 x10^-16^ | 239 | *SLC39A14, RNU6-336P, PPP3CC, RP11-582J16.4, SORBS3* |
|  | 9 | rs10414043 | A | 0.12 | 19 | 45386467 | 45428234 | -0.96 | 8.32 x10^-11^ | 19 | *PVRL2, CTB-129P6.4, TOMM40, APOE, APOC1, APOC1P1* |
| **Cortical nucleus**  **(52 significant SNPs, 78 mapped genes)** | 7 | rs13107325 | T | 0.08 | 4 | 102735510 | 103387161 | 1.08 | 2.76 x10^-26^ | 49 | *BANK1, RP11-498M5.2, SLC39A8* |
|  | 15 | rs148882579 | A | 0.01 | 13 | 32432080 | 34136094 | 2.89 | 2.65 x10^-09^ | 3 | *EEF1DP3, FRY, N4BP2L2,*  *PDS5B, LINC00423, KL, STARD13, RP11-141M1.3* |
| **Lateral nucleus**  **(903 significant SNPs, 503 genes)** | 16 | rs33931638 | A | 0.07 | 1 | 45893811 | 46587530 | -10.22 | 5.08 x10^-09^ | 10 | *TESK2, CCDC163P, MAST2, PIK3R3* |
|  | 17 | rs3176459 | G | 0.33 | 1 | 50828347 | 51742123 | -6.54 | 3.01 x10^-11^ | 186 | *HMGB1P45, FCF1P6, DMRTA2,*  *RP5-850O15.4, FAF1, CDKN2C, MIR4421* |
|  | 18 | rs6730884 | T | 0.41 | 2 | 37047901 | 37134092 | 5.48 | 4.70 x10^-09^ | 18 | *VIT, AC007382.1, STRN* |
|  | 19 | rs111737209 | T | 0.04 | 2 | 203557508 | 204372982 | 14.70 | 4.47 x10^-10^ | 14 | *FAM117B,*  *ICA1L, NBEAL1, ABI2, RAPH1* |
|  | 1 | rs61494018 | G | 0.39 | 3 | 190591911 | 190678743 | 6.58 | 7.03 x10^-12^ | 159 | *GMNC* |
|  | 20 | rs3792789 | G | 0.36 | 4 | 150434422 | 150445968 | -5.63 | 5.51 x10^-09^ | 6 | *TNIP1* |
|  | 21 | rs6478241 | A | 0.38 | 9 | 119241165 | 119266468 | -6.12 | 1.64 x10^-10^ | 17 | *ASTN2* |
|  | 8 | rs1419859 | T | 0.37 | 12 | 4004752 | 4068627 | -10.32 | 6.28 x10^-27^ | 12 | *RP11-664D1.1* |
|  | 3 | rs17178006 | G | 0.08 | 12 | 65449071 | 65874956 | -13.44 | 4.55 x10^-21^ | 121 | *MSRB3:RP11-230G5.2* |
|  | 22 | rs2195243 | C | 0.18 | 12 | 102394872 | 102942969 | -6.66 | 1.87 x10^-9^ | 159 | *DRAM1, CCDC53, RP11-554E23.4, NUP37,*  *PARPBP, PMCH, RN7SL793P, RP11-18O15.1, IGF1, RP11-210L7.1* |
|  | 5 | rs11068224 | A | 0.08 | 12 | 117309440 | 117516922 | 12.69 | 1.07 x10^-13^ | 43 | *HRK, RP11-240G22.1* |
|  | 23 | rs9927552 | G | 0.34 | 16 | 77290099 | 77320605 | -5.61 | 5.29 x10^-09^ | 8 | *RP11-538I12.3, ADAMTS18* |
|  | 24 | rs13053653 | T | 0.27 | 22 | 43612426 | 43626035 | 6.33 | 1.18 x10^-09^ | 14 | *SCUBE1* |
| **Medial nucleus (372 significant SNPs, 127 mapped genes)** | 25 | rs4256282 | T | 0.33 | 4 | 56227367 | 56477181 | 0.44 | 1.67 x10^-10^ | 210 | *SRD5A3, SRD5A3-AS1, TMEM165, CLOCK,*  *PDCL2, RP11-528I4.2, NMU* |
|  | 7 | rs13107325 | T | 0.07 | 4 | 102702364 | 103387161 | 1.56 | 1.13 x10^-33^ | 88 | *SLC39A8, BANK1, RP11-498M5.2* |
|  | 14 | rs7816557 | G | 0.49 | 8 | 22226052 | 22320806 | -0.43 | 4.77 x10^-11^ | 74 | *SLC39A14, CTD-2036J7.1, PPP3CC* |
| **Paralaminar nucleus**  **(382 significant SNPs, 182 mapped genes)** | 18 | rs12478822 | C | 0.42 | 2 | 37047901 | 37056330 | 0.46 | 1.59 x10^-09^ | 21 | *VIT, AC007382.1* |
|  | 1 | rs10937450 | C | 0.4 | 3 | 190591911 | 190678743 | 0.48 | 6.05 x10^-10^ | 159 | *GMNC* |
|  | 8 | rs11062909 | A | 0.38 | 12 | 4004752 | 4013260 | -0.81 | 3.47 x10^-25^ | 11 | *RP11-664D1.1* |
|  | 3 | rs73123652 | C | 0.08 | 12 | 65448568 | 65874956 | -0.91 | 1.03 x10^-13^ | 127 | *MSRB3:RP11-230G5.2* |
|  | 5 | rs7137149 | C | 0.11 | 12 | 117248633 | 117533214 | 1.21 | 1.50 x10^-22^ | 64 | *HRK, RP11-240G22.1* |

*SNP, single nucleotide polymorphism; BP, base pairs; Chr, chromosome; MAF, minor allele frequency*

**Table S4: Distribution of ancestries in the trans-ancestry dataset**

| **Ancestry** | **Number of participants** | **Percentage of the total dataset** |
| --- | --- | --- |
| European, validated with PCA | 31,687 | 87.17 |
| European, unvalidated | 3,604 | 9.91 |
| African | 227 | 0.62 |
| Asian | 380 | 1.01 |
| Chinese | 115 | 0.32 |
| Mixed | 163 | 0.45 |
| Other | 175 | 0.48 |
| **Total** | 36,351 | 100.00 |

**Figure S5: Comparison of PC1 and PC2 from principal component analysis of the trans-ancestry dataset**

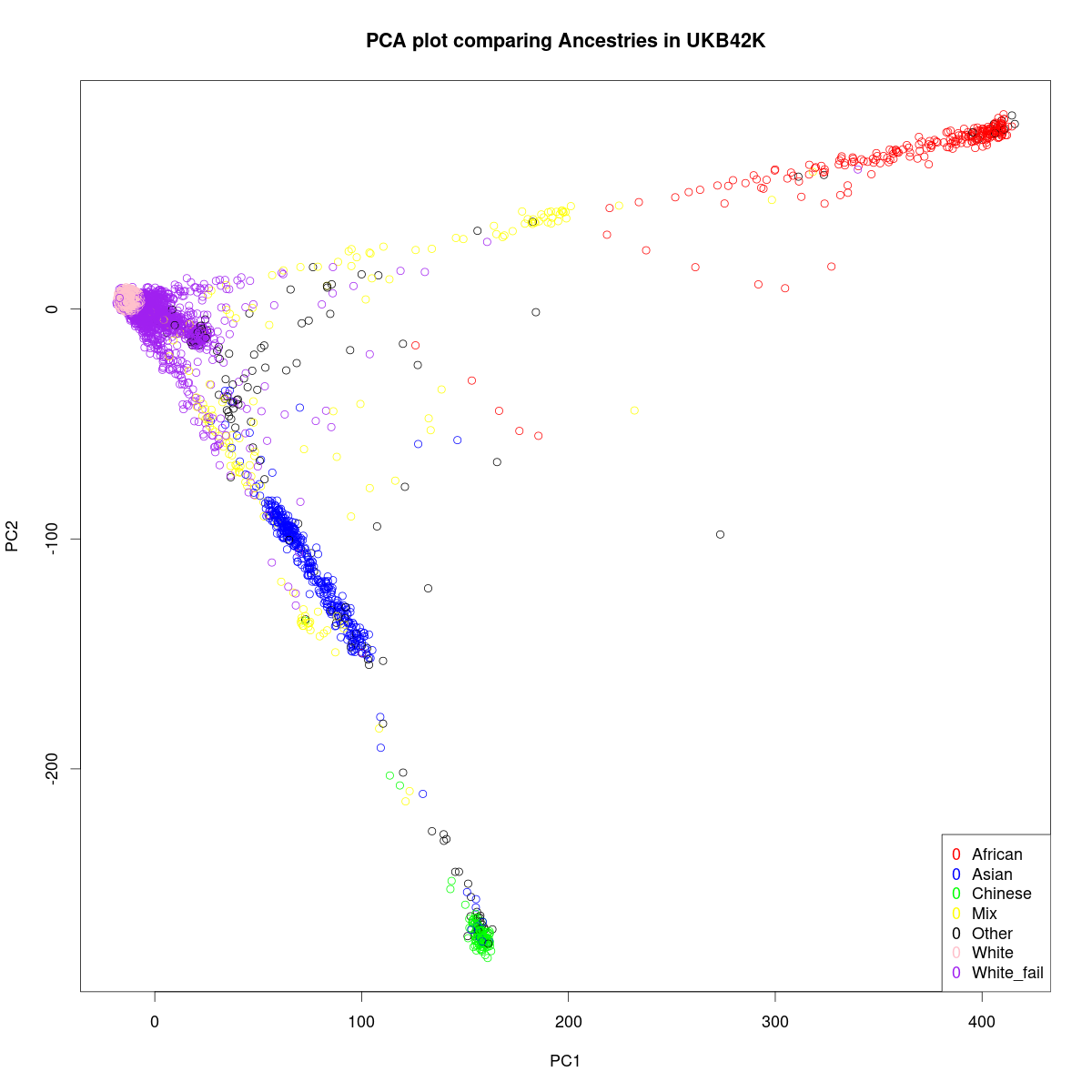

*The first principal component (PC1) is on the x-axis and the second principal component (PC2) is on the y-axis. Each participant is represented by a circle on the graph. Ancestries are color-coded as indicated in the legend on the bottom right. White_fail, individuals with self-reported European ancestry that was not confirmed with principal component analysis*

**Figure S6a: QQ plots describing MiXeR model fit for the European meta-analysis**

*The x-axis is the Empirical -log10 (q) which refers to the proportion of z-scores with p-values exceeding that threshold and the y axis is the nominal -log10 (p) which is the nominal p-value for the z-scores. Blue, GWAS curve; yellow, predicted model curve; dashed line, null hypothesis; the grey area, 95% confidence interval. Heritability is determined by the overall departure of the GWAS curve from the null line. Its curvature determines polygenicity, i.e. the point where the GWAS curve begins to bend leftwards from the null line. The slope after departure reflects the discoverability.*

*Anterior amygdaloid area	Accessory basal nucleus*

Nominal -log10 (p)

Empirical -log10 (q)

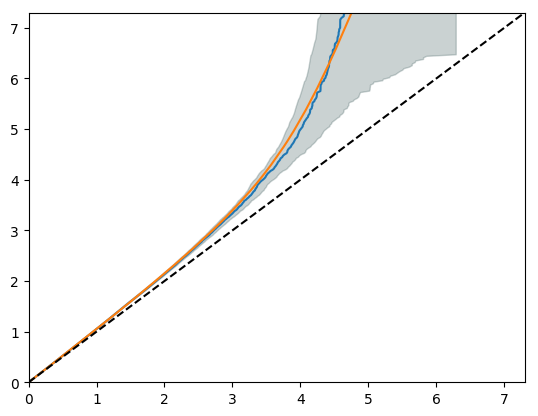

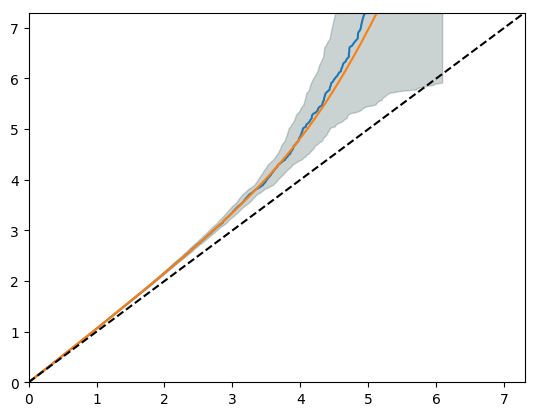

*Basal nucleus	Coricoamygdaloid transition area*

Nominal -log10 (p)

Empirical -log10 (q)

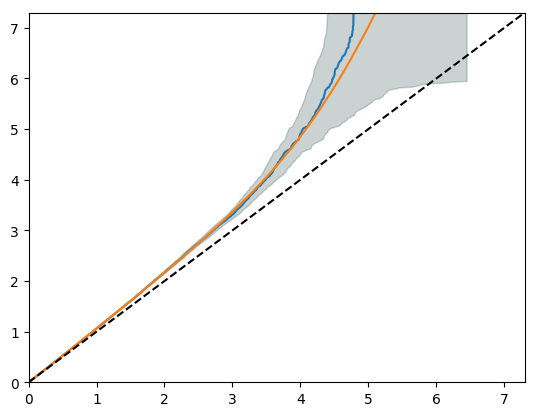

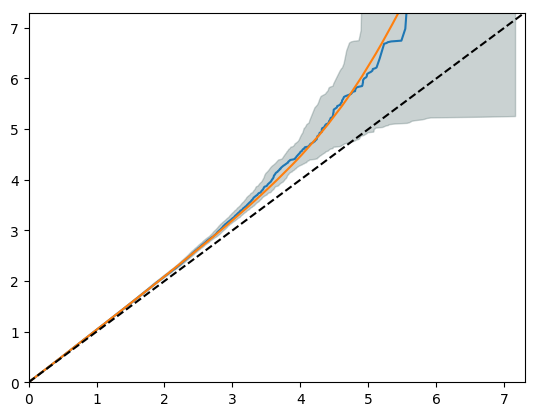

*Central nucleus	Cortical nucleus*

Nominal -log10 (p)

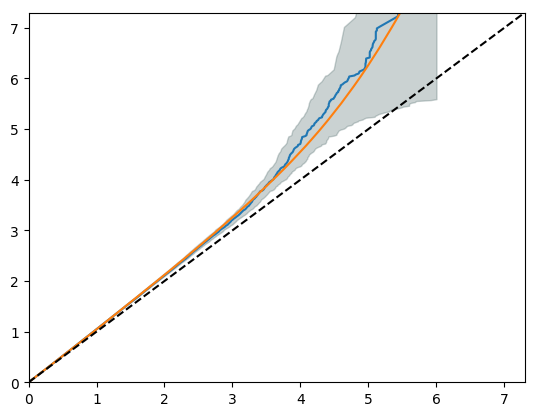

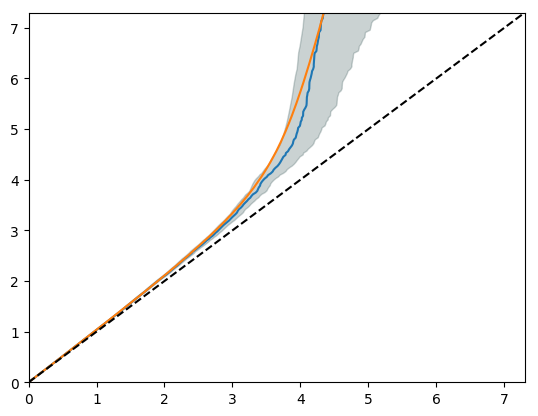

Empirical -log10 (q)

Empirical -log10 (q)

*Lateral nucleus	Medial nucleus*

Nominal -log10 (p)

Empirical -log10 (q)

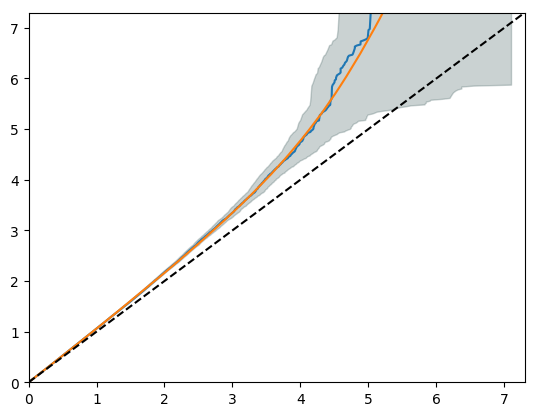

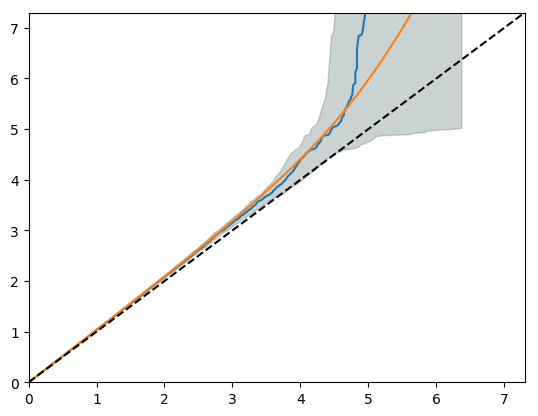

*Paralaminar nucleus	Whole amygdala*

Nominal -log10 (p)

Empirical -log10 (q)

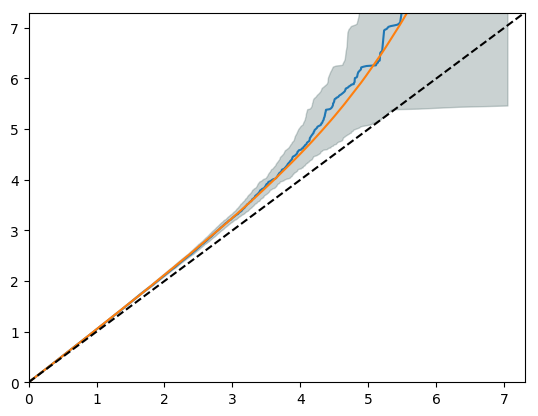

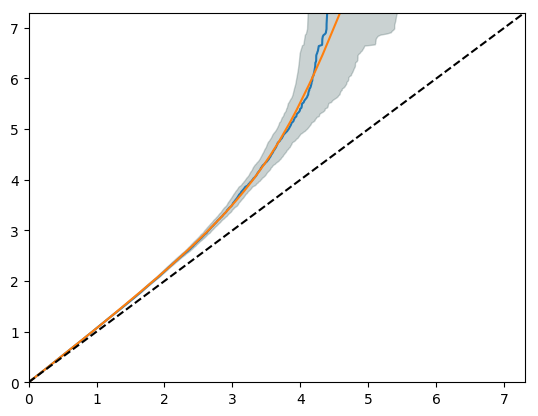

**Figure S6b: QQ plots describing MiXeR model fit for the trans-ancestry analysis**

*The x-axis is the Empirical -log10 (q) which refers to the proportion of z-scores with p-values exceeding that threshold and the y axis is the nominal -log10 (p) which is the nominal p-value for the z-scores. Blue, GWAS curve; yellow, predicted model curve; dashed line, null hypothesis; the grey area, 95% confidence interval. Heritability is determined by the overall departure of the GWAS curve from the null line. Its curvature determines polygenicity, i.e. the point where the GWAS curve begins to bend leftwards from the null line. The slope after departure reflects the discoverability.*

*Anterior amygdaloid area	Accessory basal nucleus*

Nominal -log10 (p)

Empirical -log10 (q)

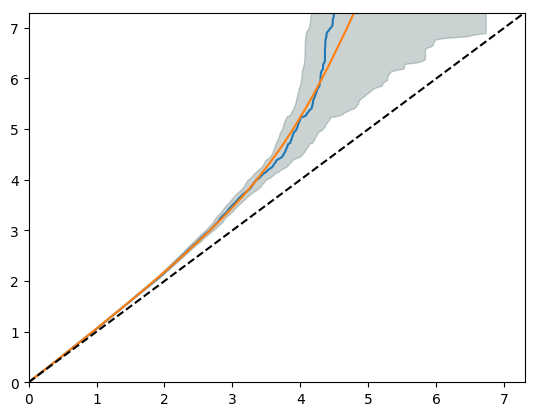

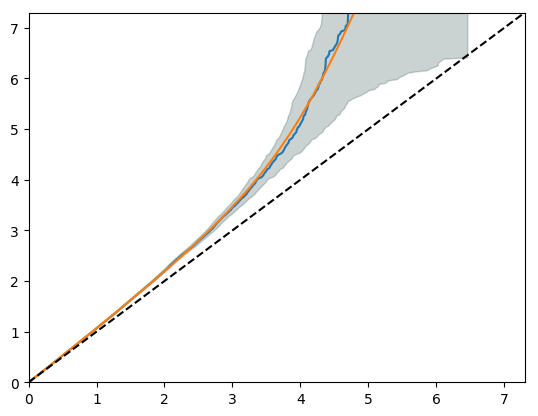

*Basal nucleus	Coricoamygdaloid transition area*

Nominal -log10 (p)

Empirical -log10 (q)

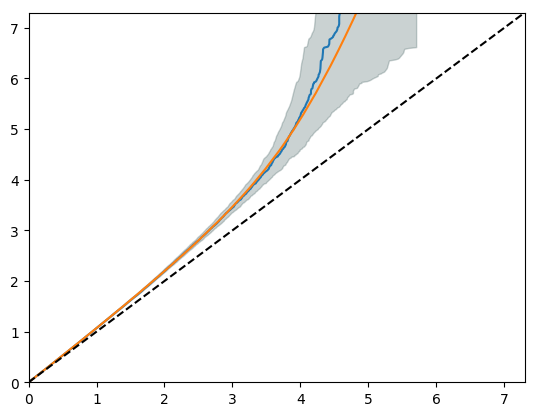

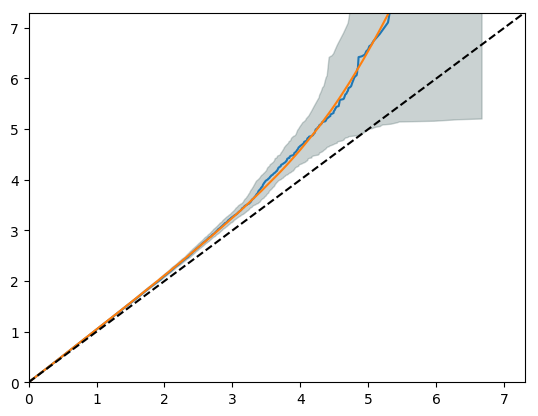

*Central nucleus	Cortical nucleus*

Nominal -log10 (p)

Empirical -log10 (q)

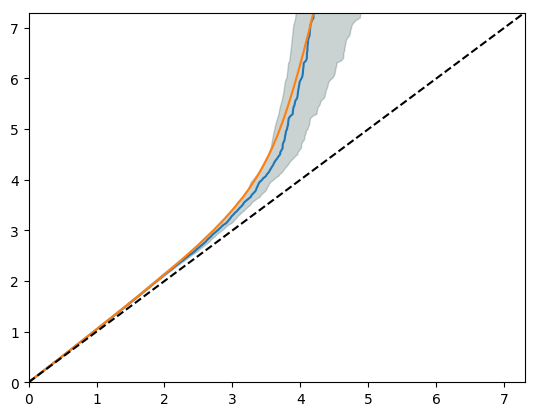

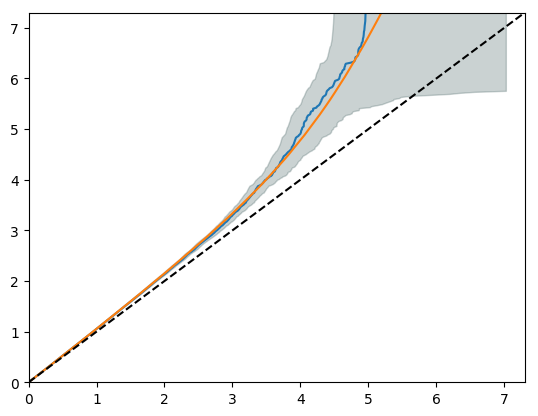

*Lateral nucleus	Medial nucleus*

Nominal -log10 (p)

Empirical -log10 (q)

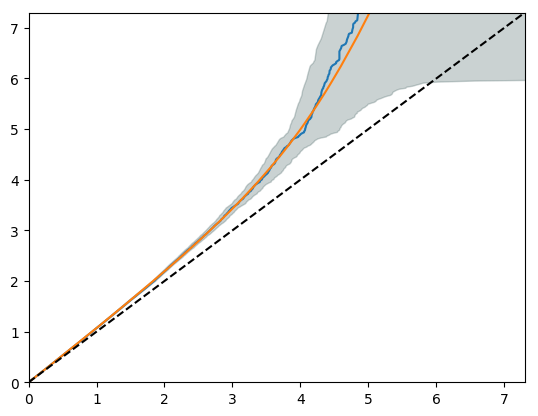

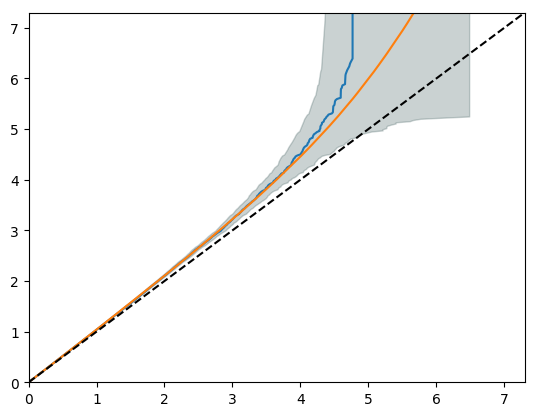

*Paralaminar nucleus	Whole amygdala*

Nominal -log10 (p)

Empirical -log10 (q)

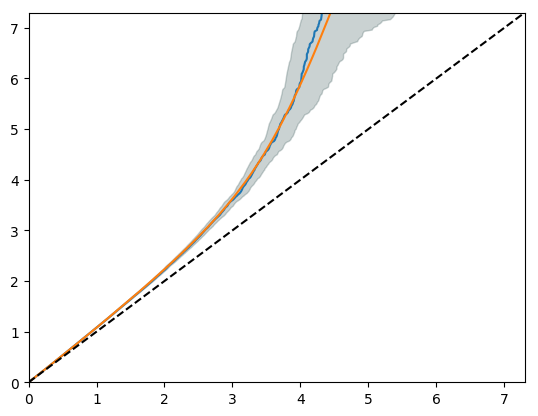

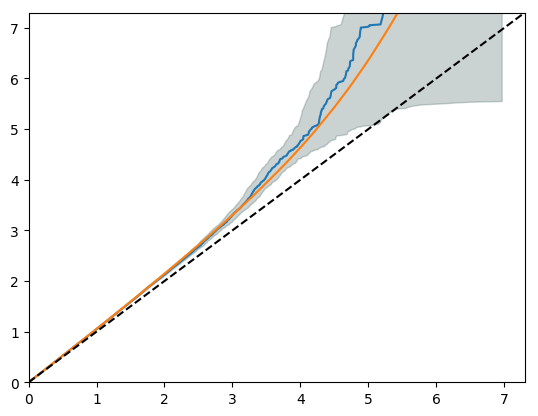

**Table S5: Heritability estimates, with full test statistics, estimated with Genome-wide complex trait analysis and Linkage Disequilibrium Score Regression for the amygdala nuclei and other subcortical regions of interest**

|  | **Region of interest** | **Genome-wide complex trait analysis** | | | **Linkage disequilibrium score regression** | | | |
| --- | --- | --- | --- | --- | --- | --- | --- | --- |
|  |  | **H2** | **SE** | **P-value** | **h^2^** | **SE** | **Lambda GC** | **Z-score** |
| European meta-analysis | Whole amygdala | 0.37 | 0.02 | 1.00 x10^-16^ | 0.28 | 0.02 | 1.15 | 14.00 |
|  | Anterior amygdaloid area | 0.28 | 0.02 | 1.00 x10^-16^ | 0.23 | 0.03 | 1.12 | 11.50 |
|  | Accessory basal nucleus | 0.33 | 0.02 | 1.00 x10^-16^ | 0.23 | 0.02 | 1.12 | 11.50 |
|  | Basal nucleus | 0.32 | 0.02 | 1.00 x10^-16^ | 0.24 | 0.02 | 1.15 | 12.00 |
|  | Corticoamygdaloid transition area | 0.17 | 0.02 | 1.00 x10^-16^ | 0.11 | 0.02 | 1.07 | 5.50 |
|  | Cortical nucleus | 0.28 | 0.02 | 1.00 x10^-16^ | 0.18 | 0.02 | 1.10 | 9.00 |
|  | Central nucleus | 0.20 | 0.02 | 1.00 x10^-16^ | 0.16 | 0.20 | 1.09 | 8.00 |
|  | Lateral nucleus | 0.31 | 0.02 | 1.00 x10^-16^ | 0.22 | 0.02 | 1.13 | 11.00 |
|  | Medial nucleus | 0.19 | 0.02 | 1.00 x10^-16^ | 0.15 | 0.02 | 1.08 | 7.50 |
|  | Paralaminar nucleus | 0.29 | 0.02 | 1.00 x10^-16^ | 0.22 | 0.02 | 1.12 | 11.00 |
|  | Accumbens | 0.31 | 0.02 | 1.00 x10^-16^ | 0.26 | 0.02 | 1.16 | 13.00 |
|  | Caudate nucleus | 0.47 | 0.02 | 1.00 x10^-16^ | 0.36 | 0.03 | 1.20 | 18.00 |
|  | Hippocampus | 0.32 | 0.02 | 1.00 x10^-16^ | 0.30 | 0.02 | 1.17 | 15.00 |
|  | Pallidum | 0.34 | 0.02 | 1.00 x10^-16^ | 0.25 | 0.02 | 1.00 | 12.50 |
|  | Putamen | 0.32 | 0.02 | 1.00 x10^-16^ | 0.29 | 0.02 | 1.02 | 14.50 |
|  | Thalamus | 0.32 | 0.02 | 1.00 x10^-16^ | 0.27 | 0.02 | 1.01 | 13.50 |
| Trans-ancestry analysis | Whole amygdala | 0.37 | 0.02 | 1.00 x10^-16^ | 0.28 | 0.02 | 1.17 | 14.00 |
|  | Anterior amygdaloid area | 0.30 | 0.02 | 1.00 x10^-16^ | 0.21 | 0.03 | 1.13 | 7.00 |
|  | Accessory basal nucleus | 0.34 | 0.02 | 1.00 x10^-16^ | 0.22 | 0.02 | 1.14 | 11.00 |
|  | Basal nucleus | 0.32 | 0.02 | 1.00 x10^-16^ | 0.22 | 0.02 | 1.15 | 11.00 |
|  | Corticoamygdaloid transition area | 0.18 | 0.02 | 1.00 x10^-16^ | 0.12 | 0.02 | 1.08 | 6.00 |
|  | Cortical nucleus | 0.29 | 0.02 | 1.00 x10^-16^ | 0.18 | 0.02 | 1.11 | 9.00 |
|  | Central nucleus | 0.20 | 0.02 | 1.00 x10^-16^ | 0.14 | 0.02 | 1.10 | 7.00 |
|  | Lateral nucleus | 0.32 | 0.02 | 1.00 x10^-16^ | 0.21 | 0.02 | 1.15 | 10.50 |
|  | Medial nucleus | 0.20 | 0.02 | 1.00 x10^-16^ | 0.15 | 0.02 | 1.09 | 7.50 |
|  | Paralaminar nucleus | 0.29 | 0.02 | 1.00 x10^-16^ | 0.22 | 0.02 | 1.14 | 11.00 |
|  | Accumbens | 0.34 | 0.02 | 1.00 x10^-16^ | 0.26 | 0.02 | 1.17 | 13.00 |
|  | Caudate nucleus | 0.52 | 0.02 | 1.00 x10^-16^ | 0.34 | 0.03 | 1.21 | 11.33 |
|  | Hippocampus | 0.41 | 0.02 | 1.00 x10^-16^ | 0.30 | 0.03 | 1.18 | 10.00 |
|  | Pallidum | 0.30 | 0.02 | 1.00 x10^-16^ | 0.23 | 0.02 | 1.14 | 11.50 |
|  | Putamen | 0.40 | 0.02 | 1.00 x10^-16^ | 0.28 | 0.03 | 1.17 | 9.33 |
|  | Thalamus | 0.39 | 0.02 | 1.00 x10^-16^ | 0.26 | 0.02 | 1.17 | 13.00 |

*h^2^, heritability; SE, standard error; GC, genomic control*

**Figure S7: The relationship between the average nucleus volume and heritability estimates**

| Mean volume (mm3) | SNP-based heritability |
| --- | --- |
| 90.36 | 0.2 |
| 1328.5 | 0.31 |
| 43.11 | 0.19 |
| 50.97 | 0.28 |
| 101.61 | 0.29 |
| 118.01 | 0.28 |
| 527.6 | 0.33 |
| 358.8 | 0.17 |
| 1423.3 | 0.32 |

*The mean nucleus volume in cubic millimetre is on the x-axis and heritability estimate is on the y-axis per nucleus. PAR, paralaminar nucleus; CAT, corticoamygdaloid transition area; COR, cortical nucleus, MED, medial nucleus; CEN, central nucleus; AAA, anterior amygdaloid area; ACC, accessory basal nucleus; BAS, basal nucleus; LAT, lateral nucleus.*

**Figure S8. Manhattan and QQ plots for the European meta-analysis of whole amygdala volume, and per nucleus volume corrected for whole amygdala volume.**

*For the Manhattan plots, the red line indicates the adjusted whole-genome significance threshold (5x10^-9^ p-value) and the blue line the standard genome-wide significance threshold (1x10^-8^).*

*Whole amygdala*

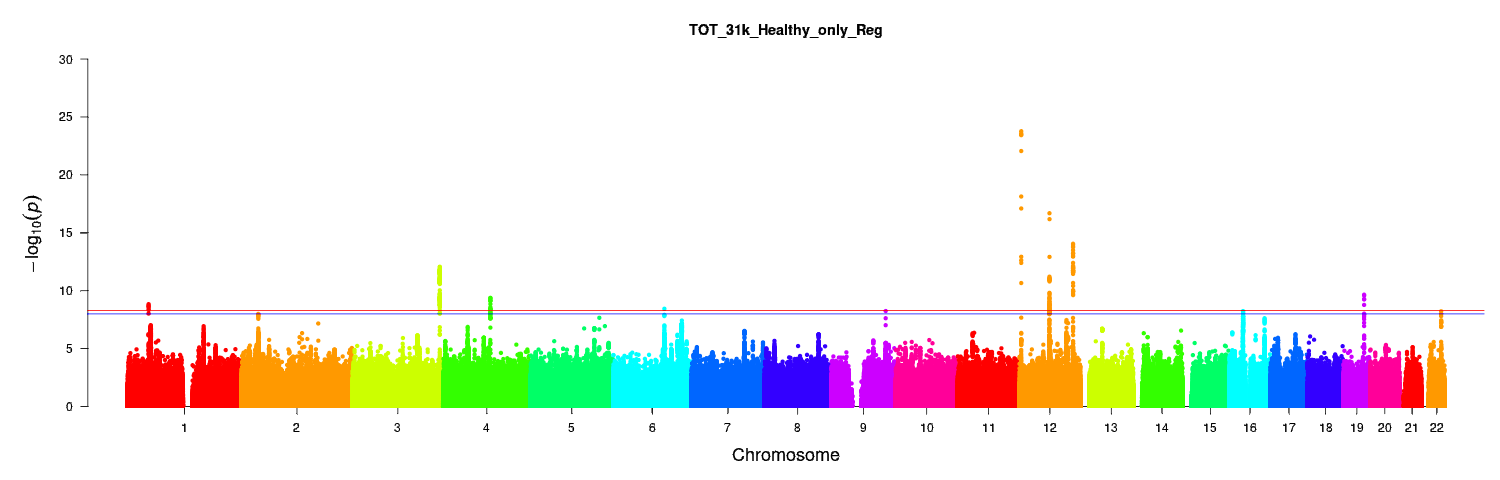

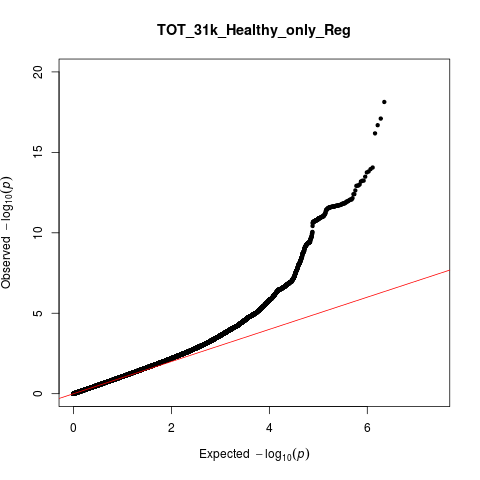

λ_GC_=1.08

*Anterior amygdaloid area*

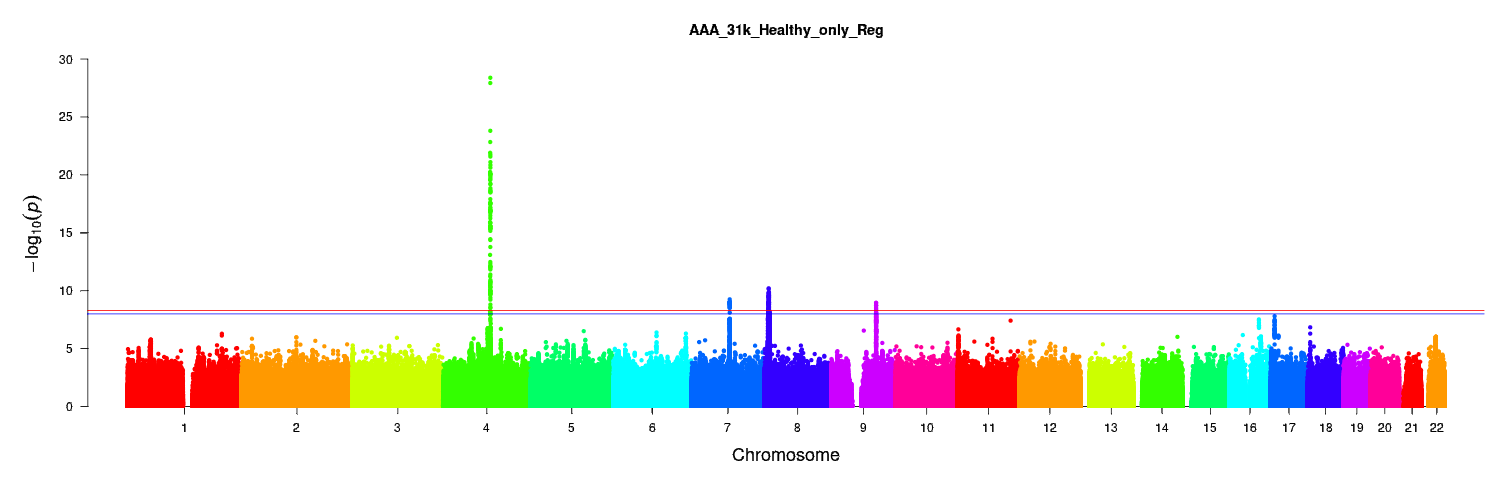

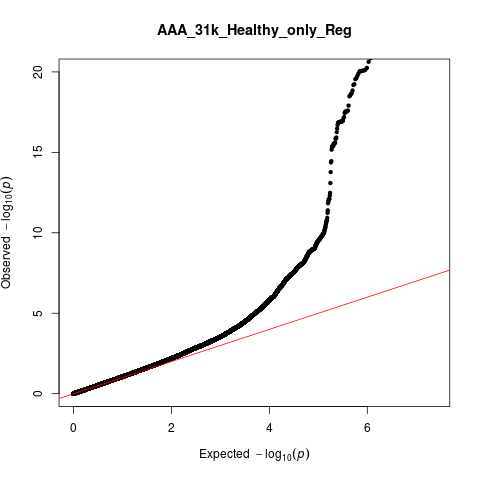

λ_GC_=1.07

*Accessory basal nucleus*

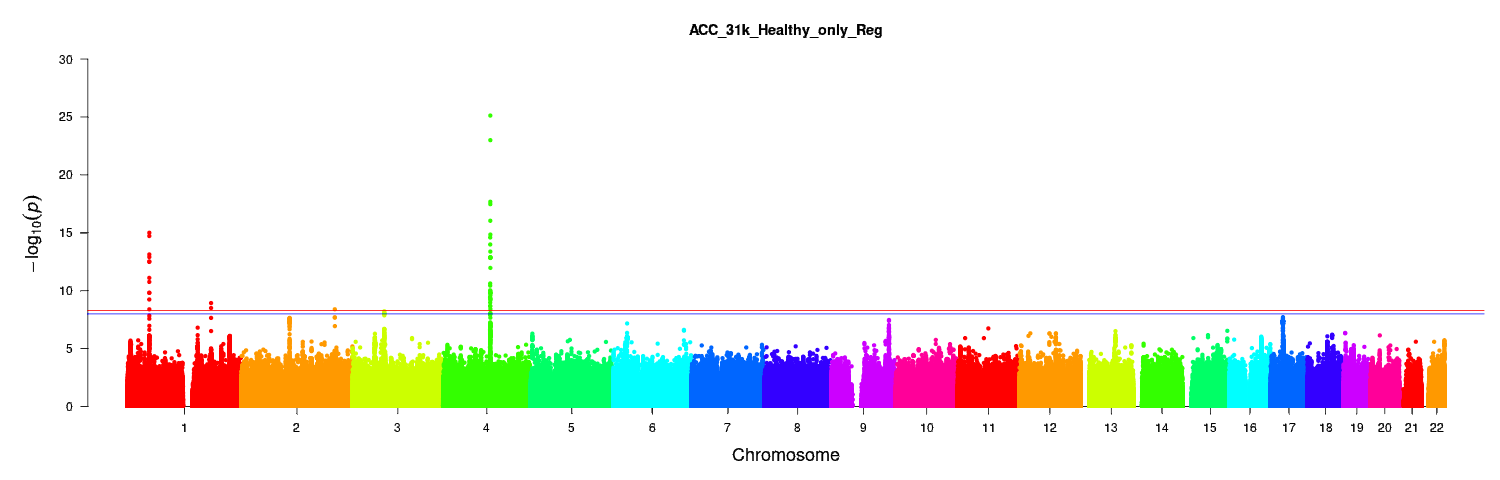

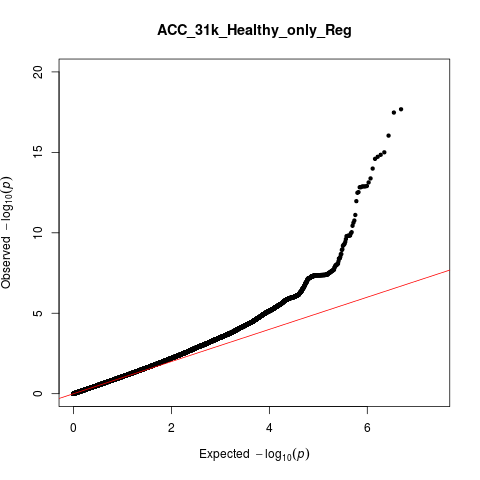

λ_GC_=1.07

*Basal nucleus*

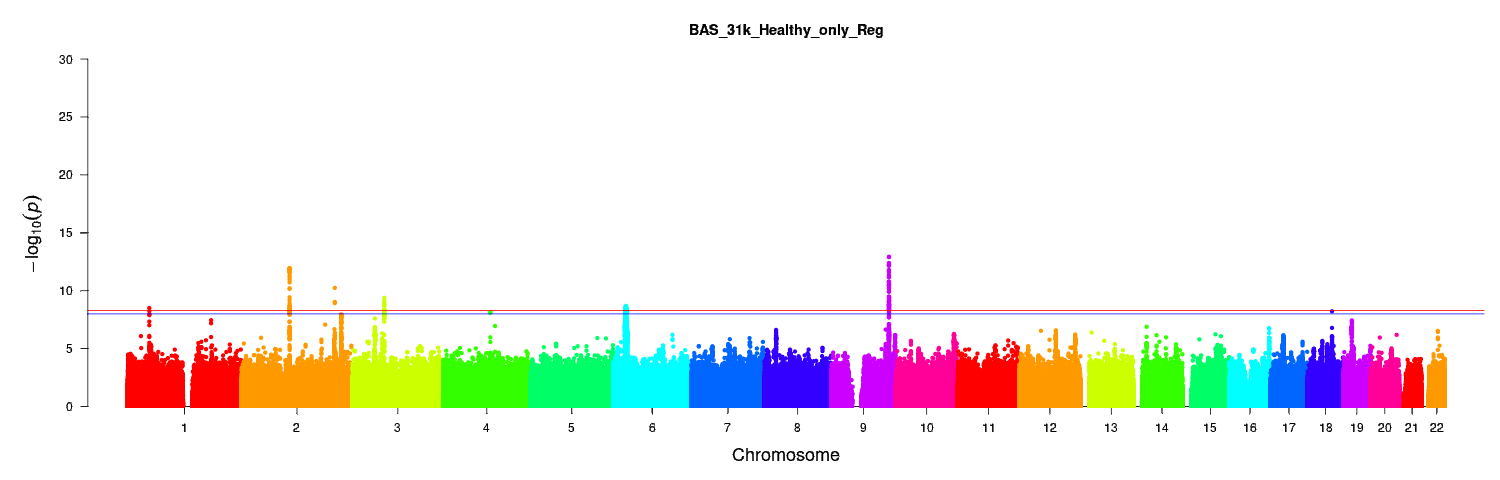

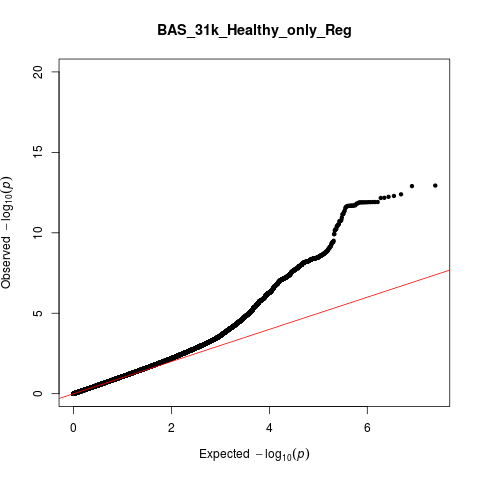

λ_GC_=1.08

*Corticoamygdaloid transition area*

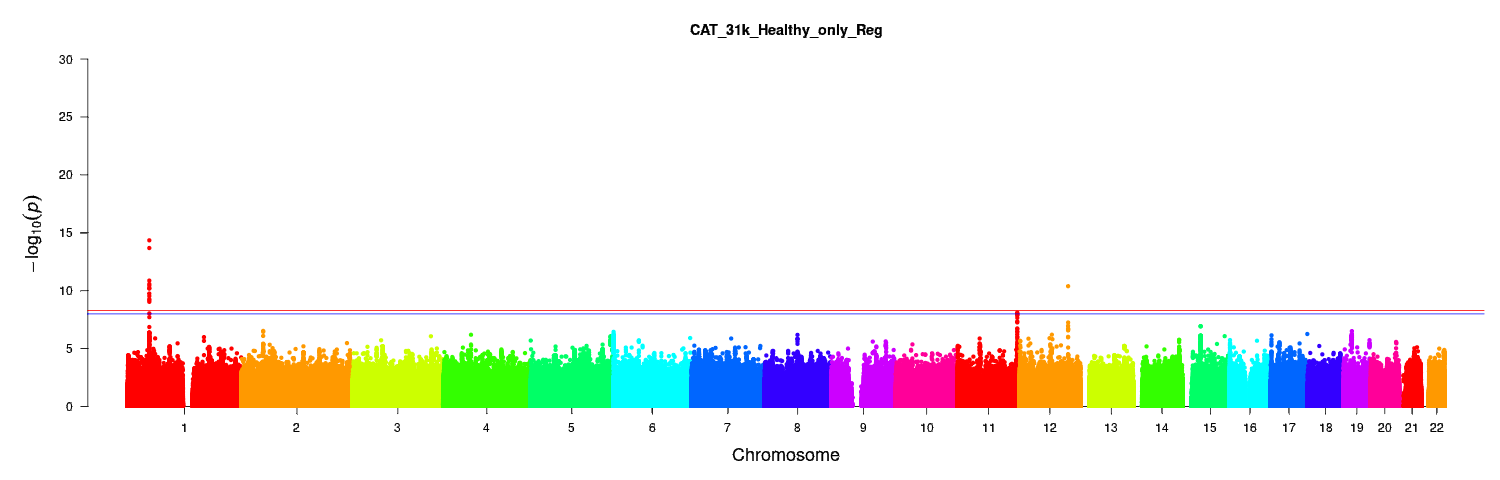

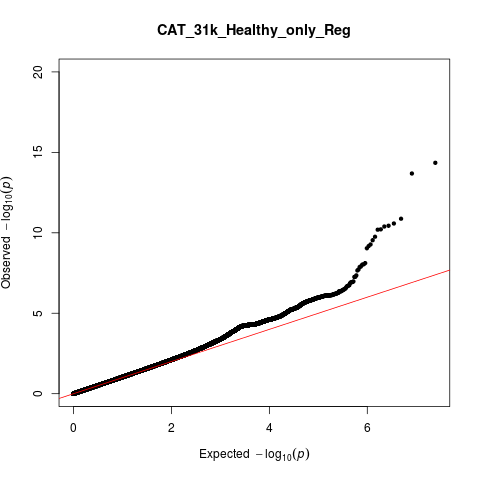

λ_GC_=1.04

*Central nucleus*

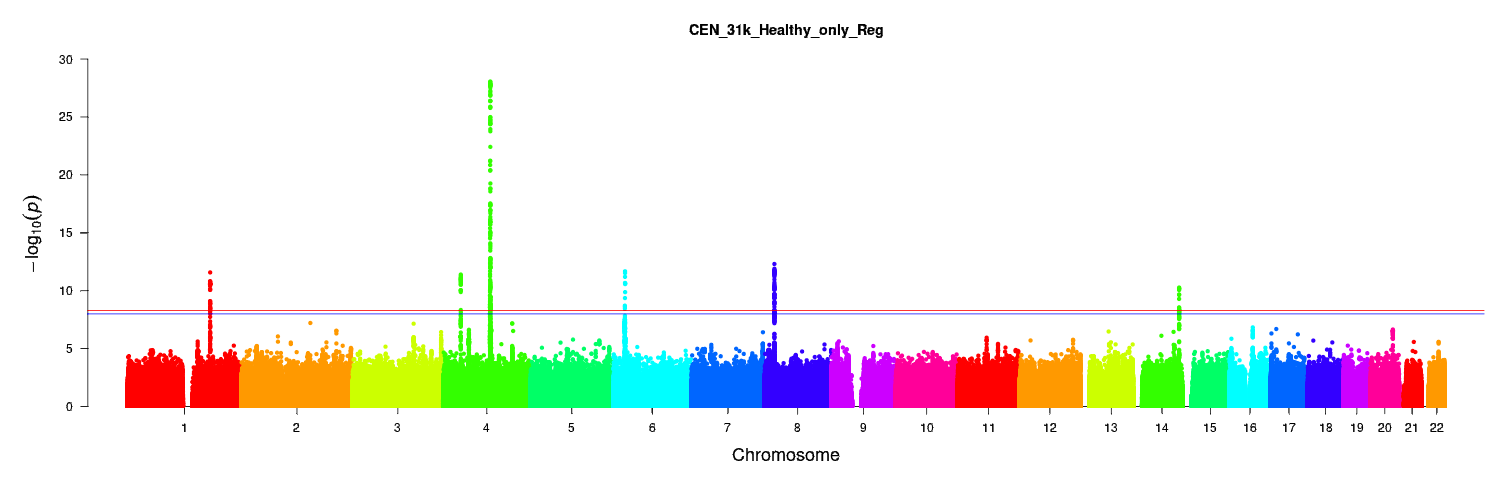

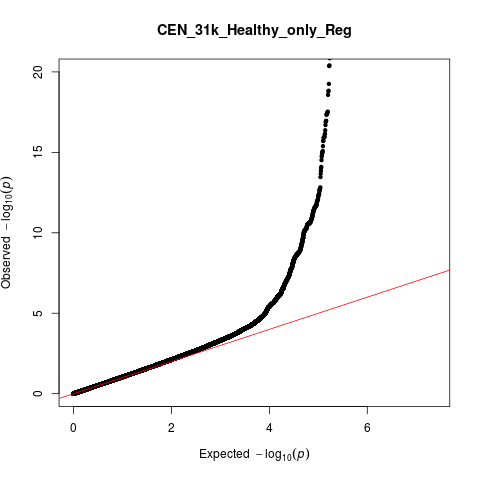

λ_GC_=1.05

*Cortical nucleus*

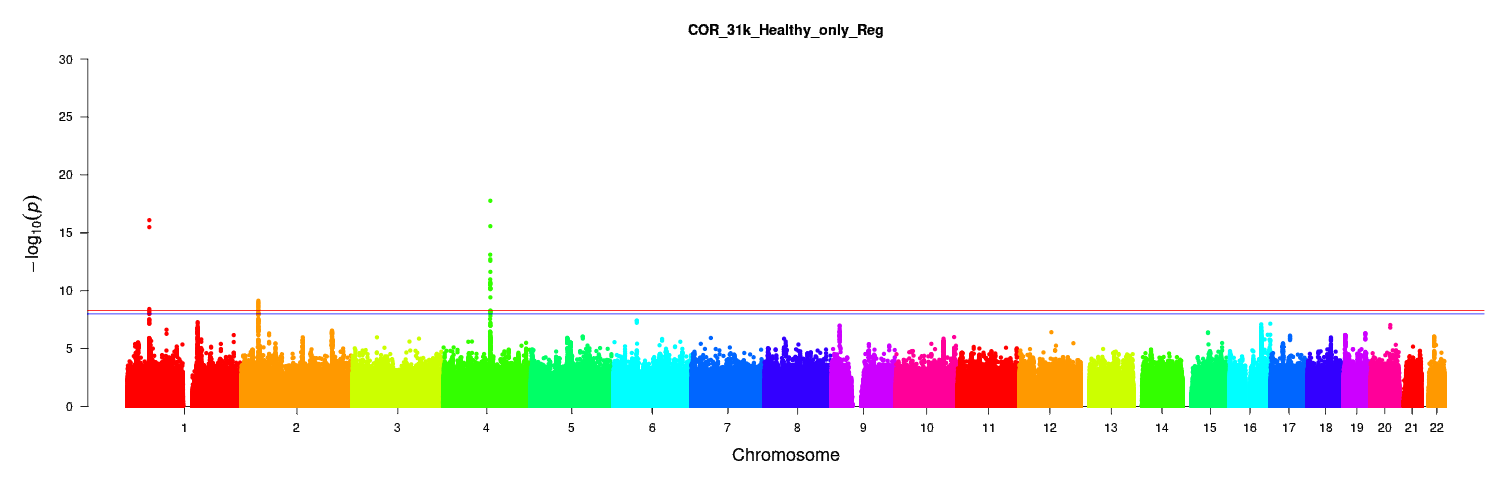

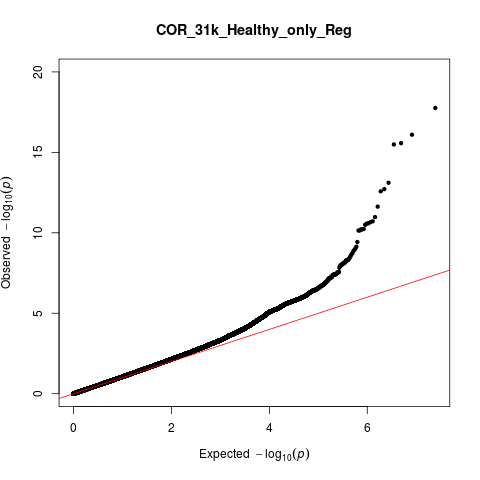

λ_GC_=1.06

*Lateral nucleus*

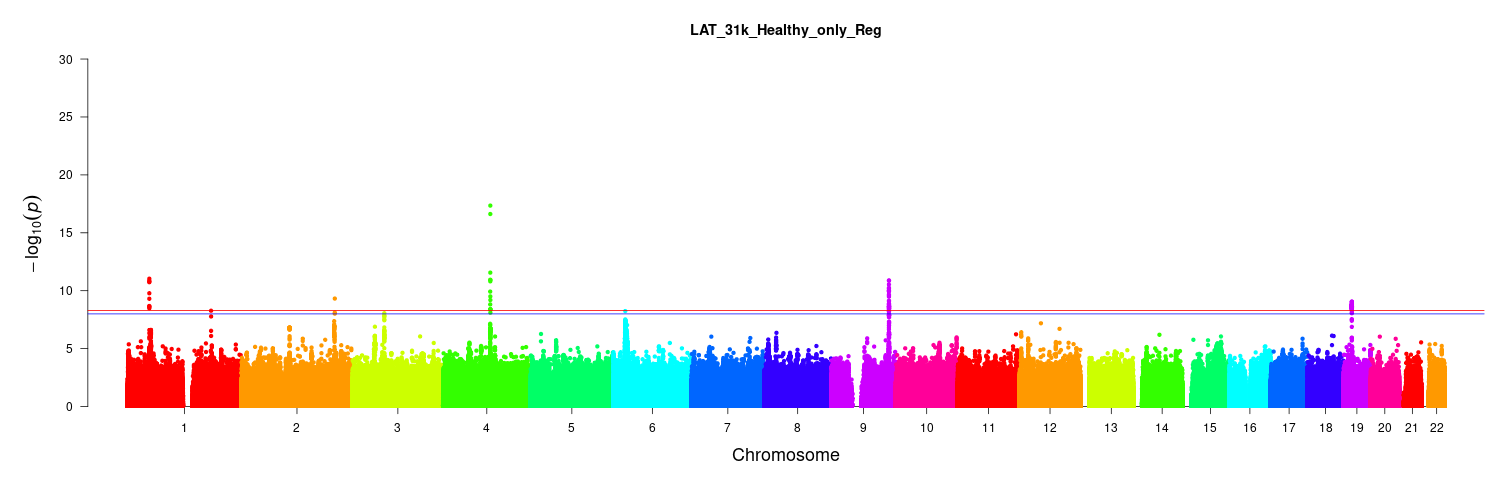

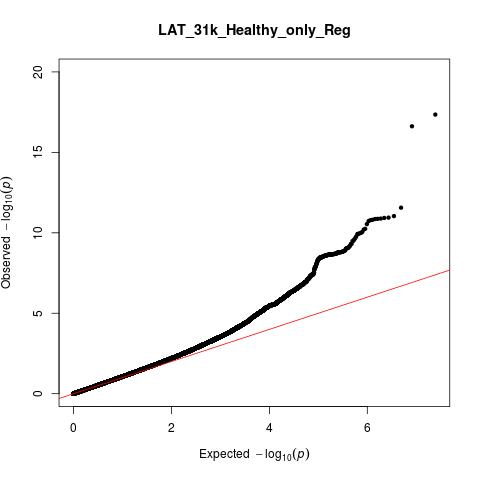

λ_GC_=1.08

*Medial Nucleus*

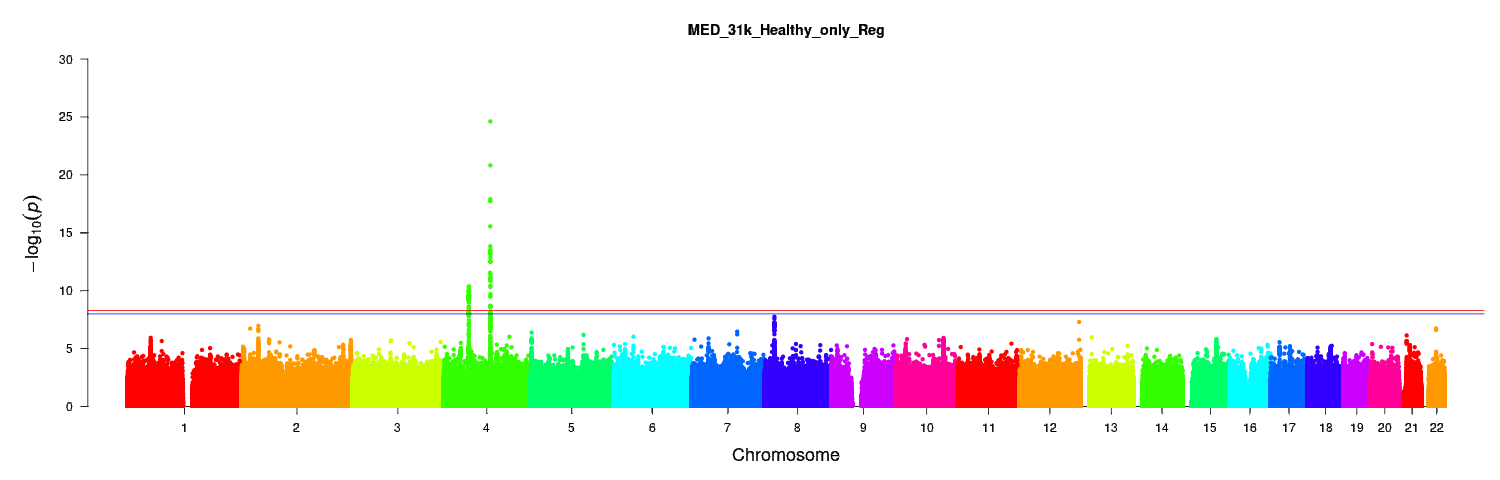

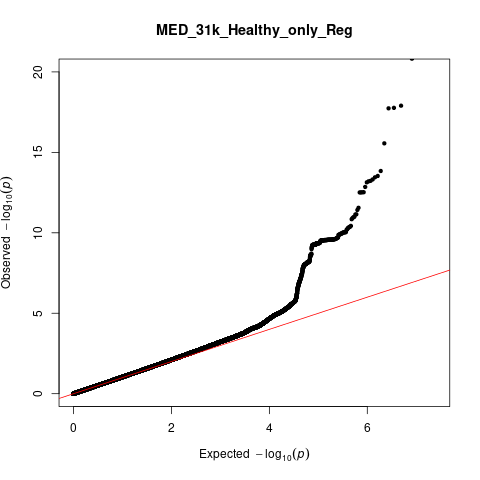

λ_GC_=1.05

*Paralaminar nucleus*

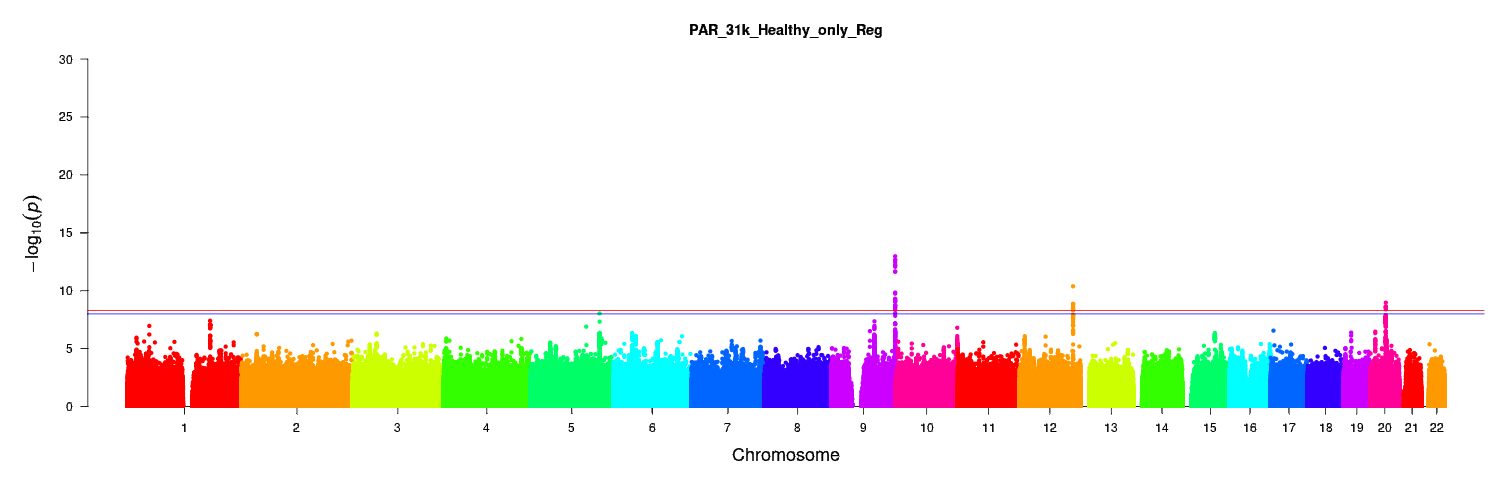

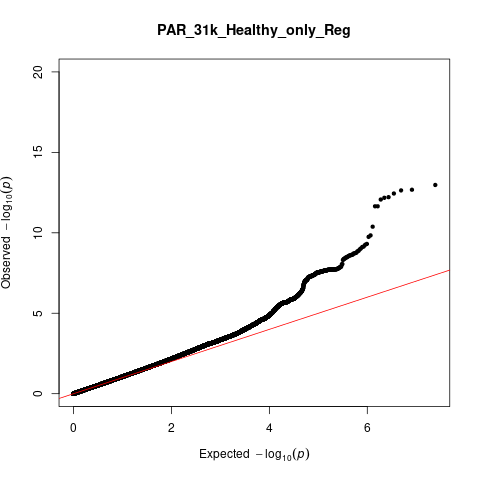

λ_GC_=1.07

**Figure S9: The relationship between nucleus volume and the number of significant SNPs**

| Mean volume (mm3) | No. of significant SNPs |
| --- | --- |
| 358.8 | 14 |
| 50.97 | 27 |
| 527.6 | 59 |
| 1328.5 | 136 |
| 43.11 | 229 |
| 1423.3 | 260 |
| 90.36 | 453 |
| 101.61 | 97 |
| 118.01 | 283 |

*The mean nucleus volume in cubic millimetre is on the x-axis and the number of significant SNPs is on the y-axis per nucleus. PAR, paralaminar nucleus; CAT, corticoamygdaloid transition area; COR, cortical nucleus, MED, medial nucleus; CEN, central nucleus; AAA, anterior amygdaloid area; ACC, accessory basal nucleus; BAS, basal nucleus; LAT, lateral nucleus*

**Figure S10: Manhattan and QQ plots for the trans-ancestry GWAS of whole amygdala volume and per nucleus volume corrected for whole amygdala volume.**

*For the Manhattan plots, the red line indicates the adjusted whole-genome significance threshold (5x10^-9^ p-value) and the blue line the standard genome-wide significance threshold (1x10^-8^).*

*Whole amygdala*

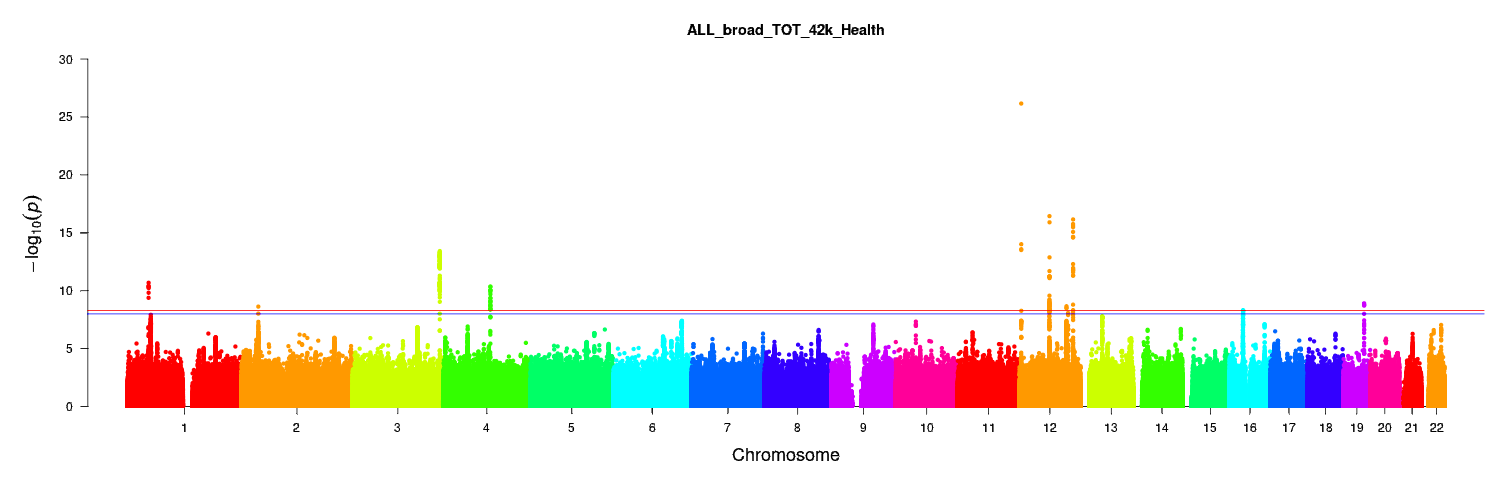

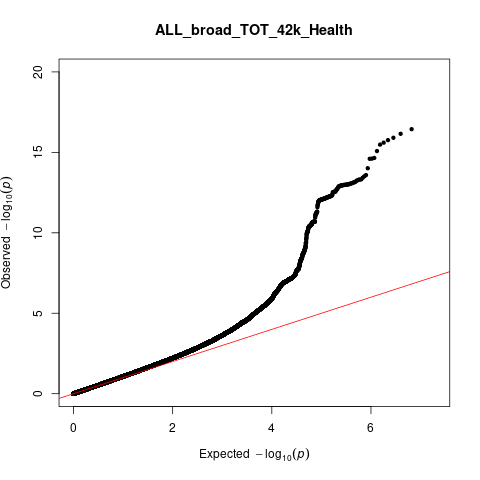

λ_GC_=1.08

*Anterior amygdaloid area*

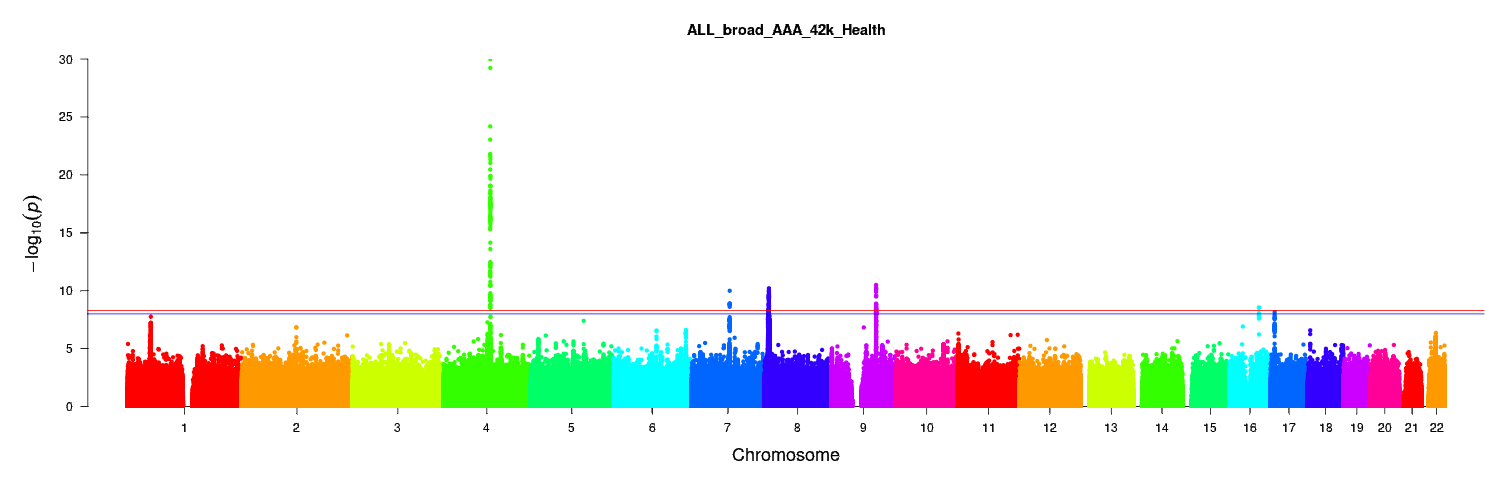

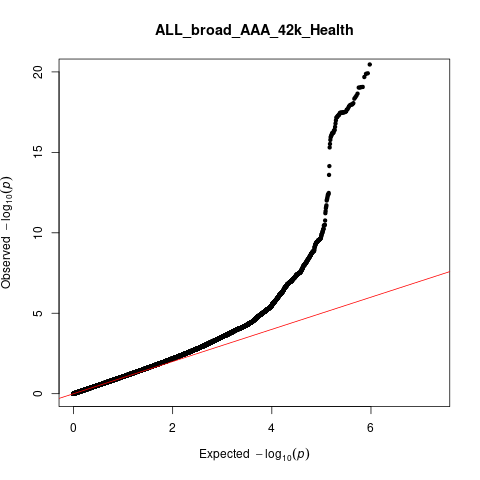

λ_GC_=1.08

*Accessory basal nucleus*

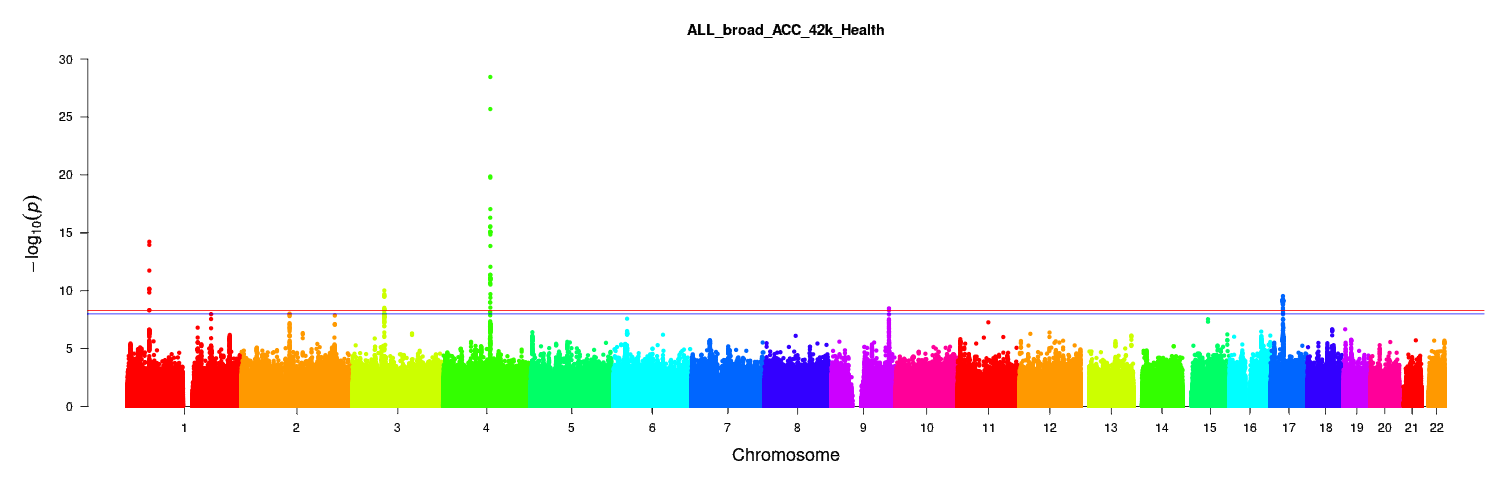

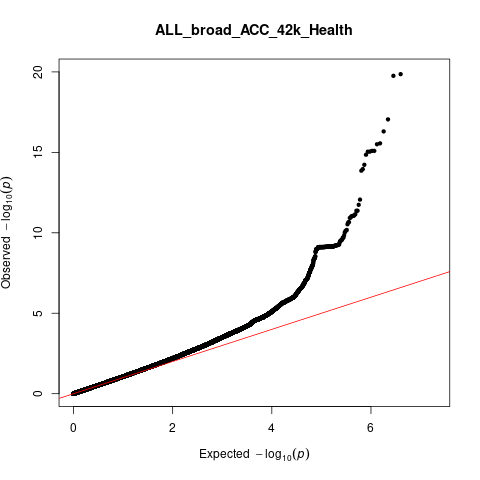

λ_GC_=1.07

*Basal nucleus*

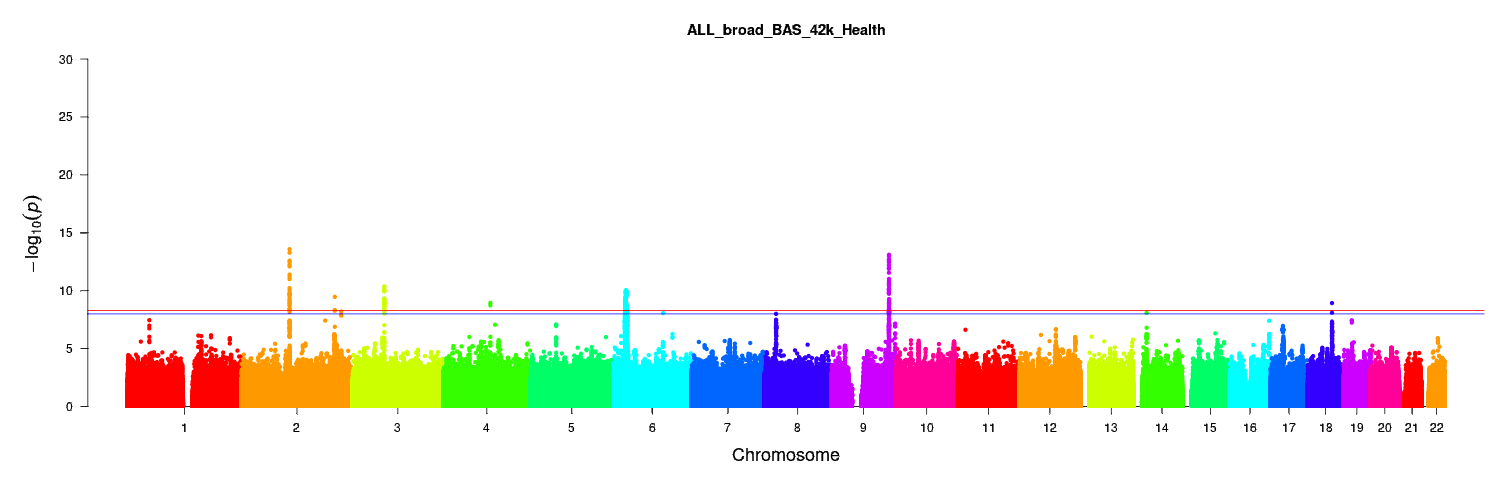

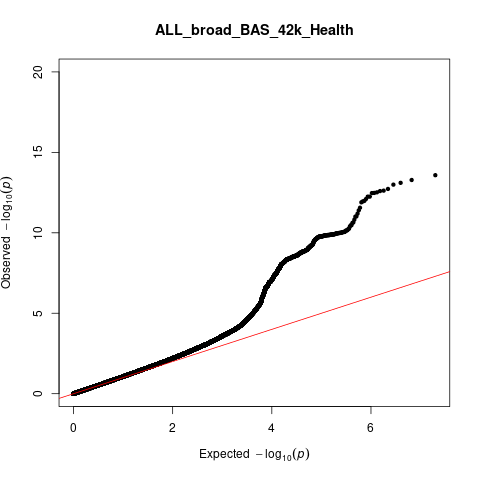

λ_GC_=1.08

*Central nucleus*

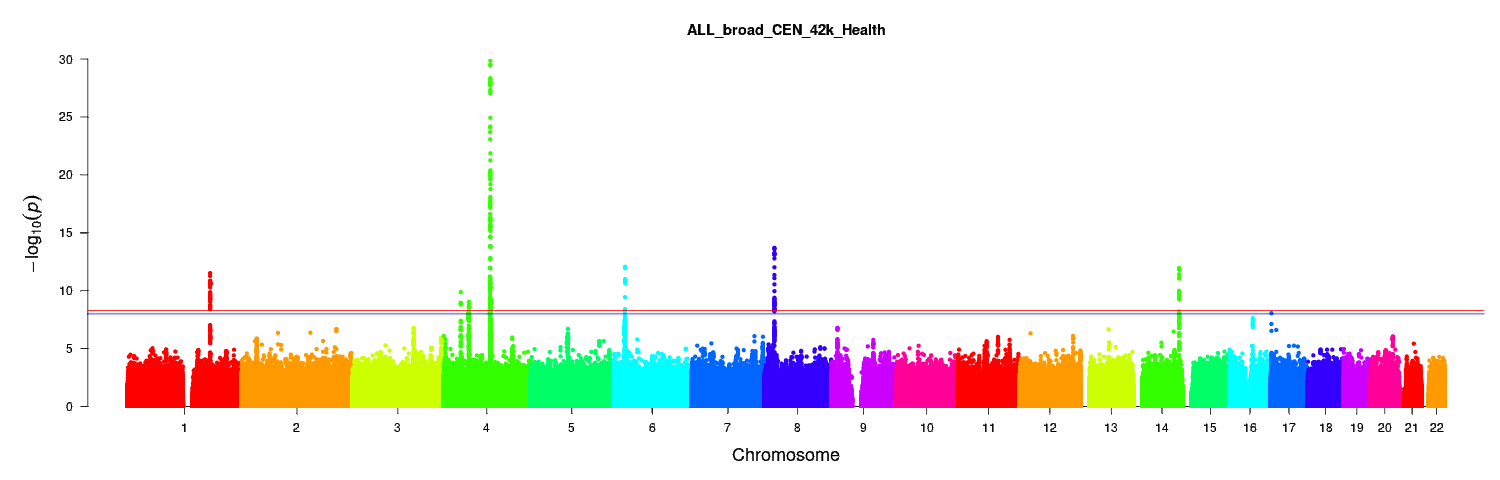

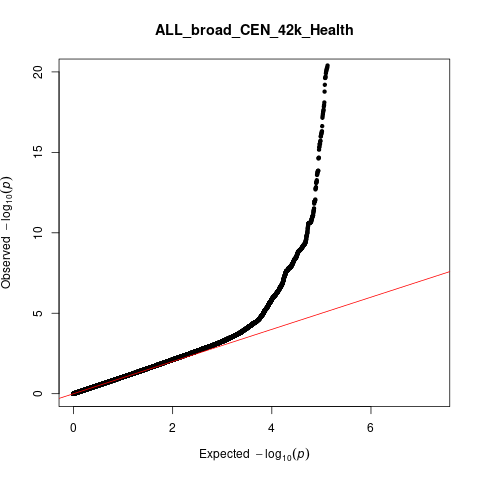

λ_GC_=1.05

*Corticoamygdaloid transition area*

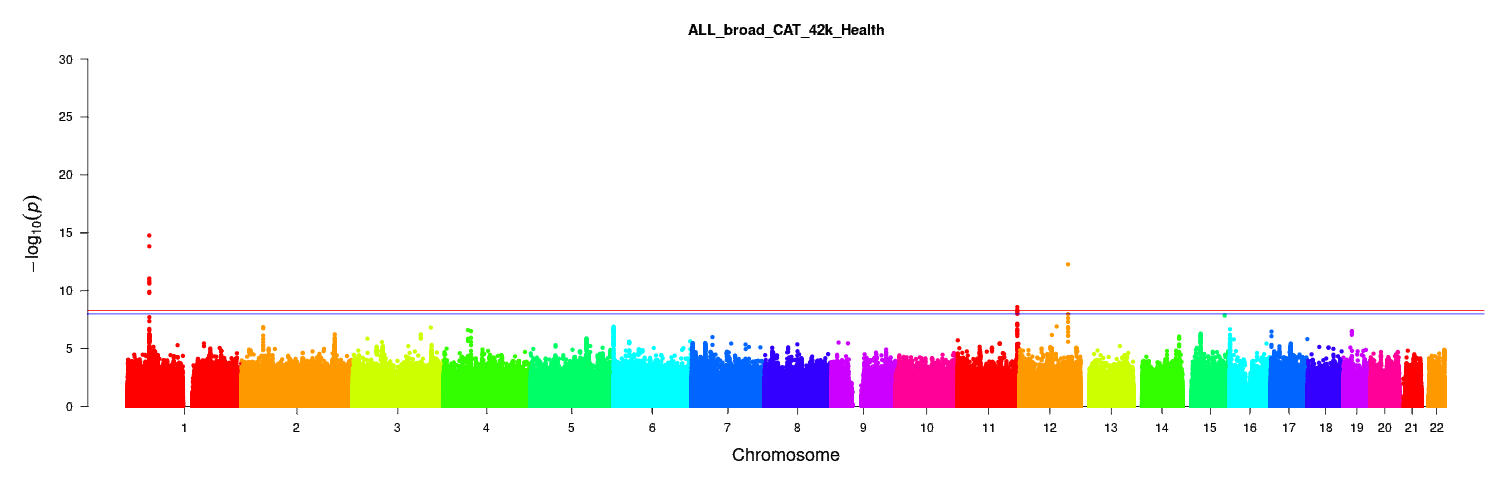

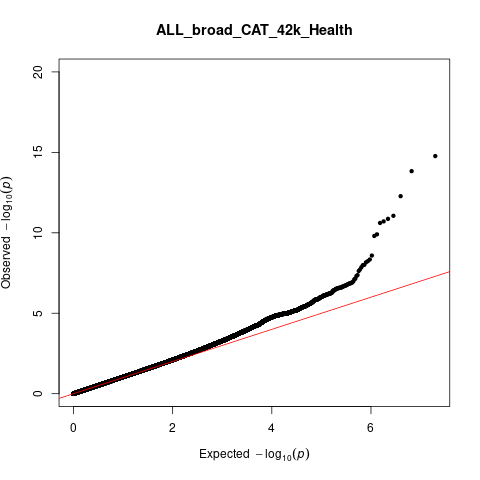

λ_GC_=1.05

*Cortical nucleus*

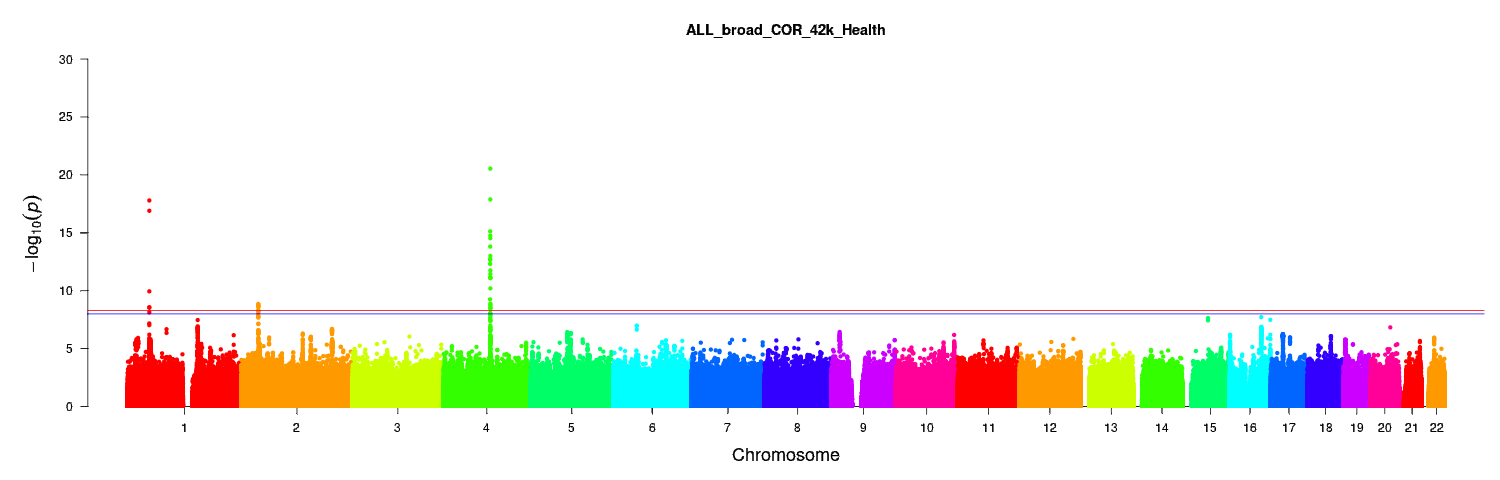

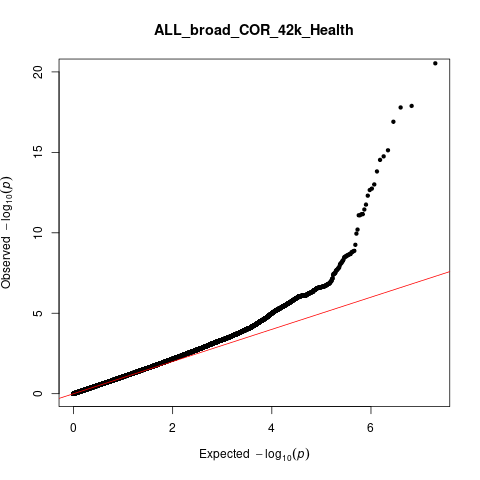

λ_GC_=1.06

*Lateral nucleus*

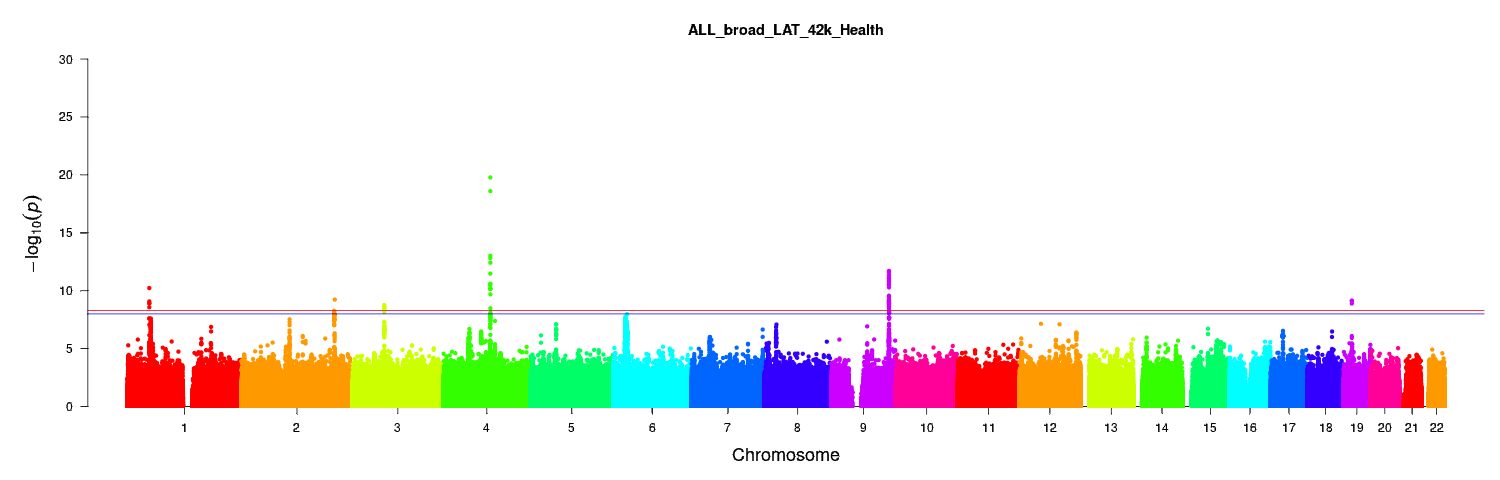

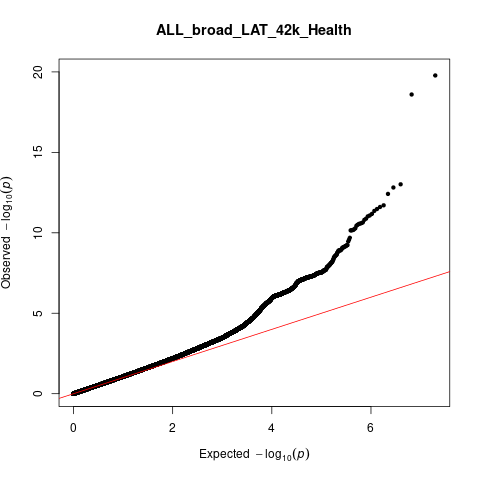

λ_GC_=1.08

*Medial Nucleus*

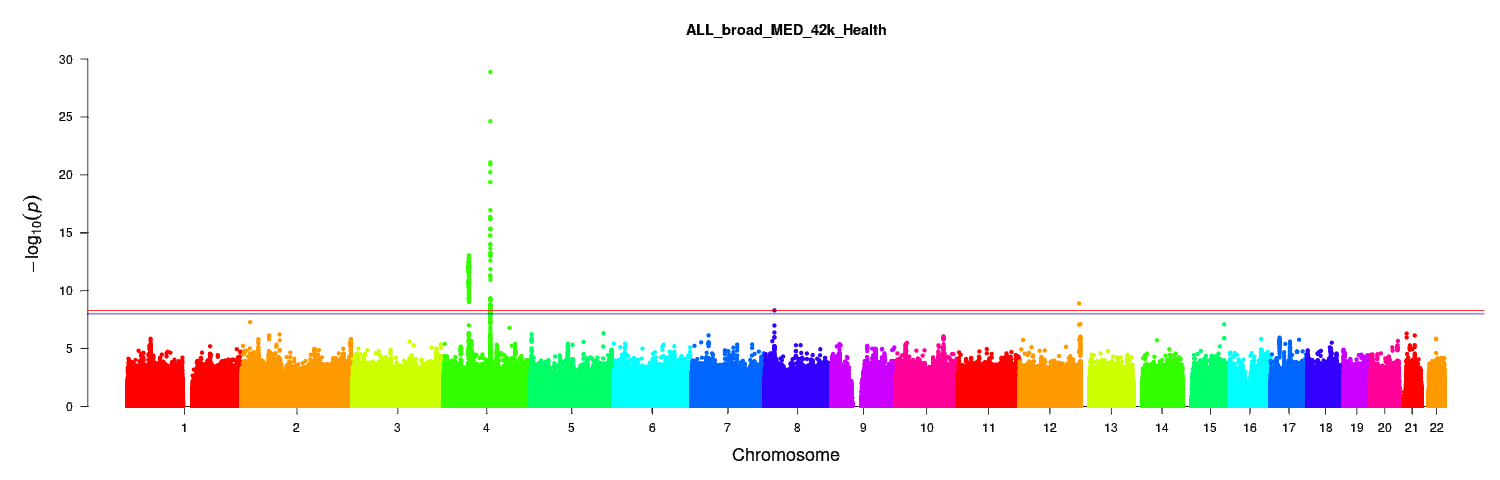

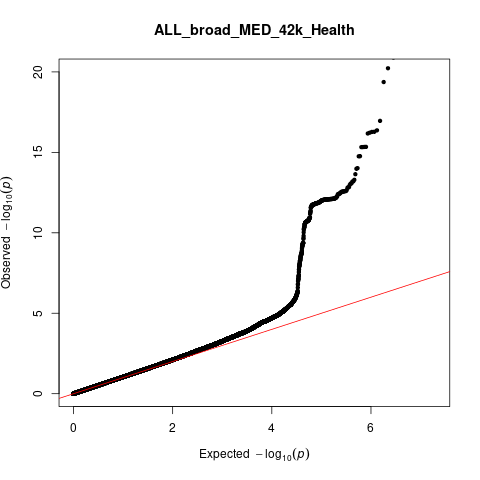

λ_GC_=1.05

*Paralaminar nucleus*

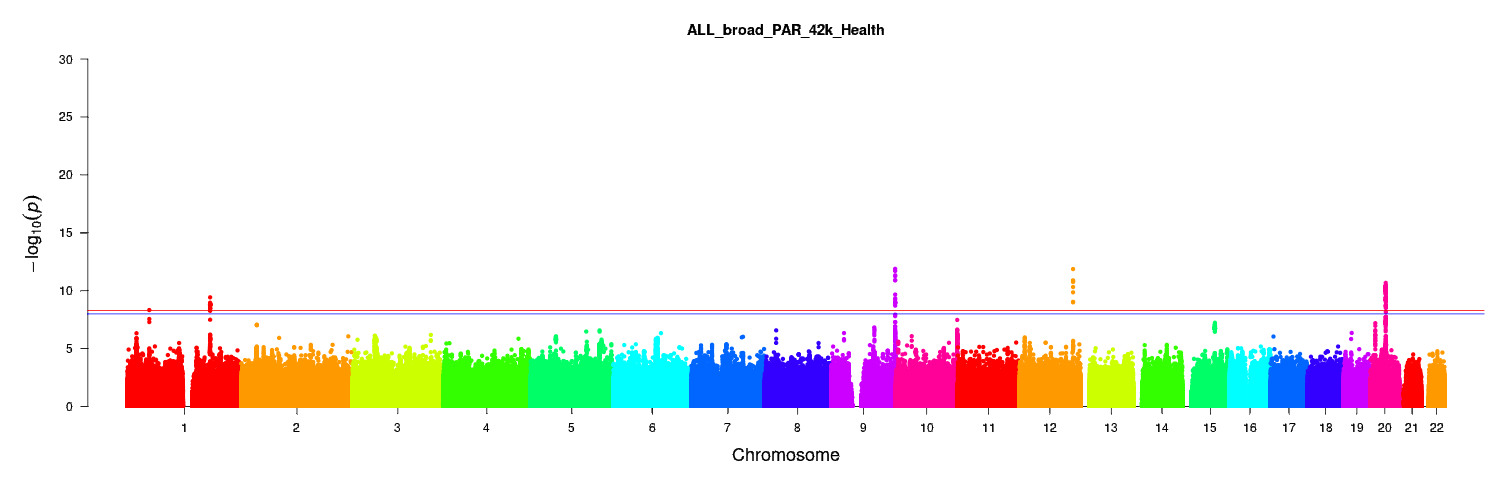

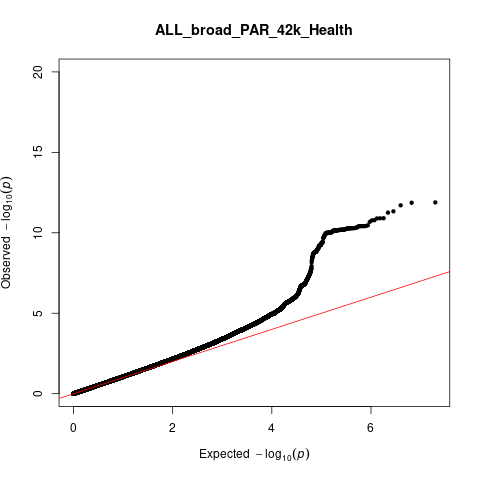

λ_GC_=1.07

**Table S6: Trending significant (P<0.05) associations from LDHUB across amygdala nuclei volumes**

| Nucleus | Traits | PubMed ID | rg | se | z | P |
| --- | --- | --- | --- | --- | --- | --- |
| Whole amygdala | Mean Hippocampus | 25607358 | 0.61 | 0.12 | 5.02 | 5.26 x10^-05^* |
|  | Urinary albumin-to-creatinine ratio | 26631737 | -0.28 | 0.08 | -3.33 | 9 x10^-04^ |
|  | Age at Menarche | 25231870 | 0.11 | 0.04 | 3.18 | 2 x10^-03^ |
|  | Mean Accumbens | 25607358 | 0.40 | 0.14 | 2.88 | 4 x10^-03^ |
|  | Body mass index | 20935630 | -0.11 | 0.04 | -2.78 | 0.01 |
|  | Extreme Body mass Index (BMI) | 23563607 | -0.15 | 0.06 | -2.44 | 0.01 |
|  | Crohns disease | 26192919 | 0.11 | 0.04 | 2.54 | 0.01 |
|  | Mean Putamen | 25607358 | 0.17 | 0.07 | 2.37 | 0.02 |
|  | Alzheimers disease | 24162737 | -0.25 | 0.11 | -2.35 | 0.02 |
|  | Obesity class 1 | 23563607 | -0.09 | 0.04 | -2.19 | 0.03 |
|  | Obesity class 2 | 23563607 | -0.11 | 0.05 | -2.12 | 0.03 |
|  | Lung adenocarcinoma | 27488534 | -0.25 | 0.11 | -2.17 | 0.03 |
|  | Urinary albumin-to-creatinine ratio (non-diabetes) | 26631737 | -0.20 | 0.09 | -2.24 | 0.03 |
|  | Excessive daytime sleepiness | 27992416 | -0.12 | 0.05 | -2.23 | 0.03 |
|  | Mean Thalamus | 25607358 | 0.22 | 0.11 | 1.96 | 0.04 |
| Anterior amygdaloid area | 18:2 linoleic acid (LA) | 27005778 | 0.27 | 0.11 | 2.38 | 0.02 |
|  | Height Females at age 10 and males at age 12 | 23449627 | -0.14 | 0.07 | -2.20 | 0.03 |
|  | Childhood Intelligence Quotient (IQ) | 23358156 | 0.20 | 0.09 | 2.15 | 0.03 |
|  | Idiopathic pulmonary fibrosis | 29066090 | -0.32 | 0.15 | -2.20 | 0.03 |
|  | Concentration of small Low Density Lipoprotein (LDL) particles | 27005778 | 0.23 | 0.11 | 2.14 | 0.03 |
|  | Childhood obesity | 22484627 | -0.14 | 0.07 | -2.10 | 0.04 |
|  | Free cholesterol | 27005778 | 0.46 | 0.22 | 2.10 | 0.04 |
|  | Triglycerides in very large High Density Lipoprotein (HDL) | 27005778 | 0.22 | 0.11 | 2.10 | 0.05 |
|  | Total lipids in small LDL | 27005778 | 0.24 | 0.12 | 1.97 | 0.06 |
| Accessory basal nucleus | Sleep duration | 27494321 | -0.16 | 0.06 | -2.70 | 0.01 |
|  | Child birth length | 25281659 | 0.20 | 0.08 | 2.42 | 0.02 |
|  | Insomnia | 28604731 | 0.13 | 0.06 | 2.33 | 0.02 |
|  | Birth weight | 27680694 | 0.09 | 0.04 | 2.15 | 0.03 |
|  | Height 2010 | 20881960 | 0.08 | 0.04 | 2.05 | 0.04 |
| Basal nucleus | Mothers age at death | 27015805 | 0.20 | 0.09 | 2.15 | 0.03 |
|  | Height Females at age 10 and males at age 12 | 23449627 | 0.14 | 0.07 | 2.10 | 0.04 |
|  | Birth weight | 27680694 | 0.08 | 0.04 | 1.97 | 0.05 |
| Corticoamygdaloid transition area | Insomnia | 27992416 | 0.19 | 0.07 | 2.74 | 0.01 |
|  | Height 2010 | 20881960 | 0.11 | 0.05 | 2.34 | 0.02 |
|  | Average number of double bonds in a fatty acid chain | 27005778 | -0.28 | 0.12 | -2.37 | 0.02 |
|  | Description of average fatty acid chain length not actual carbon number | 27005778 | -0.40 | 0.18 | -2.24 | 0.02 |
|  | Ratio of bisallylic groups to total fatty acids | 27005778 | -0.24 | 0.12 | -2.08 | 0.04 |
|  | Acetate | 27005778 | -0.39 | 0.19 | -2.05 | 0.04 |
|  | Insomnia | 28604731 | 0.17 | 0.08 | 2.10 | 0.04 |
|  | Mean Thalamus | 25607358 | 0.31 | 0.16 | 1.98 | 0.05 |
| Central nucleus | Mean Pallidum | 25607358 | 0.40 | 0.12 | 3.24 | 1 x10^-03^ |
|  | Waist-to-hip ratio | 25673412 | 0.12 | 0.05 | 2.47 | 0.01 |
|  | Omega-3 fatty acids | 27005778 | -0.29 | 0.12 | -2.49 | 0.01 |
|  | 22:6 docosahexaenoic acid | 27005778 | -0.26 | 0.12 | -2.12 | 0.03 |
| Cortical nucleus | Insomnia | 27992416 | 0.20 | 0.07 | 2.88 | 4 x10^-03^ |
|  | Sleep duration | 27494321 | -0.18 | 0.07 | -2.60 | 9 x10^-03^ |
|  | Age of smoking initiation | 20418890 | -0.37 | 0.15 | -2.51 | 0.01 |
|  | Height 2010 | 20881960 | 0.10 | 0.04 | 2.39 | 0.02 |
|  | Insomnia | 28604731 | 0.16 | 0.07 | 2.16 | 0.03 |
|  | Waist circumference | 25673412 | 0.10 | 0.05 | 2.07 | 0.04 |
|  | Waist-to-hip ratio | 25673412 | 0.10 | 0.05 | 2.06 | 0.04 |
|  | Child birth length | 25281659 | 0.19 | 0.10 | 2.05 | 0.04 |
|  | Forced expiratory volume in 1 second (FEV1) | 21946350 | 0.18 | 0.09 | 2.01 | 0.04 |
|  | Alanine | 27005778 | 0.20 | 0.10 | 2.03 | 0.04 |
|  | Number of children ever born | 27798627 | 0.12 | 0.06 | 1.99 | 0.04 |
| Lateral nucleus | Height 2010 | 20881960 | -0.09 | 0.04 | -2.14 | 0.03 |
| Medial nucleus | Obesity class 1 | 23563607 | 0.19 | 0.05 | 3.66 | 2 x10^-04^ |
|  | Overweight | 23563607 | 0.19 | 0.05 | 3.70 | 2 x10^-04^ |
|  | Body mass index | 20935630 | 0.18 | 0.05 | 3.48 | 5 x10^-04^ |
|  | Waist circumference | 25673412 | 0.16 | 0.05 | 3.31 | 9 x10^-04^ |
|  | Waist-to-hip ratio | 25673412 | 0.15 | 0.05 | 3.23 | 1 x10^-03^ |
|  | Leptin not adjBMI | 26833098 | 0.25 | 0.09 | 2.89 | 4 x10^-03^ |
|  | Obesity class 2 | 23563607 | 0.16 | 0.07 | 2.45 | 0.01 |
|  | Hip circumference | 25673412 | 0.11 | 0.05 | 2.32 | 0.02 |
|  | Crohn’s disease | 26192919 | 0.14 | 0.06 | 2.26 | 0.02 |
|  | Mean Pallidum | 25607358 | 0.26 | 0.12 | 2.28 | 0.02 |
|  | Lung cancer (all) | 24880342 | 0.21 | 0.09 | 2.32 | 0.02 |
|  | Omega-3 fatty acids | 27005778 | -0.28 | 0.12 | -2.35 | 0.02 |
|  | Age of first birth | 27798627 | -0.14 | 0.06 | -2.38 | 0.02 |
|  | HOMA-IR | 20081858 | 0.25 | 0.11 | 2.17 | 0.03 |
|  | Urinary albumin-to-creatinine ratio | 26631737 | 0.26 | 0.12 | 2.15 | 0.03 |
|  | Difference in height between adolescence and adulthood age 14 | 23449627 | -0.22 | 0.11 | -2.00 | 0.04 |
|  | Ratio of bisallylic groups to double bonds | 27005778 | -0.22 | 0.11 | -2.04 | 0.04 |
|  | Triglycerides in very large Very Low Density Lipoprotein (VLDL) | 27005778 | 0.19 | 0.10 | 1.99 | 0.04 |
|  | Age of smoking initiation | 20418890 | -0.30 | 0.15 | -2.03 | 0.04 |
| Paralaminar nucleus | Forced expiratory volume in 1 second (FEV1) | 21946350 | -0.23 | 0.09 | -2.55 | 0.01 |
|  | Fathers age at death | 27015805 | -0.21 | 0.09 | -2.34 | 0.02 |
|  | Child birth length | 25281659 | -0.19 | 0.08 | -2.34 | 0.02 |
|  | Sleep duration | 27494321 | 0.14 | 0.06 | 2.34 | 0.02 |
|  | Sitting height ratio | 25865494 | -0.14 | 0.07 | -2.14 | 0.03 |
|  | Apolipoprotein A-I | 27005778 | 0.27 | 0.12 | 2.18 | 0.03 |
|  | Albumin | 27005778 | 0.29 | 0.13 | 2.15 | 0.03 |
|  | Age of first birth | 27798627 | 0.10 | 0.04 | 2.23 | 0.03 |
|  | Ever vs never smoked | 20418890 | 0.14 | 0.07 | 2.17 | 0.03 |
|  | Total cholesterol in HDL | 27005778 | 0.20 | 0.10 | 2.07 | 0.04 |
|  | Free cholesterol in medium HDL | 27005778 | 0.22 | 0.11 | 2.06 | 0.04 |
|  | Smoking Initiation | 30617275 | 0.19 | 0.09 | 2.03 | 0.04 |
|  | Lung cancer (squamous cell) | 24880342 | -0.29 | 0.15 | -1.98 | 0.05 |
|  | Phospholipids in medium HDL | 27005778 | 0.21 | 0.11 | 1.99 | 0.05 |
|  | Age of smoking initiation | 20418890 | 0.25 | 0.13 | 1.97 | 0.05 |

**Significant at corrected p<2.63x10^-5^*
